# Subclonal *CHD1* deletion is more frequent in African American prostate cancers and associated with rapid disease progression and limited levels of homologous recombination deficiency

**DOI:** 10.1101/2021.02.08.21251199

**Authors:** Miklos Diossy, Viktoria Tisza, Hua Li, Jia Zhou, Zsofia Sztupinszki, Denise Young, Darryl Nousome, Claire Kuo, Jiji Jiang, Yongmei Chen, Reinhard Ebner, Isabell A. Sesterhenn, Joel T. Moncur, Gregory T. Chesnut, Gyorgy Petrovics, Gregory T. Klus, Gabor Valcz, Pier Vitale Nuzzo, Dezso Ribli, Aimilia Schina, Judit Börcsök, Aurel Prosz, Marcin Krzystanek, Thomas Ried, David Szuts, Salma Kaochar, Shailja Pathania, Alan D. D’Andrea, Istvan Csabai, Shiv Srivastava, Matthew L Freedman, Albert Dobi, Sandor Spisak, Zoltan Szallasi

## Abstract

Chromodomain helicase DNA-binding protein 1 (*CHD1*) is frequently deleted in a subset of prostate cancers. We show here that subclonal deletion of *CHD1* is nearly three times as frequent in prostate tumors of African American men than in men of European ancestry and it associates with rapid disease progression. We further show that *CHD1* deletion is associated with only one type of the homologous recombination deficiency associated mutational signatures in prostate cancer. In prostate cancer cell line models *CHD1* deletion did not induce HRD as detected by RAD51 foci formation assay, which was consistent with the moderate increase of olaparib sensitivity. *CHD1* deficient prostate cancer cells, however, still showed markedly increased sensitivity to talazoparib, which could be effective for the treatment of *CHD1* deficient tumors in the context of hormone therapy resistant prostate cancer.

## Introduction

African American (AA) men have significantly higher incidence and mortality rates from prostate cancer (PCa) compared to individuals of European ancestry (EA)^1^. Recent studies demonstrated that AA men are at higher risk of progression after radical prostatectomy, even in equal access settings and when accounting for socioeconomic status^2, 3^. While the reasons underlying these disparities are multifactorial, these data strongly argue that germline and/or somatic genetic differences between AA and EA men may in part explain these differences.

Comparative analysis of AA and EA prostate tumors have identified several genomic differences. *PTEN* deletions, *ERG* rearrangements and consequent *ERG* over-expression are more frequent in PCas of EA men^4–6^. In contrast, *LSAMP* and *ETV3* deletions, *ZFHX3* mutations, *MYC* and *CCND1* amplifications and *KMT2D* truncations are more frequent in PCas of AA men^7–9^. *ERF*, an ETS transcriptional repressor, also showed an increased mutational frequency in AA prostate cancer cases with probable functional consequences such as increased anchorage independent growth^10^, and *SPINK1* expression is also enriched in African American PCa^11^.

Chromodomain helicase DNA-binding protein 1 (*CHD1*) deletion is frequently present in prostate cancer. Deletions are associated with increased Gleason score and faster biochemical recurrence^12^, activation of transcriptional programs that drive prostate tumorigenesis^13^ and enzalutamide resistance^14^. A recent publication showed that CHD1 loss is also associated with increased risk of metastases at diagnosis ^15^.

Mechanistically, *CHD1* loss influences prostate cancer biology in at least two ways. *CHD1*, an ATPase-dependent chromatin remodeler, contributes to a specific distribution of androgen receptor (AR) binding in the genome of prostate tissue. When lost, the AR cistrome redistributes to HOXB13 enriched sites and thus alters the transcriptional program of prostate cancer cells ^13^. *CHD1* also contributes to genome integrity. It is required for the recruitment of CtIP, an exonuclease, to DNA double strand breaks (DSB) to initiate end resection. Impairing this important step of DSB repair upon CHD1 loss was proposed to lead to homologous recombination deficiency^16, 17^. The functional impact of *CHD1* loss is likely further influenced by the presence of SPOP mutations, which were reported to be associated with the suppression of DNA repair^18^.

*CHD1* loss is frequently subclonal^19^ (present only in a subset of cells), which makes its detection by next generation sequencing more challenging^20^ and it may go undetected depending on the fraction of cells harboring this aberration. Therefore, the true proportion of PCa cases with *CHD1* may be underestimated. Thus, we decided to investigate the frequency of *CHD1* loss in EA and AA PCa by methods more sensitive to detecting subclonal deletions including evaluations of multiple tumor foci present in each prostatectomy specimen.

## RESULTS

### Subclonal *CHD1* deletion is more frequent in African American prostate cancers and associated with worse clinical outcome

*CHD1* is frequently subclonally deleted in prostate cancer ^19^. Our initial analysis on the SNP array data from TCGA comparing AA and EA PCa cases suggested that the subclonal loss of *CHD1* may be a more frequent event in AA men (Suppl. Figures 1 and 2). To independently validate this observation, we assessed *CHD1* copy number by FISH (for probe design see Suppl. Figure 3) in tissue microarrays (TMAs) sampling multiple tissue cores from each tumor focus. Sampling included index tumors and non-index tumors per whole mounted radical prostatectomy sections in a matched cohort of 91 AA and 109 EA patients from the equal-access military healthcare system (Figure 1a). The mean of age at diagnosis in AA patients is younger than EA patients, but other key clinico-pathological features including serum PSA levels at diagnosis, pathological T-stages, Gleason sums, Grade groups, margin status, biochemical recurrence (BCR) and metastasis had no significant differences between AA and EA cases (Suppl. Table 1a). Consistent with other studies, we observed a 40% biochemical recurrence (BCR) and 13% metastasis rate^21^. For each case up to four cancerous foci were analyzed, each sampled by two TMA punch cores on average (for details see Material and Methods and Suppl. Table 1b and 1c). We detected monoallelic *CHD1* loss in 27 out of 91 AA cases (29.7%), and 14 out of 109 (13%) EA cases indicating that *CHD1* deletion is about 2.3 times more frequent in prostate tumors of AA men. Our FISH data showed only 3 cases (2 AA and 1 EA) where all TMA punch cores in a single tumor focus harbored *CHD1* deletion in the entire samples areas of a given tumor. (Figure 1b and see Materials and Methods “FISH assay part” for details.) In most cases *CHD1* deletion was present in only a subset of tumor glands within a 1 mm TMA punch, which further confirmed the subclonal nature of *CHD1* deletion in prostate cancer. As a control, we performed FISH staining for *PTEN* deletion and immunohistochemistry (IHC) staining for ERG overexpression in a subset of the cohort (42 AA and 59 EA prostate cancer cases) confirming previously described frequency differences between AA and EA PCa^4, 5^ (Suppl. Table 1e). There was a frequent exclusivity between *CHD1* deletion, *PTEN* deletion and ERG expression both when individual tumor cores or when all tumor cores from a given patient were considered (Suppl. Figures 4a and 4b). In general, the genomic defects including *CHD1* deletion, *PTEN* deletion and ERG expression were mainly detected in index tumors.

**FIGURE 1:**
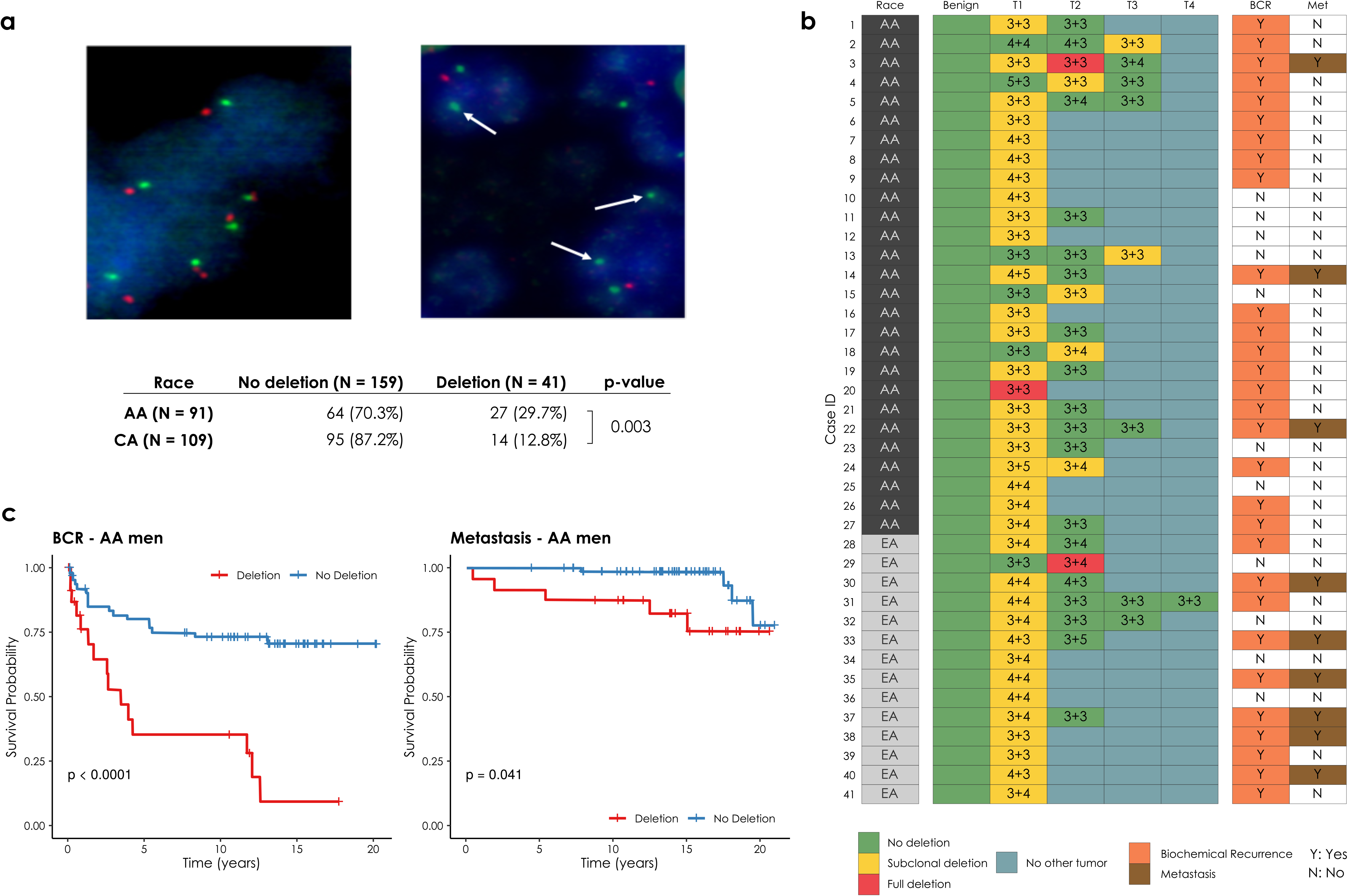
CHD1 copy number by FISH in tissue microarrays. **(a)** Prostate cancer cells with wild type (diploid) *CHD1* (upper left) vs. prostate cancer cells harboring mono-allelic deletion for *CHD1* (upper right) are visualized by FISH assay. Orange signal: *CHD1* probe; green signal: human chromosome 5 short arm probe; blue color: DAPI nuclear stain. Arrows are representing the lack of *CHD1* . Representative view fields capture 3-3 cell nuclei at 60X magnification. Inset table summarizes the higher frequency of *CHD1* deletion in prostatic carcinoma of AA vs. EA patients (The p-value is from a one-sided Fisher exact test). **(b)** *CHD1* deletion is a subclonal event in prostate cancer. Multiple tumor samples from 200 patients were assessed by FISH assay that identified 41 patients with *CHD1* deletion (left panel). The heatmap depicts the sampled largest tumor 1 (T1), second largest tumor (T2), and so on. Numbers denote pathological Gleason grade for each tumor. BCR: biochemical recurrence (orange); Met: metastasis (brown). **(c)** Deletion of *CHD1* (clonal or subclonal in any of the nodes) is strongly associated with disease progression in AA prostate cancer patients (N=91). BCR: univariable Kaplan-Meier curve; Metastasis: univariable Kaplan-Meier curve.

Further analyses revealed a significant association between *CHD1* deletion and pathologic stages and Gleason sum. Higher frequency of *CHD1* deletion was detected in T3-4 pathological stage compared to T2 stage (P=0.043, Suppl. Table 1d). Prostate cancer cases with higher Gleason sum scores (3+4, 4+3, 8-10) were seen more frequently in the *CHD1* deletion group than in the non-deletion group (P<0.001). In contrast, lower Gleason sum score (3+3) was more often seen in non-deletion cases (P<0.001, Suppl. Table 1d). The *CHD1* deletion was more commonly detected in the cases with higher grade group (GG3 and GG4-GG5) (P=0.024, Suppl. Table 1d). *CHD1* deletion was strongly associated with biochemical recurrence in AA cases (P<0.0001, Figure 1c) but it showed a marginal significance in EA cases (P=0.051, Suppl Figure 5a). The univariable survival analysis was conducted to assess the association of the clinical features including *CHD1* deletion with BCR and metastasis for further multivariable model analysis (Suppl. Figures 5b and c, respectively). The multivariable Cox model analysis showed that *CHD1* deletion was an independent predictor of BCR (Gleason Group P=0.012 or Gleason Score P=0.032, Suppl. Figure 5d) after adjusting for age at diagnosis, PSA at diagnosis, race, surgical margins and either grade group or Gleason scores. Moreover, statistically significant correlation between *CHD1* deletion and metastasis were also detected in both AA (P=0.041, Figure 1d) and EA (P=0.023, Suppl. Figure 5e) patients with Kaplan-Meier analysis. Following multivariable Cox proportional hazards model, controlling for other clinical features, *CHD1* deletion was significantly associated with metastasis (Gleason Group P=0.032 or Gleason Score and P=0.048, Suppl. Figure 5f). Taken together, our data strongly support the higher frequency of *CHD1* deletions in AA compared to EA patients, and the association of CHD1 deletions with aggressive prostate cancer and worse clinical outcomes in AA PCa.

### Estimating the frequency of subclonal *CHD1* loss in next generation sequencing data of AA and EA prostate cancer

Previous publications characterizing the genome of AA prostate cancer cases ^10, 22^ did not report an increased frequency of *CHD1* loss as we observed in the FISH-based analysis presented above. Methods to detect copy number variations from WGS or WES data have at least two major limitations. First, subclonal copy number variations (sCNV) can be missed if they are present in fewer than 30%, of the sampled cells^20^. Second, copy number loss can be underestimated with smaller deletions (e.g., <10 kb). Although various tools are available for inferring sCNVs from WES, WGS or SNP array data, such as TITAN^20^, THetA^23^, and Sclust^24^, they are designed to work on the entire genome, and likely miss small (∼1-10kb) CNVs during the data segmentation process. In order to maximize the accuracy of our analysis we performed a gene focused analysis of the copy number loss in *CHD1* . We considered several factors such as the change in the normalized coverage in the tumors relative to their normal pairs’, the cellularity of the tumor genome, and the approximate proportion of tumor cells exhibiting the loss. We also evaluated whether the deletion was heterozygous or homozygous using a statistical method designed for calling subclonal loss of heterozygosity (LOH) events within a confined genomic region (details are available in the Materials and Methods section, and in the Supplementary Material).

Using this approach in a large cohort (N=530 cases; 59 AA WES, 18AA WGS, 408 EA WES and 45 EA WGS, for details see supplementary material and Suppl. Figures 6-25), we observed that *CHD1* is more frequently deleted in AA tumors (N=20; 26%) than in EA tumors (N=73 EA; 16%). Taken together, when next generation sequencing based copy number variations were analyzed with a more sensitive method, on the combined cohorts of whole exomes and whole genomes, *CHD1* loss was detected more frequently in AA cases than in EA cases (p=0.029, Fisher exact test), which is consistent with our observations with FISH method in the TMA cohort.

### *CHD1* loss is associated with only one of the types of genomic aberration features frequently observed in *BRCA2-* deficient prostate cancers

*CHD1* loss was proposed to be associated with reduced HR competence in cell line model systems^16, 25^. Detecting and quantifying HR deficiency in tumor biopsies is currently best achieved by analyzing whole genome sequencing data for specific HR deficiency associated mutational signatures. Those include: 1) A single nucleotide variation based mutational signature (“COSMIC signatures 3^26^ and SBS3^27^); 2) a short insertions/deletions based mutational profile, often dominated by deletions with microhomology, a sign of alternative repair mechanisms joining double-strand breaks in the absence of HR, which is also captured by COSMIC indel signatures ID6 and ID8^27^; 3) large scale rearrangements such as non-clustered tandem duplications in the size range of 1-100kb (mainly associated with *BRCA1* loss of function) or deletions in the range of 1kb-1Mb (mainly associated with *BRCA2* loss of function)^28^. All of these signatures can be efficiently induced by the inactivation of *BRCA1*, *BRCA2* or several other key downstream HR genes (Suppl. Figures 26-44) ^29^ . HR deficiency is also assessed in the clinical setting by a large scale genomic aberration based signature, namely the HRD score^30^, which is also approved as companion diagnostic for PARP inhibitor therapy. Recently a composite mutational signature, HRDetect^31^, combining several of the mutational features listed above was evaluated as an alternative method to detect HR deficiency in prostate adenocarcinoma^32^. In order to further strengthen the link between *CHD1* loss, HR deficiency and potentially increased PARP inhibitor sensitivity we performed a detailed analysis on the mutational signature profiles of *CHD1* deficient prostate cancer.

We analyzed whole exome and whole genome sequencing data of several prostate adenocarcinoma cohorts (For the detailed results see the Supplementary Material) containing samples both from AA (52 WES and 18 WGS cases) and EA (387 WES and 45 WGS) individuals in order to determine whether *CHD1* loss is associated with the HRD mutational signatures.

We divided the cohorts into three groups: 1) *BRCA2* deficient cases that served as positive controls for HR deficiency, 2) *CHD1* deleted cases without mutations in HR genes, and 3) cases without *BRCA* gene aberration or *CHD1* deletion (for details see Supplementary Material).

In the WGS cohorts *CHD1* deficient cases showed increased HRD score relative to the control cases but lower than the *BRCA2* deficient cases and none of the *CHD1* deficient cases had an HRD score above the threshold currently accepted in the clinic as an indicator of HR deficiency (Figure 2a). It is important to emphasize, however, that the HRD score was positively correlated with the estimated fraction of the subclonal loss of *CHD1* (Figure 2a, Suppl. Figure 26-27), suggesting that the signal of HRD score was “diluted” likely due to decreased *CHD1* deleted tumor cells/tissue sample ratio. The most characteristic HRD associated single nucleotide variation signature (signature 3), was significantly increased in the *BRCA2* deficient cases and slightly increased in the CHD1 deficient cases (Figure 2b). The increase of the relative contribution of short indel signatures ID6 and ID8 to the total number of indels characteristic of loss of function on BRCA2 biallelic mutants was not observed in the CHD1 loss cases (Suppl. Fig. 32-34). This suggests, that the alternative end-joining repair pathways do not dominate the repair of DSBs in *CHD1* deleted tumors.

**FIGURE 2:**
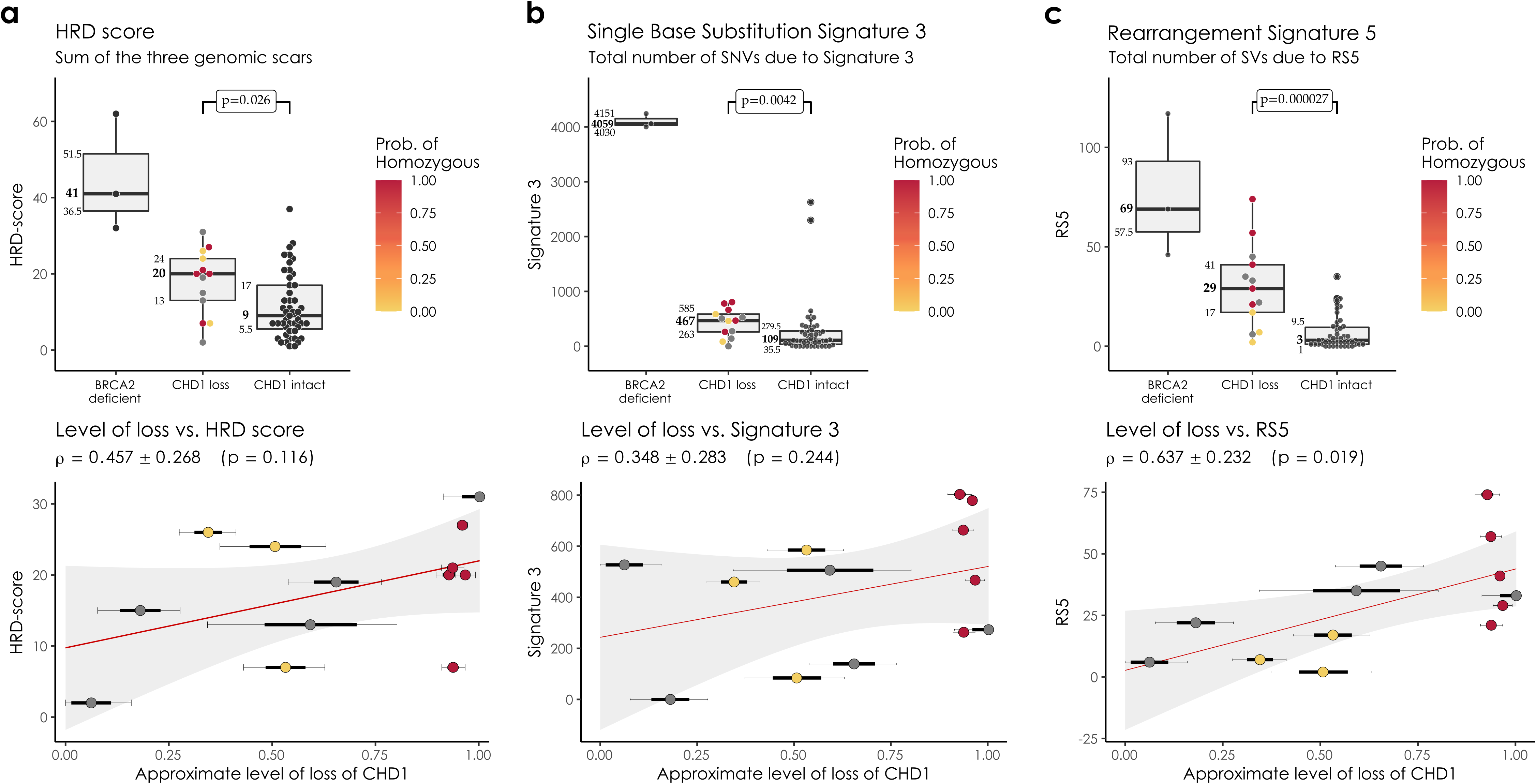
HRD markers in the PRAD WGS cohorts. **(a)** HRD-score, the sum of the three genomic scars, HRD-LOH, LST, and ntAI, **(b)** n*umber of somatic mutations due to single-base substitution signature 3, **(c)** n*umber of structural variants due to rearrangement signature 5. The significance of the difference between the means of the “*CHD1* loss” and “control” groups were assessed with Wilcoxon ranked sum tests. Below the box plots are the correlations between the approximate levels of loss in *CHD1* and the HRD measures are visualized. The standard errors and the corresponding p-values of the correlation coefficients (Pearson) are also indicated. Horizontal lines indicate the uncertainty in the level of loss in each sample. Thick black lines correspond to the 66%, thin black error-bars to the 95% percentile intervals.

In the WGS cohort we also determined the number of structural variants as previously defined (Suppl. Fig. 35)^28^. As expected, RS5 was significantly increased in the *BRCA2* mutant cases since this signature (an increase in the number of non-clustered 1kb-1Mb deletions) was identified as a specific feature of such tumors. *CHD1* deficient cases also displayed a significant increase in RS5 structural variations but the signal showed a strong subclonal dilution (Figure 2c). Finally, the *BRCA2* deficient cases showed high HRDetect scores (Suppl. Figures 36-38). However, since the HRDetect scores arise from a logistic regression, which involves the non-linear transformation of the weighted sum of its attributes, even slightly lower linear sums in the *CHD1* loss cases compared to the *BRCA2* mutant cases can result in substantially lower HRDetect scores (Suppl. Figure 38).

We have previously processed WES prostate adenocarcinoma data for the various HR deficiency associated mutational signatures^32^. When the *CHD1* deficient cases were compared to the *BRCA1/2* deficient and *BRCA1/2* intact cases we obtained results that were consistent with the WGS based results outlined above (Suppl. Figures 39-44).

### Deleting *CHD1* in prostate cancer cell lines does not induce homologous recombination deficiency as detected by the RAD51 foci formation assay or mutational signatures

In order to investigate the functional impact of the biallelic loss of CHD1 we created several CRISPR-Cas9 edited clones of the AR-PC-3 and AR+ 22Rv1 cell lines (Figure 4a, Suppl Figure 47a). RAD51 foci formation was induced by 4Gy irradiation. The CHD1 deficient prostate cancer cell lines did not show reduction of RAD51 foci formation. (Figure 3a). As controls, non-irradiated cells were used (Suppl Figure 46) DNA repair pathway aberration induced mutational signatures can also be detected in cell lines by whole genome sequencing^29, 33^. We grew single cell clones from the PC-3 and 22Rv1 cell lines for 45 generations to accumulate the genomic aberrations induced by CHD1 loss (Suppl. Figure 45). Two of such late passage clones and an early passage clone were subjected to WGS analysis. All the clones retained the *BRCA2* wild type background of their parental clone.

**FIGURE 3:**
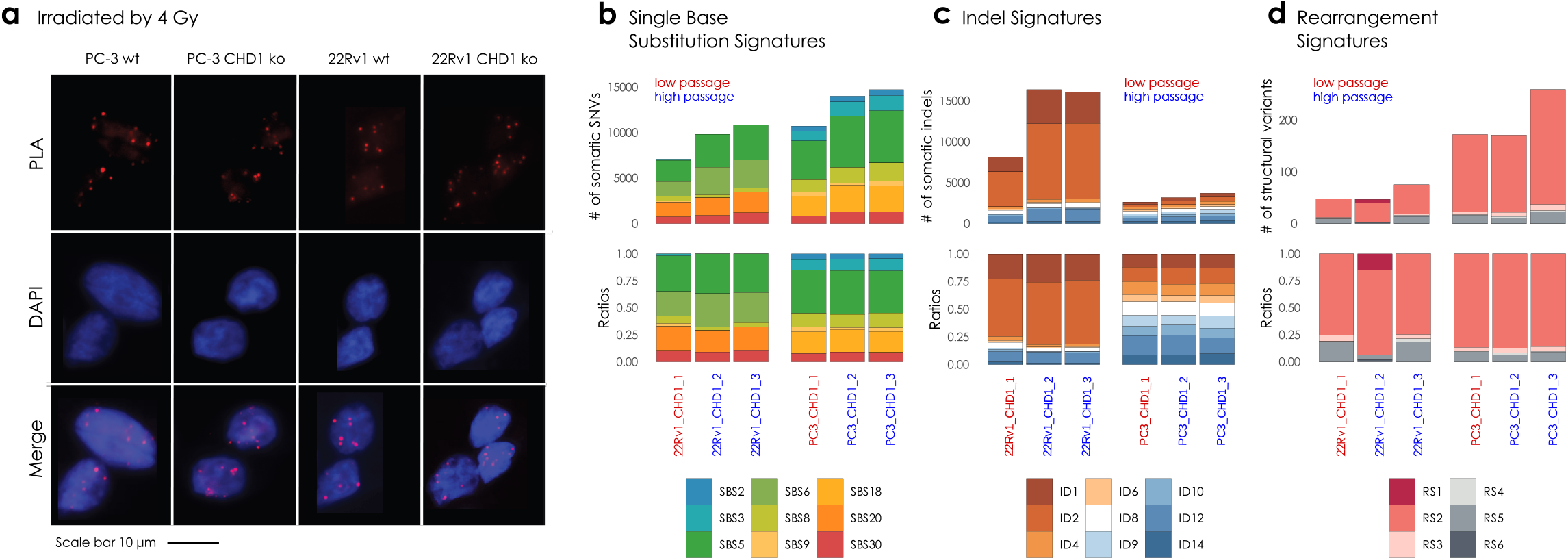
PC-3 and 22Rv1 CHD1 ko cell line experiment and somatic signature extraction. **(a)** RAD51 foci formation. Examples of the most common staining patterns in WT and CHD1 ko 22Rv1 and PC-3 cell lines. Cells were fixed by 4% PFA 3hrs afte irradiation (IR=4Gy) PLA was carried out using antibodies against γH2Ax and RAD51 proteins. **(b)** Single Nucleotide Substitution (SBS) signatures, **(c)** Indel signatures, **(d)** Rearrangement signatures. The number of mutations indicated originate from the reconstructed mutational spectra.

**FIGURE 4:**
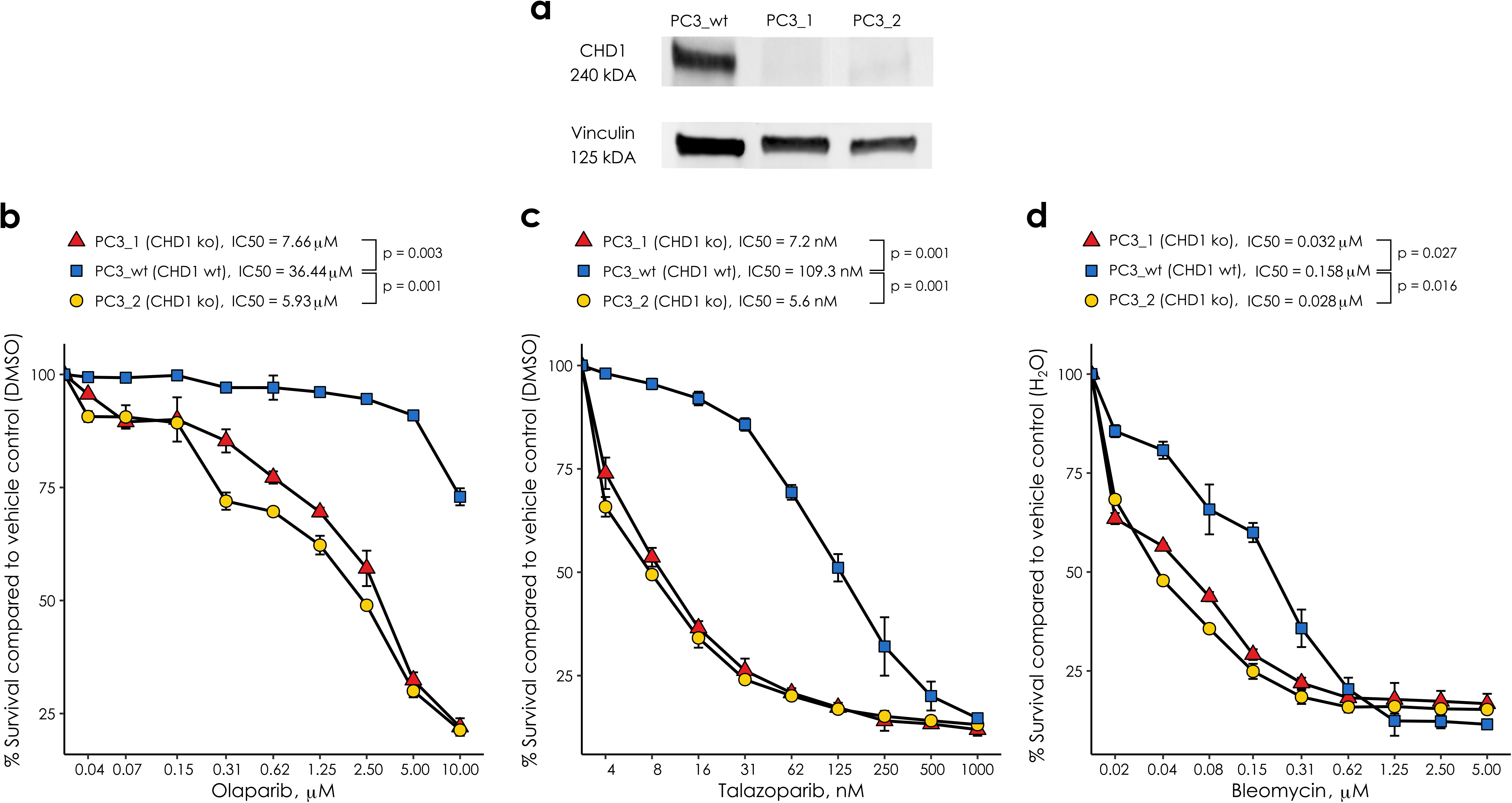
CHD1 loss PC-3 prostate cells show significant response to HR deficiency directed therapy. **(a)** Immunoblot shows that *CHD1* was successfully knocked out in PC-3 cells. Sensitivity assays of parental wt and *CHD1* ko clones to PARP inhibitor Olaparib **(b)**, Talazoparib **(c)**, and the radiomimetics bleomycin sulfate **(d)**. Cells viability was measured using PresoBlue^TM^ reagent. SD of triplicates are shown, p-values were calculated using student’s t-test. p-values <0.05 were considered statistically significant.

CHD1 elimination did not induce any of the mutational signatures commonly associated with HR deficiency (Figure 3b-d) Taken together, *CHD1* loss in prostate cancer cell line model systems did not induce robust signs of HR deficiency.

### *CHD1* deficient cell lines show increased sensitivity to talazoparib and the radiomimetic agent bleomycin

*CHD1* deficient cancer cells were reported to have moderately increased sensitivity to the PARP inhibitor olaparib^16^, which is consistent with the lack of observed HR deficiency described in the previous section. PARP inhibitors were initially thought to exert their therapeutic activity by inhibiting the enzymatic activity of PARP, but it was later revealed that trapped PARP on DNA may have a more significant contribution to cytotoxicity (reviewed in^34^). Therefore, we tested the efficacy of the strong PARP trapping agent talazoparib in the prostate cancer cell lines PC-3 and 22Rv1 with or without CRISPR-Cas9-mediated *CHD1* deletion. Consistent with previous reports, deleting CHD1 induced an approximately 5-fold increase in olaparib sensitivity in PC-3 (Figure 4b)^16^ but no increase in the CHD1 deficient 22Rv1 cells (Suppl. Figure 47b). In contrast, the sensitivity to talazoparib increased by about 15-20-fold in the same *CHD1* deficient PC-3 cells (Figure 4c), with no increase in the 22Rv1 cells again (Suppl. Figure 47c).

These data suggest that trapped PARP may have a more toxic effect in cells with *CHD1* deficiency in AR-negative prostate cancer cells.

Consistent with the significant functional evidence linking *CHD1* deletion and HR repair of DSBs, *CHD1* deficient cells also showed increased sensitivity to irradiation^16^. We investigated, whether this increased sensitivity also applies to chemotherapy agents that induce DSBs, such as radiomimetic drugs. As shown in Figure 4d, *CHD1* deficient PC-3 cells show significantly increased (5-fold) sensitivity to bleomycin, the most frequently used radiomimetic therapeutic agent. However, we observed no increase in 22Rv1 cells again (Suppl. Figure 47d).

### The impact of SPOP mutations on the clonality of *CHD1* deletions and HR deficiency associated mutational signatures

Although less frequent, SPOP mutations and *CHD1* deletions may co-exist in a subset of prostate cancer^35^ and SPOP mutations have been shown to suppress key HR genes^18^. Therefore, we investigated whether the presence of *SPOP* mutation in a *CHD1* deficient prostate cancer is associated with a further increase of HR deficiency associated mutational signatures. We identified cases with *SPOP* mutations or *CHD1* deletions only, cases with both *SPOP* mutations and *CHD1* deletions and cases without either of those aberrations (Figure 5a). Cases with both mutations showed significantly higher levels of signature SBS3, RS5 and the total number of large-scale structural rearrangements relative to cases with either mutation alone. It should be noted, however, that the proportion of cells in a given tumor with *CHD1* deletions tended to be significantly higher in *SPOP* mutant cases than those with *CHD1* deletions without *SPOP* mutations. Thus, considering the previously demonstrated impact of *CHD1* subclonality on the intensity of HR deficiency associated mutational signatures (Figure 2), it is possible that the presence of *SPOP* will intensify HR deficiency associated mutational signatures by enhancing the proportion of *CHD1* deficient cells in a tumor (Figure 5b).

**FIGURE 5:**
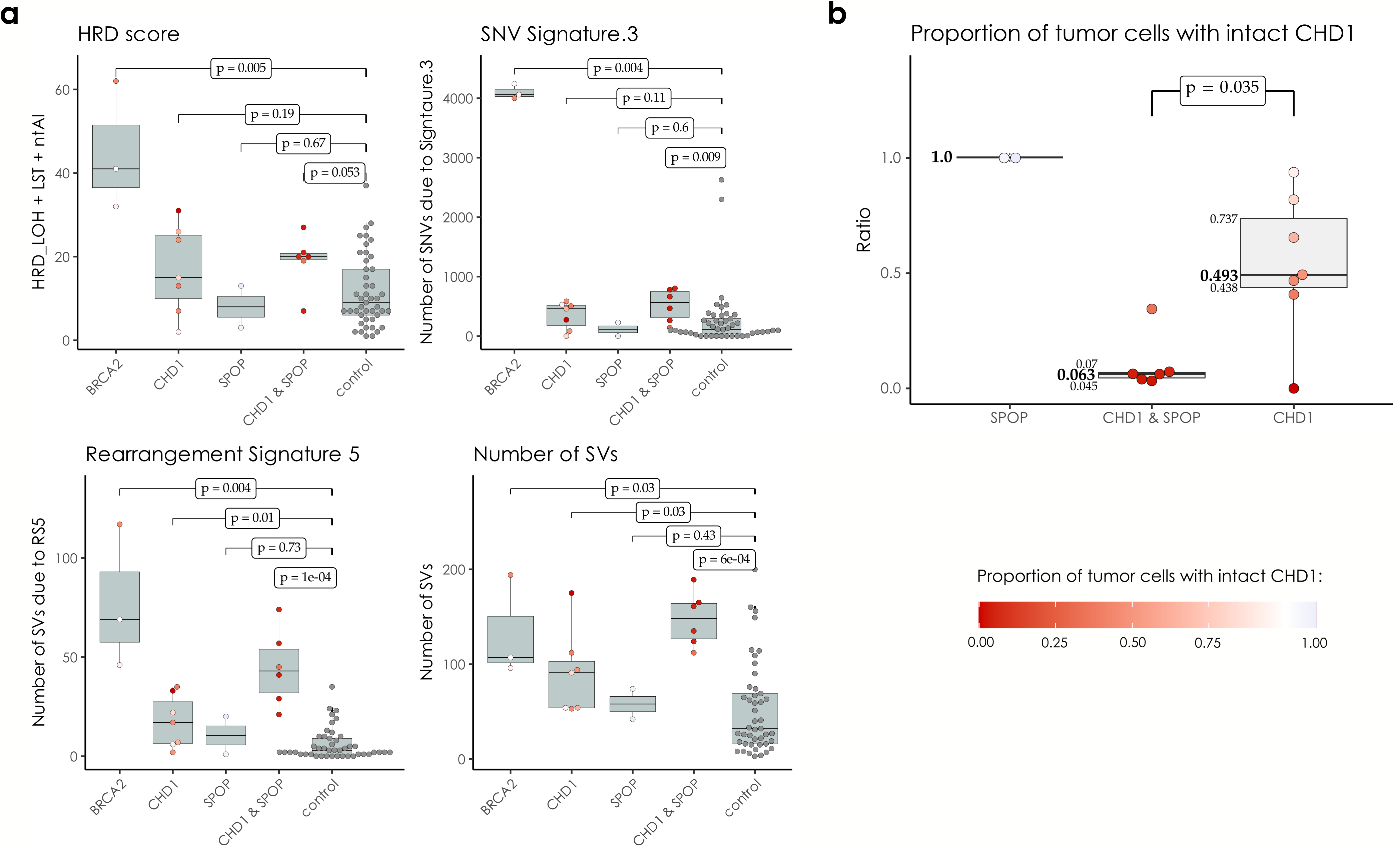
CHD1 loss and SPOP mutation in the WGS cohorts. **(a)** HRD-related markers and total number of structural variants in samples with mutations in *SPOP*, *BRCA2* and loss in *CHD1* versus the controls. Samples that simultaneously harbor mutations in *SPOP* and a loss in *CHD1* tend to have higher markers. P-values were estimated using non-parametric Wilcoxon signed-rank tests. **(b)** Proportion of cells with intact *CHD1* in *SPOP* mutants and samples identified with *CHD1* loss. While the deletion in *CHD1* in *SPOP* mutants is mostly clonal, in samples with wild type *SPOP* background it is mostly subclonal. The color-code for points in both panels A and B is illustrated in the bottom right corner of the figure.

## DISCUSSION

The presence of functionally relevant subclonal mutations in various solid tumor types is well documented^36, 37^. Deletions present only in a minority of tumor cells are difficult to detect unless more targeted analytical approaches are applied. Here we present one example of such detection bias with significant functional relevance. We used a FISH based approach to detect *CHD1* deletion in PCa. Consistent with the previously described subclonal nature of *CHD1* loss, we found that while this gene is often deleted in prostate cancer, it is rarely deleted in every tumor core or tumor focus. When we took the subclonal nature of *CHD1* loss into consideration a significant racial disparity emerged, with an approximately 3-fold increase in the frequency of CHD1 deletion in AA PCa patients vs EA patients. This loss was also significantly associated with rapid disease progression to biochemical recurrence and metastasis. Since *CHD1* loss is associated with a more malignant phenotype, the significantly higher frequency of *CHD1* loss in AA PCa may account for the diverging clinical course observed in PCa between men of African and European Ancestry. It is possible that *CHD1* loss is in fact more frequent in EA PCa as well but with a lower focal density than in AA cases. This is certainly a limitation of our bioinformatics approach. However, CHD1 single cell-level deletions have not been observed in our high-resolution FISH assay in tumors of EA patients.

Several studies pointed out a potentially intimate link between *CHD1* loss and homologous recombination deficiency^16, 17, 25^. Interestingly, *CHD1* null cells showed only a modest (3-fold) increase in sensitivity to olaparib or platinum-based therapy^16, 17, 25^. This suggested that *CHD1* loss may not lead to the same level or the same completeness of HR deficiency as that detected upon loss of function of *BRCA1* or *BRCA2* . The loss of function of those key HR genes usually leads to various DNA repair deficiencies such as stalled fork destabilization or reduced capacity of DSB repair. The presence of those DNA repair deficiencies can often be detected by different types of DNA aberration profiles, and they can be associated with an up to 1000-fold increase in PARP inhibitor sensitivity. The modest increase in PARP inhibitor sensitivity suggests that *CHD1* loss may lead to some but not all DNA repair aberrations usually associated with loss of function of *BRCA1/2* . Indeed, *CHD1* deficient tumors displayed strong signals of the *BRCA2* deficiency associated structural variation signature (SV5), but only modest or no increase of the single nucleotide variation or short indel based signatures. This suggests, that *CHD1* loss “mimics” some but not all of the consequences of *BRCA2* deficiency. It is also possible that *CHD1* is predominantly deleted in tumors with loss of function BRCA2 like-features. The precise mechanistic nature of this similarity needs further clarifications in mechanistic studies.

Identification of synthetic lethal agents with *CHD1* deficiency is expected to benefit those prostate cancer cases that harbor this aberration. In early clinical studies, patients with *CHD1* deficient prostate cancer responded to PARP inhibitor and platinum-based therapy^25^. However, the subclonal nature of *CHD1* loss we have highlighted here may have considerable clinical consequences. Tumors with *CHD1* loss in a significant subset of the cells may show significant response to HR deficiency directed therapy. HR deficiency associated mutational signatures are used to prioritize ovarian cancer patients for PARP inhibitor therapy and a similar strategy may be considered for prostate cancer as well. However, as we showed here, the HR deficiency associated mutational signatures are “diluted out” proportionally to the subclonality of *CHD1* loss. Therefore, diagnostic cut-off values may need to be readjusted for *CHD1* deficient cases as well as cellular level of diagnostic confirmation by FISH assay will be necessary for diagnostic consideration. If only a smaller subset of tumor cells harbor *CHD1* deficiency, then a synthetic lethal agent may have only a modest benefit in terms of tumor shrinkage. However, as it was suggested recently, *CHD1* loss may play a key role developing enzalutamide resistance^14^. Therefore, it is possible that eliminating the subset of *CHD1* deleted tumor cells, even if a minority, will significantly delay antiandrogen therapy resistance. Moreover, the majority of specimens analyzed for *CHD1* deletions in our study represent treatment naïve primary prostate tumor specimens. It would be reasonable to expect clonal expansion of *CHD1* deleted tumor cells in more advanced heavily treated metastatic and castration resistant prostate cancers (CRPC), which calls for further analysis. Therefore, agents with higher specificity for *CHD1* deficiency, such as talazoparib or bleomycin, may be used as agents to stave off resistance to antiandrogen therapy. Considering the higher frequency of *CHD1* loss in AA PCa, such *CHD1* directed therapy may stop the early development of more malignant clones and may reduce the racial differences in the overall outcome of prostate cancer.

## Materials and Methods

### Cohort selection and Tissue Microarray (TMA) generation

The aggregate cohort was composed of 2 independently selected cohort samples from Bio-specimen bank of Center for Prostate Disease Research and the Joint Pathology Center. Wholemount prostates were collected from 1996 to 2008 with minimal follow-up time of 10 years. The first cohort of 42 AA and 59 EA cases was described before^7, 38^. Similarly, the second cohort of 50 AA and 50 EA cases was selected based on the tissue availability (>1.0 cm tumor tissue) and tissue differentiation status (1/3 well differentiated, 1/3 moderately differentiated and 1/3 poorly differentiated). All the selected cases had the signed patient consent forms for tissue research applications. Patients who have donated tissue for this study also contributed to the long-term follow-up data (the mean follow-up time was 14.5 years). Our study was reviewed by the Uniformed Services University’s Human Research Protections Program (HRPP) Office and “determined to be considered research not involving human subjects as defined by 32 CFR 219.102(e) because the research involves the use of de-identified specimens and data not collected specifically for this study.” (Ref #910230). TMA block was assigned as 10 cases each slide and each case with 2 benign tissue cores, 2 Prostatic intraepithelial neoplasia (PIN) cores if available and 4-10 tumor cores covering the index and non-index focal tumors from formalin fixed paraffin embedded (FFPE) wholemount blocks. The description of numbers of patients, tumors and tumor cores of combined cohort was in Supplementary table 1d. All the blocks were sectioned into 8 µM tissue slides for FISH staining.

Fluorescence in situ hybridization (FISH) assay: A gene-specific FISH probe for *CHD1* was generated by selecting a combination of bacterial artificial chromosome (BAC) clones (Thermo Fisher Scientific, Waltham, MA) within the region of observed deletions near 5q15-q21.1, resulting in a probe matching ca. 430 kbp covering the CHD1 gene as well as some upstream and downstream adjacent genomic sequences including the complete repulsive guidance molecule B (RGMB) gene. Due to the high degree of homology of chromosome 5-specific alpha satellite centromeric DNA to the centromere repeat sequences on other chromosomes, and the resulting potential for cross-hybridization to other centromere sequences, particularly on human chromosomes 1 and 19, a control probe matching a stable genomic region on the short arm of chromosome 5 – instead of a centromere 5 probe - was used for chromosome 5 counting (supplementary figure 3). The FISH assay of *CHD1* was performed on TMA as previously described^7^. The green signal was from probe detecting control chromosome 5 short arm and the red signal was from probe detecting *CHD1* gene copy. The FISH-stained TMA slides were scanned with Leica Aperio VERSA digital pathology scanner for further evaluation. The criteria for *CHD1* deletion was that in over 50% of counted cancer cells (with at least 2 copies of chromosome 5 short arm detected in one tumor cell) more than one copy of *CHD1* gene had to be undetected. Total 50 counted cancer cells (with at least 2 copies of chromosome 5 short arm) from representative malignant glands or all available tumor cells if the number of tumor cells were below 50 in TMA tumor foci were used for assessing CHD1 deletion. Examining tumor cores, deletions were called when more than 75% of evaluable tumor cells showed loss of allele. Focal deletions were called when more than 25% of evaluable tumor cells showed loss of allele or when more than 50% evaluable tumor cells in each gland of a cluster of two or three tumor glands showed loss of allele. Benign prostatic glands and stroma served as built-in control.

The sub-clonality of *CHD1* deletion was presented with a heatmap showing *CHD1* deletion status in all the given tumors sampled from whole-mount sections of each patient. The color designations were denoted as: red color (full deletion) meaning all the tumor cores carrying *CHD1* deletion within a given tumor, yellow color (sub-clonal deletion) meaning only partial tumor cores carrying *CHD1* deletion within a given tumor and green color (no deletion) meaning no tumor core carry *CHD1* deletion (supplementary table 1b).

#### Statistical Analysis

The correlations of *CHD1* deletion and clinic-pathological features, including pathological stages, Gleason score sums, Grade groups, margin status, and therapy status were calculated using an unpaired t-test or chi-square test. Gleason Grade Groups were derived from the Gleason patterns for cohort from Grade group 1 to Grade group 5. Due to the small sample sizes within each Grade group, Grade group 1 through Grade group 3 were categorized as one level as well as Grade group 4 through Grade group 5. A BCR was defined as either two successive post-RP PSAs of ≥0.2 ng/mL or the initiation of salvage therapy after a rising PSA of ≥0.1 ng/mL. A metastatic event was defined by a review of each patient’s radiographic scan history with a positive metastatic event defined as the date of a positive CT scan, bone scan, or MRI in their record. The associations between patient’s clinical characteristics and the outcomes of BCR and metastasis were analyzed by univariate Cox proportional hazards regression model, the associations of *CHD1* deletion and clinical outcomes with time to event outcomes, including BCR and metastasis, were analyzed by a Kaplan–Meier survival curves and tested using a log-rank test. Multivariable Cox proportional hazards models were used to estimated hazard ratios (HR) and 95% confidence intervals (Cis) after adjusting for age at diagnosis, PSA at diagnosis, race, pathological tumor stage, grade group, and surgical margins. We checked the proportional hazards assumption by plotting the log-log survival curves. A P-value < 0.05 was considered statistically significant. Analyses were performed in R version 4.0.2.

### Immunohistochemistry for ERG

ERG immunohistochemistry was performed as previously described^39^. Briefly, four μm TMA sections were dehydrated and blocked in 0.6% hydrogen peroxide in methanol for 20 min. and were processed for antigen retrieval in EDTA (pH 9.0) for 30 min in a microwave followed by 30 min of cooling in EDTA buffer. Sections were then blocked in 1% horse serum for 40 min and were incubated with the ERG-MAb mouse monoclonal antibody developed at CPDR (9FY, Biocare Medical Inc.) at a dilution of 1:1280 for 60 min at room temperature. Sections were incubated with the biotinylated horse anti-mouse antibody at a dilution of 1:200 (Vector Laboratories) for 30 min followed by treatment with the ABC Kit (Vector Laboratories) for 30 min. The color was developed by VIP (Vector Laboratories,) treatment for 5 minutes, and the sections were counter stained by hematoxylin. ERG expression was reported as positive or negative. ERG protein expression was correlated with clinico-pathologic features.

### Prostate cancer patients and specimens in the in-silico study cohorts Evaluation of the self-declared ancestries

Since the available ancestry data were based on the self-assessment of the patients, and it was a crucial part of our study to identify the samples accurately, we have interrogated the genotypes of 3000 SNPs that are specific to one of the greater Caucasian, African and Asian ancestries, in each of the germline samples ^40^. The data was collected into a single genotype matrix, the first two principal components of which was used to train a non-naïve Bayes classifier to differentiate between the three ancestries (details are available in the supplementary material, Supp. Figures 5-21).

### Identification of local subclonal loss of CHD1 in prostate adenocarcinoma

The paired germline and tumor binary alignment (bam) files were analyzed using bedtools genomcecov (v2.28.0)^41^, and their mean sequencing depths were determined. The coverage above and within the direct vicinity of CHD1 (*chr5:98,853,485-98,930,272 in grch38 and chr5:98,190,408-98,262,740 in grch37*) was collected in 50 bp wide bins into d-dimensional vectors (d_grch37 = 1447, d_grch38 = 1536) using an in-house tool and samtools (v1.6)^42^, and were normalized using their corresponding mean sequencing depths. The linear relationship between the paired germline-tumor coverages were determined in the following form:

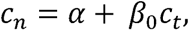

where *C_n_*is the normalized coverage of the germline sample and *C_t_* is the normalized coverage of its corresponding tumor pair. The intercept (*α*) was used to ensure that the data was free of outliers, and the slope (*β*_0_) was used as a raw measure of the

observable loss in the tumor. Similar slopes were calculated for 14 housekeeping genes in each of the sample-pairs, which were used to assess the significance of the loss (Supplementary Material).

The cellularity (c) of the tumors were estimated using sequenza^43^ after the rigorous selection of the most reliable cellularity-ploidy pair offered by the tool as alternative solutions. In order to account for the uncertainty of the reported cellularity values, a beta distribution was fitted on the grid-approximated marginal posterior densities of c. These were used to simulate random variables to determine the proportion of the approximate loss of CHD1 in the tumors, by the following formula:

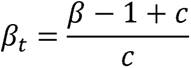

Here, *β*∼Normal(*β*_0_, *σ*), where *σ* is the standard error of *β*_0_, *c* ∼ Beta(*s*_1_, *s*_2_), where *s*_1_and *s*_2_ are the fitted shape-parameters of the cellularity, and *β*_t_ is the cellularity-adjusted slopes of the curve. The approximate level of loss in CHD1 is distributed as 1-*β*_0t_ (Further details are available in the supplementary materials, Suppl. Figures 23).

### Local subclonal LOH-calling

The SNP variant allele frequencies (VAF) in the close vicinity of *CHD1* in the tumor were collected with GATK HaplotypeCaller (v4.1.0) ^44^. The coverage and VAF data were carefully analyzed in order to ensure that we are strictly focusing on regions that have suffered the most serious loss (e.g., if only a part of the gene were lost, the unaffected region was excluded from the analysis). By using the tumor cellularity (c) and the estimated level of loss in the tumor (*β*_t_), we assessed whether a heterozygous or a homozygous subclonal deletion is more likely to result in the observed frequency pattern (A detailed explanation is available in the supplementary notes, Suppl. Figure 25, Suppl. Tables 2-3).

### Mutational signatures

Second generation somatic point-mutational signatures were estimated with the deconstructSigs R package^45^. The list of considered mutational processes whose signatures’ linear combination could lead to the final mutational catalogs (a.k.a. mutational spectra) were extracted in a dynamic process in which every single signature components were investigated one by one in an iterative manner and only those were kept that have improved the cosine similarity between the reconstructed and original spectra by a considerable margin (>0.001).

### HRD-scores

The calculation of the genomics scar scores (loss-of-heterozygosity: LOH, large-scale transitions: LST and number of telomeric allelic imbalances: ntAI) was performed using the scarHRD R package^46^. The allele-specific segmentation data of the samples were provided by sequenza^43^.

### Cell culture models

PC-3 and 22Rv1 prostate cell lines were purchased from ATCC^®^ and grown in RPMI 1640 (Gibco) supplemented with 10% FBS (Gibco) at 37°C in 5% CO_2,_ and regularly tested negative for Mycoplasma spp. contamination.

### Stable CRISPR-Cas9 expressing isogenic PC-3 cell line generation

Full length SpCas9 ORF was introduced in PC-3 cell population by Lentiviral transduction using lentiCas9-Blast (Addgene #52962) construction. After antibiotics (blasticidin) selection, survival populations were single cell cloned, isogenic cell lines were generated and tested for Cas9 activity by cleavage assay.

### Gene knock-out induction

CHD1 was targeted in CRISPR-Cas9 expressing PC-3 cell line using guide RNA CHD1_ex2_g1 (gCTGACTGCCTGATTCAGATC), resulted PC-3 CHD1 ko 1, and CHD1 ko 2 homozygous knock out cell lines. The same guide RNA was used to transiently knock out CHD1 gene in the 22Rv1 parental cell line.

### Transfection

Cells were transiently transfected by Nucleofector® 4D device (Lonza) by using supplemented, Nucleofector® SF solution and 20 μl Nucleocuvette® strips following the manufacturer’s instructions. Following transfection, cells were resuspended in 100 μl culturing media and plated in 1.5 ml pre-warmed culturing media in a 24 well tissue culture plate. Cells were subjected to further assays 72 h post transfection.

### In vitro T7 EndonucleaseI (T7E1) Assay

Templates used for T7E1 were amplified by PCR using CGTCAACGATGTCACTAGGC forward and ATGATTTGGGGCTTTCTGCT reverse oligos generating a 946 bp amplicon. 500 ng PCR products were denatured and reannealed in 1x NEBuffer 2.1 (New England Biolabs) using the following protocol: 95°C, 5 min; 95-85°C at -2°C/sec; 85-25°C at -0.1°C/sec; hold at 4°C. Hybridized PCR products were then treated with 10 U of T7E1 enzyme (New England Biolabs) for 30 min in a reaction volume of 30 μl. Reactions were stopped by adding 2 μl 0.5 M EDTA, fragments were visualized by agarose gel electrophoresis.

### Immunoblot Analysis

Freshly harvested cells were lysed in RIPA buffer. Protein concentrations were determined by Pierce BCA^TM^ Protein Assay Kit (Pierce). Proteins were separated via Mini Protean TGX stain free gel 4-15% (BioRad) and transferred to polyvinilydene difluoride membrane by using iBlot 2 PVDF Regular Stacks (Invitrogene) and iBlot system transfer system (LifeTechnologies).

Membranes were blocked in 5% BSA solution (Sigma). Primary antibodies were diluted following the manufacturer’s instructions: anti-Vinculin antibody (Cell Signaling) (1:1000) and antiCHD1 (Novus Biologicals) (1:2000).

Signals were developed by using Clarity Western ECL Substrate (BioRad) and Image Quant LAS4000 System (GEHealthCare).

### Proximity Ligation Assay (PLA)

Cells were seeded in μ-slide 8 well chambers (Ibidi GmbH, Germany) and incubated overnight. Next day, cells were subjected to irradiation (4Gy). Irradiated and control cells (0Gy) were recovered for 3hrs, then fixed with 4% PFA and permeabilized with 0.3% Triton X-100.

Duolink® Proximity Ligation Assay (Sigma) was carried out using antibodies against γH2Ax and RAD51(Cell Signaling) according to the manufacturer’s instruction. Signals were detected by fluorescent microscopy (Nikon Ti2-e Live Cell Imaging System). Quantification of fluorescent signals were carried out by using the Fiji-ImageJ software.

### Sample preparation for Whole Genome Sequencing (WGS)

DNA was extracted from 22Rv1 and PC-3 *CHD1* knock out isogenic cell lines at low passage number of the cells (22Rv1_1, PC-3_1). Following 45 passages, CHD1 knock out isogenic cell line was single cell cloned, and two colonies per cell line (22Rv1_2, 22Rv1_3, PC-3_2, PC-3_3) were propagated for DNA isolation.

DNA was extracted by using QIAamp DNA Mini Kit (QIAGENE). Whole Genome Sequencing of the DNA samples was carried out at Novogene service company.

### Viability cell proliferation assays

Exponentially growing PC-3 cell lines WT, *CHD1* ko1, *CHD1* ko2, and 22Rv1 WT and chd1 ko respectively, were seeded in 96-well plates (1500 PC-3 cells/well, and 3000 22 Rv1 cells/well) and incubated for 36 hrs to allow cell attachment. Identical cell numbers of seeded paralel isogenic lines were verified by the Celigo Imaging Cytometer after attachment. Cells were exposed to Talazoparib (Selleckchem), Olaparib (MedChemExpress) and Bleomycin sulfate (Fisher Scientific) for 24 hrs, then kept in drug-free fresh media for 5 days until cell growth was determined by the addition of PrestoBlue^TM^ (Invitrogen) and incubated for 2.5 hrs. Cell viability was determined by using the BioTek plate reader system. Fluorescence was recorded at 560 nm/590 nm, and values were calculated based on the fluorescence intensity. IC50 values were determined by using the AAT Bioquest IC50 calculator tool. P-values were calculated using Student’s t-test. P-values <0.05 were considered statistically significant.

### NGS analysis of the PC-3 and 22Rv1 whole genomes sequences

The reads of the six WGS (3 PC-3 and 3 22Rv1) were aligned to the grch37 reference genome using the bwa-mem^47^ aligner. The resulting bam files were post-processed according to the GATK best-practices guidelines. Novel variants were called using Mutect2 (v4.1.0) by using CHD1 intact WGS references downloaded from the Sequence Read Archive (SRA, with accession IDs; PC-3: SRX5466646, 22Rv1: SRX5437595) as “normal” and the *CHD1* ko clones as “tumor” specimens^44^. These vcfs were converted into tab-delimited files and further analyzed in R. Annotation was performed via Intervar^48^ .

## Acknowledgement

The authors thank Zita Bratu for technical assistance, Alimamy Bundu and Treissy Soares for FISH probe preparation and testing, Dr. Hua Zou, Audrey Flores and Safaa Khairi for valuable experimental support and Orsolya Pipek for the technical support. This work was supported by the Research and Technology Innovation Fund (KTIA_NAP_13-2014-0021 and NAP2-2017-1.2.1-NKP-0002); Breast Cancer Research Foundation (BCRF-21-159 to Z. Szallasi) and the Novo Nordisk Foundation Interdisciplinary Synergy Program Grant (NNF15OC0016584), Det Fri Forskningsrad (award number #7016-00345B; to Z. Szallasi); Department of Defense through the Prostate Cancer Research Program (award number is W81XWH-18-2-0056; to Z. Szallasi, A. Dobi and M.L. Freedman); and the National Cancer Institute (P01CA228696, to A. D’Andrea, Z. Szallasi, M.L. Freedman). Z. Szallasi, Z. Sztupinszki and J. Borcsok were supported by Velux Foundation 00018310 grant.

S.K. is supported by the Prostate Cancer Foundation (18YOUN09 and 19CHAL07).

## Disclaimer

The contents of this publication are the sole responsibility of the author(s) and do not necessarily reflect the views, opinions or policies opinions of Uniformed Services University of the Health Sciences (USUHS), the Henry M. Jackson Foundation for the Advancement of Military Medicine, Inc., the Department of Defense (DoD) or the Departments of the Army, Navy, or Air Force. Mention of trade names, commercial products, or organizations does not imply endorsement by the U.S. Government.

## Author Contributions

*Conception and design:* Miklos Diossy, V. Tisza, H. Li, J. Zhou, Zs. Sztupinszki, S. Spisak, G. Valcz, P. V. Nuzzo, D. Ribli, T. Ried, S. Kaochar, S. Pathania, A. D’Andrea, I. Csabai, S. Srivastava, A. Dobi, M. L. Freedman, Z. Szallasi

*Development of methodology*: M. Diossy, V. Tisza, H. Li, J. Zhou, Zs. Sztupinszki, M. Krzystanek, A. Dobi, Z. Szallasi

*TMA analysis:* H. Li, D. Young, D. Nousome, C. Kuo, Y. Chen, R. Ebner, I. A. Sesterhenn, Gy. Petrovics, G. Valcz

*Acquisition of data* : M. Diossy, V. Tisza, H. Li, J. Zhou, Zs. Sztupinszki, D. Young, D. Nousome, C. Kuo, J. Jiang, G. Valcz, D. Ribli

*Cell line experiments* : M. Diossy, V. Tisza, J. Zhou, G. T. Klus, S. Spisak, T. Ried, Z. Szallasi

*Analysis and interpretation of data (e.g., statistical analysis, biostatistics, computational analysis)* : M. Diossy, V. Tisza, H. Li, Zs. Sztupinszki, D. Nousome, A. Schina, J. Börcsök, A. Prosz, I. Csabai, S. Srivastava, M. L. Freedman, Z. Szallasi

*Administrative, technical, or material support (i.e., reporting or organizing data, constructing databases)* : M. Diossy, V. Tisza, H. Li, J. T. Moncur, G. T. Chesnut, S. Srivastava, A. Dobi, Z. Szallasi

*Study supervision* : A. Dobi, S Spisak, Z. Szallasi

All authors were involved in the preparation of the manuscript and the supplementary materials.

## Data Availability

Whole exome and whole genome TCGA data presented in this study are available from the GDC (https://portal.gdc.cancer.gov/) and ICGC (https://dcc.icgc.org/) data portals respectively. The whole genomes from the Mayo clinic are available from dbGap (phs001105.v1.p1), while whole genomes from DFCI and CPDR are available upon request.

## Code availability

All analysis was done using standard R (v4.1) codes with the help of the following packages: ggplot2, data.table, deconstructSigs, lsa, ggbeeswarm, RColorBrewer, sequenza, copy number, and cluster. In particular, standard variant files were converted to tab-delimited tables using GATK (v3.8) VariantsToTable and manipulated using the data.table package in R. Figures were created using ggplot2. Every tool mentioned in the Methods section were used with default parameters unless stated otherwise.

## Ethical statement

Patients in this retrospective cohort were treated with radical prostatectomy (RP) at the Walter Reed National Military Medical Center, donated specimens under the approved protocol #393738 and follow up data under the approved protocol #GT90CM Uniformed Services University of the Health Sciences. The study proposal was reviewed by the Uniformed Services University’s Human Research Protections Program (HRPP) Office and “determined to be considered research not involving human subjects as defined by 32 CFR 219.102(e) because the research involves the use of de-identified specimens and data not collected specifically for this study.” (Ref #910230).

## Notes

### Competing Interest Statement

The authors have declared no competing interest.

### Summary of Updates

In this revision, the article underwent substantial revisions, resulting in significant improvements and updates. The core content of the article was extensively reworked to enhance its overall clarity, coherence, and depth. Notably, the figures presented within the article were carefully revised to better illustrate the concepts discussed and to align more closely with the updated textual content. Additionally, the supplementary material accompanying the article was thoroughly updated, incorporating the latest findings and ensuring a comprehensive and cohesive presentation of the research.

