## Supplementary Notes for "Subclonal *CHD1* deletion is more frequent in African American prostate cancers and associated with rapid disease progression and limited levels of homologous recombination deficiency"

Supplementary Material

*Authors:*

Miklos Diossy, Viktoria Tisza, Hua Li, Jia Zhou, Zsofia Sztupinszki, Denise Young, Darryl Nousome, Claire Kuo, Jiji Jiang, Yongmei Chen, Reinhard Ebner, Isabell A. Sesterhenn, Joel T. Moncur, Gregory T. Chesnut, Gyorgy Petrovics, Gregory T. Klus, Sandor Spisak, Gabor Valcz, Pier Vitale Nuzzo, Dezso Ribli, Aimilia Schina, Judit Börcsök, Aurel Prosz, Marcin Krzystanek, Thomas Ried, David Szuts, Salma Kaochar, Shailja Pathania, Alan D'Andrea, Istvan Csabai, Shiv Srivastava, Albert Dobi, Matthew L Freedman, Zoltan Szallasi

*Date of the latest update:*
November 17, 2022

**5 Determining the sub-clonal loss of *CHD1*** **25**

**6 *CHD1* loss summary - NGS samples** **27**

**7 Genotyping** **28**

**8 Assessing the local loss of heterozygosity** **29**

### List of Supplementary Tables

1 Supplementary tables of the TMA analysis . . . . . . . . . . . . . . . . . . . . . . . . . . . . . . . 3

### Supplementary tables not included in this notebook

- **Supplementary Table 2**: Subclonal *CHD1* loss prediction in the WGS cohorts (available separetely in xlsx format)
- **Supplementary Table 3**: Subclonal *CHD1* loss prediction in the WES cohorts (available separately in xlsx format)

### 1 TCGA SNP Array Data

The Affymetrix SNP Array 6.0 data of 495 TCGA patients were downloaded from the TCGA legacy archive data portal: https://portal.gdc.cancer.gov/legacy-archive, and the raw CEL files were preprocessed using the AROMA affymetrix R package. Raw copy number estimates, i.e. piecewise constant segments were fitted (PCF) and B-allele frequencies (BAF) were collected by following the guidelines of the CRMAv2 pipeline:
(https://aroma-project.org/vignettes/CRMAv2/).

The BAFs of the patients' probes were collected into a single **G** matrix, the first five eigenvalues and eigenvectors (i.e. principle components) of which were subsequently calculated. We have found, that PC2 and PC3 separated the samples according to their ancestries, and by using patients with self-declared racial data as templates, the ancestries of non-declared samples were identified using the density-based spatial clustering of applications with noise (DBSCAN) [1] clustering algorithm (Suppl.Fig.1):
https://scikit-learn.org/stable/auto_examples/cluster/plot_dbscan.html.

Patients who were classified as outliers were excluded from the PCF-part of the analysis. DBSCAN was able to identify 251 Caucasian (CA) and 46 African American (AA) patients.

In our initial investigation the piecewise constant fitted raw copy numbers were averaged across the Caucasian and African American patient groups, and the entire genome was surveyed for clearly observable differences between the two. A substantial depletion was found around the centromere of chromosome 5 in both ancestry groups, along with an even more severe loss in AA patients, which was centered around *CHD1* (Suppl.Fig.2). This preliminary analysis have found that 9/46 (19.6%) AA patients and 18/251 (7.2%) CA patients had a deep loss in the vicinity of *CHD1*. This observation has fueled our entire investigation.

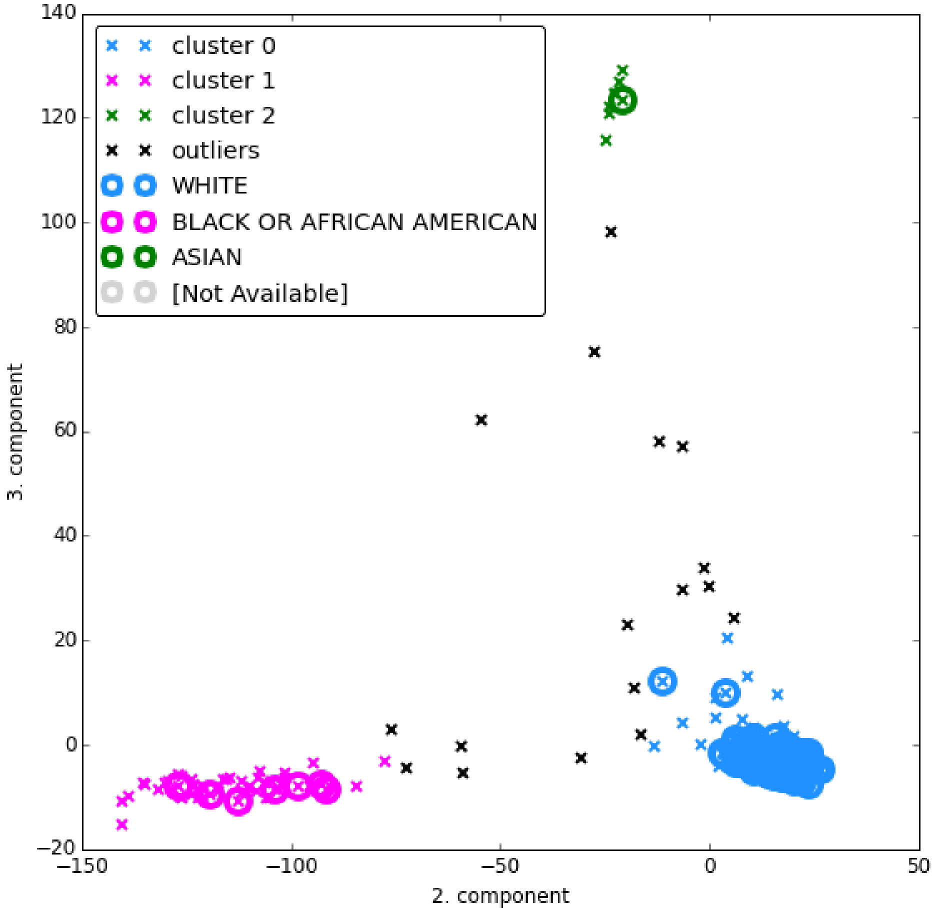

**Suppl.Fig. 1:** Non-declared ancestry classification on SNP-array data. Circles indicate samples with self declared ancestry, while colored crosses indicate the density-estimated clusters, according to the DBSCAN classification algorithm.

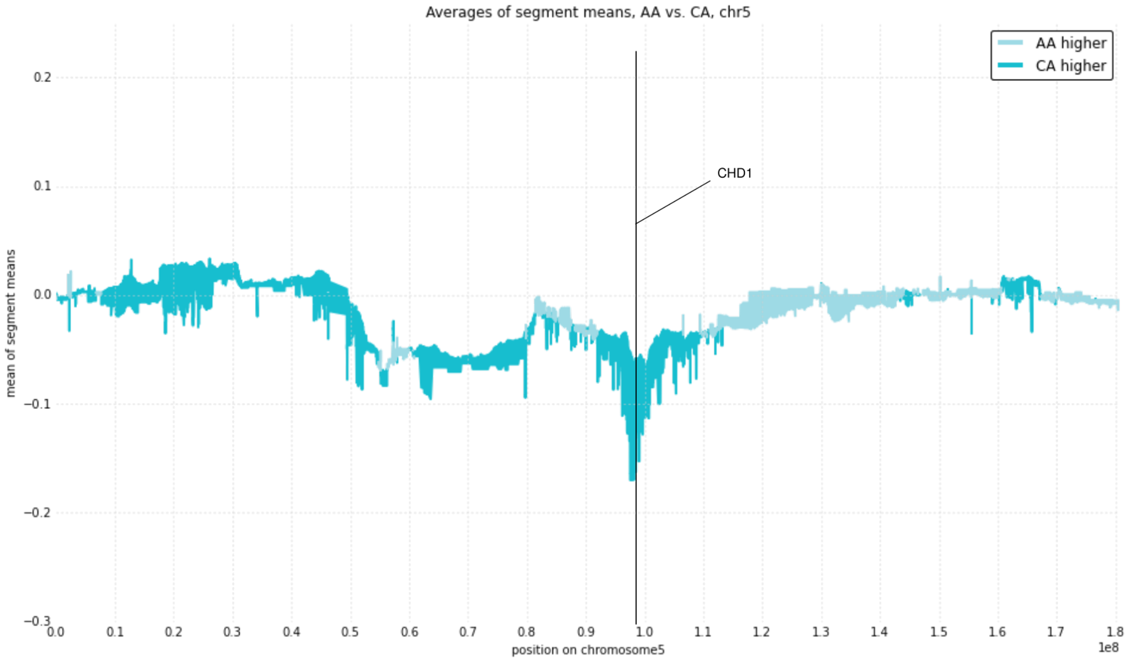

**Suppl.Fig. 2:** The average raw copynumbers of Caucasian (CA) and African American (AA) individuals on chromosome 5. The plotting style is fill between the lines. Darker shades indicate regions where the average copynumber of CA patients was higher, lighter color correspond to regions where AA mean segments were higher. A vertical line indicates the location of *CHD1* on the chromosome. The fitted segment mean of the African American samples ($\langle s_{AA} \rangle$ = -0.057, 95% CI: [-0.059, -0.055] ) was almost three times lower than of the fitted mean of the European Americans ($\langle s_{CA} \rangle$ = -0.167, 95% CI: [-0.176, -0.158])

### 2 TMA

**a**

| | African American (N=91) | European American (N=109) | P value |
|---|---|---|---|
| **Age at Dignosis** | | | 0.020 |
| Mean (SD) | 59.7 (6.6) | 61.9 (6.2) | |
| Range | 44.8-73.2 | 45.7-74.6 | |
| **PSA at Diagnosis (ng/ml)** | | | 0.435 |
| <4.0 | 7 (8.1%) | 16 (15.5%) | |
| 4.0-9.0 | 54 (62.8%) | 63 (61.2%) | |
| 10.0-20.0 | 21 (24.4%) | 20 (19.4%) | |
| >20.0 | 4 (4.7%) | 4 (3.9%) | |
| **Pathological T Stage** | | | 0.631 |
| pT2 | 44 (48.4%) | 49 (45.0%) | |
| pT3-4 | 47 (51.6%) | 60 (55.0%) | |
| **Pathological Gleason Score** | | | 0.601 |
| 3+3 | 33 (36.2%) | 41 (37.7%) | |
| 3+4 | 29 (31.9%) | 35 (32.1%) | |
| 4+3 | 10 (11.0%) | 7 (6.4%) | |
| 8 to 10 | 16 (17.6%) | 18 (16.5%) | |
| Treatment | 3 (3.3%) | 8 (7.3%) | |
| **Grade Group** | | | 0.406 |
| GG1-GG2 | 62 (68.1%) | 76 (69.7%) | |
| GG3 | 10 (11.0%) | 7 (6.4%) | |
| GG4-GG5 | 16 (17.6%) | 18 (16.5%) | |
| Treatment | 3 (3.3%) | 8 (7.3%) | |
| **Margin Status** | | | 0.297 |
| Negative | 51 (56.0%) | 69 (63.3%) | |
| Positive | 40 (44.0%) | 40 (36.7%) | |
| **BCR** | | | 0.324 |
| No | 58 (63.7%) | 62 (56.9%) | |
| Yes | 33 (36.3%) | 32 (29.4%) | |
| **Metastasis** | | | 0.440 |
| No | 81 (89.0%) | 93 (85.3%) | |
| Yes | 10 (11.0%) | 16 (14.7%) | |

**b**

| Patient | TMA Block | TMA Row | CORE a | CORE b | CORE c | CORE d | CORE e | CORE f | CORE g | CORE h | CORE i | CORE j | CORE k | CORE l |
|---|---|---|---|---|---|---|---|---|---|---|---|---|---|---|
| 1 | 1 | 1 | Normal | Normal | PIN | PIN | T1A | T1A | T2A | T2A | T3A | T3A | T4A | T4A |
| 2 | 1 | 2 | Normal | Normal | PIN | PIN | T1A | T1A | T1B | T2A | T2A | T3A | | |
| 3 | 1 | 3 | Normal | Normal | T1A | T1A | T1B | T1B | | | | | | |
| 4 | 1 | 4 | Normal | Normal | PIN | PIN | T1A | T1A | T1B | T1B | T2A | T2A | | |
| 5 | 1 | 5 | Normal | Normal | PIN | PIN | T1A | T1A | T1B | T1B | | | | |
| 6 | 1 | 6 | Normal | Normal | PIN | PIN | T1A | T1A | T1B | T1B | T1C | T1C | | |
| 7 | 1 | 7 | Normal | Normal | T1A | T1A | T2A | T2A | T4A | T4A | T7A | T7A | | |
| 8 | 1 | 8 | Normal | Normal | T1A | T1A | T1B | T1B | T2A | T2A | T2B | T2B | | |
| 9 | 1 | 9 | Normal | Normal | PIN | PIN | T1A | T1A | T2A | T2A | T3A | T3A | T4A | T4A |

**c**

| Race | Patients | Focal Tumors | Tumor Cores |
|---|---|---|---|
| **AA (African American)** | 91 | 162 | 385 |
| **EA (European American)** | 109 | 189 | 478 |

**d**

| | *CHD1* Deletion (N=41) | No *CHD1* Deletion (N=159) | P value |
|---|---|---|---|
| **Pathologic T Stage** | | | 0.043 |
| pT2 | 13 (31.7%) | 81 (50.9%) | |
| pT3-pT4 | 28 (68.3%) | 78 (49.1%) | |
| **Pathologic Gleason Score** | | | <0.001 |
| 3+3 | 3 (7.3%) | 70 (44.0%) | |
| 3+4 | 17 (41.5%) | 47 (29.6%) | |
| 4+3 | 7 (17.1%) | 10 (6.3%) | |
| 8 to 10 | 11 (26.8%) | 25 (15.7%) | |
| Treatment | 3 (7.3%) | 7 (4.4%) | |
| **Grade Group** | | | 0.024 |
| GG1-GG2 | 20 (48.8%) | 117 (7.36%) | |
| GG3 | 7 (17.1%) | 10 (6.3%) | |
| GG4-GG5 | 11 (26.8%) | 25 (15.7%) | |
| Treatment | 3 (7.3%) | 7 (4.4%) | |

**e**

| Gene Defects | AA (N=42) | EA (N=59) | P value |
|---|---|---|---|
| **PTEN status** | | | <0.0001 |
| Deletion | 8 (19.0%) | 38 (64.4%) | |
| No-deletion | 34 (81.0%) | 21 (35.6%) | |
| **ERG status** | | | 0.000525 |
| Positive | 9 (21.4%) | 33 (55.9%) | |
| Negative | 33 (78.6%) | 26 (44.1%) | |

**Suppl.Tab. 1:** **a**: Distributions of clinico-pathological features in combined prostate cancer cohort (N=200).; **b**: The grid map of a representative TMA block construction (PIN=prostatic intraepithelial neoplasia, T1= index tumor; T2=secondary tumor and so on) **c**: Multiple tumor samples of 91 AA and 109 EA patients derived tumor TMA; **d**: *CHD1* deletion frequency correlations with pathological T-stage, pathological Gleason score and Grade group in prostate cancer patients (N=200) **e**: Frequency of PTEN deletion and expression of ERG protein in a subset of the cohort (N=101, AA=42 and CA=59).

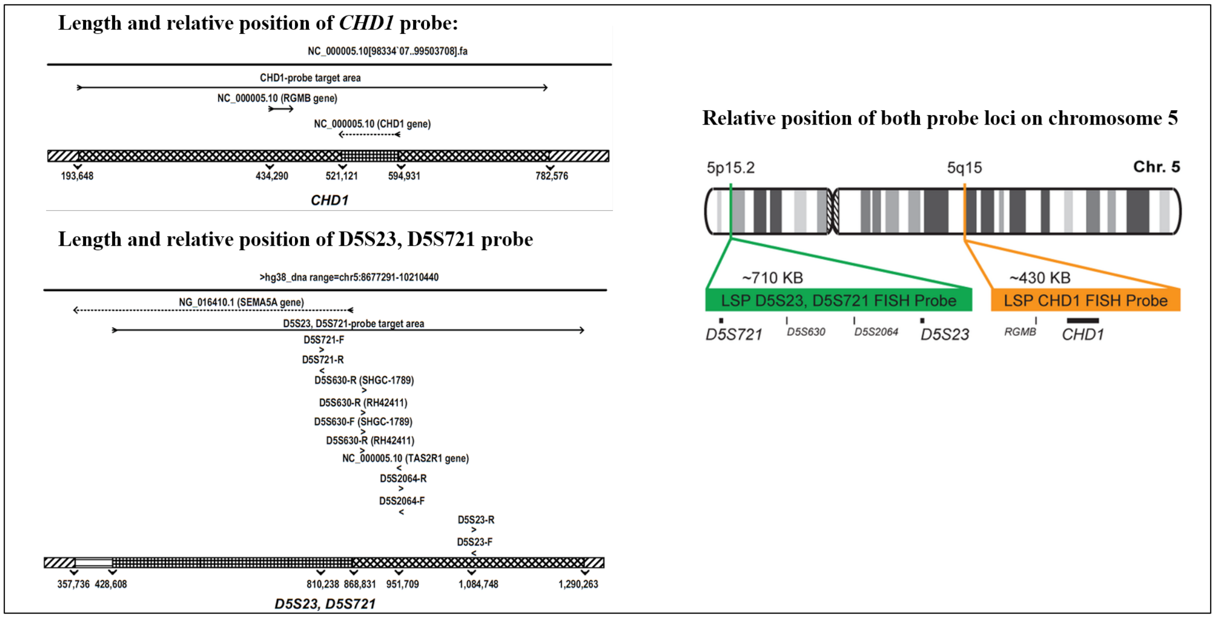

**Suppl.Fig. 3:** The design strategy of FISH probes for *CHD1* and chromosome 5 short arm.

**Suppl.Fig. 4: a:** The discordance map of *CHD1* deletion vs. PTEN deletion and ERG expression in TMA cores carrying *CHD1* deletion (purple, AA cases 1 to 12 and 6 EA cases 43-48). Samples from benign tissue are marked in CORE a and b. T1-IN indicates index, T2-NIN marks secondary tumors and so on, in TMA cores c to i. Patient codes are shown in the "Case" column **b:** The discordance map of *CHD1* deletion, PTEN deletion and ERG expression at patient level. Patient codes are shown in the "Case" column.

**a**

| | Case | Race | CORE a | CORE b | CORE c | CORE d | CORE e | CORE f | CORE g | CORE h | CORE i | CORE j | CORE k | CORE i | BCR | Met |
|---|---|---|---|---|---|---|---|---|---|---|---|---|---|---|---|---|
| PTEN | 1 | AA | | | | | T1-IN | T1-IN | | | T2-NIN | T2-NIN | | | Y | N |
| ERG | 1 | AA | | | | | T1-IN | T1-IN | | | T2-NIN | T2-NIN | | | Y | N |
| CHD1 | 1 | AA | | | | | T1-IN | T1-IN | | | | | | | Y | N |
| PTEN | 2 | AA | | | | | | T1-IN | | | | | | | Y | N |
| ERG | 2 | AA | | | | | | | | | | | | | Y | N |
| CHD1 | 2 | AA | | | | | T1-IN | | T2-NIN | | | | | | Y | N |
| PTEN | 3 | AA | | | | | T1-IN | T1-IN | | | | | | | Y | N |
| ERG | 3 | AA | | | | | | | | | | | | | Y | N |
| CHD1 | 3 | AA | | | | | T1-IN | T1-IN | | | T2-NIN | T2-NIN | T2-NIN | | Y | N |
| PTEN | 4 | AA | | | | | T1-IN | T1-IN | T2-NIN | | | | | | Y | N |
| ERG | 4 | AA | | | | | | | | | | | | | Y | N |
| CHD1 | 4 | AA | | | | | | | | | T3-NIN | | | | Y | N |
| PTEN | 5 | AA | | | | | | | | | | | | | Y | Y |
| ERG | 5 | AA | | | | | | | T1-IN | | | | | | Y | Y |
| CHD1 | 5 | AA | | | | | | | T1-IN | T1-IN | | | | | Y | Y |
| PTEN | 6 | AA | | | | | | | | | | | | | Y | Y |
| ERG | 6 | AA | | | | | | | | | | | | | Y | Y |
| CHD1 | 6 | AA | | | | | | | T1-IN | T1-IN | | | | | Y | Y |
| PTEN | 7 | AA | | | | | | | | | | | | | Y | N |
| ERG | 7 | AA | | | | | | | | | | | | | Y | N |
| CHD1 | 7 | AA | | | | T1-IN | | | | | | T1-IN | | | Y | N |
| PTEN | 8 | AA | | | | | | | | | | | | | Y | N |
| ERG | 8 | AA | | | | | | | | | | | | | Y | N |
| CHD1 | 8 | AA | | | | | | | | T2-NIN | | | | | Y | N |
| PTEN | 9 | AA | | | | | | | | | | | | | Y | N |
| ERG | 9 | AA | | | | | | | | | | | | | Y | N |
| CHD1 | 9 | AA | | | | | T1-IN | T1-IN | | | | | | | Y | N |
| PTEN | 10 | AA | | | | | | | | | | | | | Y | N |
| ERG | 10 | AA | | | | | | | | | | | | | Y | N |
| CHD1 | 10 | AA | | | | | | | T1-IN | | | | | | Y | N |
| PTEN | 11 | AA | | | | | | | | | | | | | Y | N |
| ERG | 11 | AA | | | | | | | | | | | | | N | N |
| CHD1 | 11 | AA | | | | | | T1-IN | T1-IN | T1-IN | | | | | N | N |
| PTEN | 12 | AA | | | | | | | | | | | | | N | N |
| ERG | 12 | AA | | | | | | | | | | | | | N | N |
| CHD1 | 12 | AA | | | | | | | T1-IN | T1-IN | | | | | N | N |

| | Case | Race | CORE a | CORE b | CORE c | CORE d | CORE e | CORE f | CORE g | CORE h | CORE i | CORE j | CORE k | CORE i | BCR | Met |
|---|---|---|---|---|---|---|---|---|---|---|---|---|---|---|---|---|
| PTEN | 43 | EA | | | | T1-IN | T1-IN | T1-IN | T1-IN | T1-IN | T2-NIN | T2-NIN | | | Y | Y |
| ERG | 43 | EA | | | | T1-IN | T1-IN | T1-IN | T1-IN | T1-IN | T2-NIN | T2-NIN | | | Y | Y |
| CHD1 | 43 | EA | | | | | T1-IN | | | | | | | | Y | Y |
| PTEN | 44 | EA | | | T1-IN | T1-IN | | T1-IN | | T1-IN | | | | | Y | Y |
| ERG | 44 | EA | | | T1-IN | T1-IN | T1-IN | T1-IN | T1-IN | T1-IN | | | | | Y | Y |
| CHD1 | 44 | EA | | | | | | T1-IN | T1-IN | | | | | | Y | Y |
| PTEN | 45 | EA | | | | | T1-IN | T1-IN | | T1-IN | | | | | Y | N |
| ERG | 45 | EA | | | | | T1-IN | T1-IN | | | | | | | Y | N |
| CHD1 | 45 | EA | | | | | | T1-IN | | T1-IN | | | | | Y | N |
| PTEN | 46 | EA | | | | | | | T2-NIN | | | | | | N | N |
| ERG | 46 | EA | | | | T1-IN | | | | | | | | | N | N |
| CHD1 | 46 | EA | | | | | | | T2-NIN | T2-NIN | | | | | N | N |
| PTEN | 47 | EA | | | | | T1-IN | T1-IN | T1-IN | T1-IN | T2-NIN | | | | Y | N |
| ERG | 47 | EA | | | | | | | | | | | | | Y | N |
| CHD1 | 47 | EA | | | | | | | T1-IN | | | | | | Y | N |
| PTEN | 48 | EA | | | | | | | | | | | | | N | N |
| ERG | 48 | EA | | | | | | | | | | | | | N | N |
| CHD1 | 48 | EA | | | T1-IN | | T1-IN | T1-IN | T1-IN | | | | | | N | N |

| | |
|---|---|
| | *PTEN* deletion |
| | ERG expression |
| | *CHD1* deletion |
| | No defect |
| | Benign |
| T1-IN | Index Tumor |
| T2-NIN | Secondary Tumor |
| T3-NIN | Tertial Tumor |
| Y | Yes, BCR |
| Y | Yes, Met |
| N | No BCR/Met |

**b**

| Case | Race | CHD1 | PTEN | ERG | BCR | Met |
|---|---|---|---|---|---|---|
| 1 | AA | | | | Y | N |
| 2 | AA | | | | Y | N |
| 3 | AA | | | | Y | N |
| 4 | AA | | | | Y | N |
| 5 | AA | | | | Y | Y |
| 6 | AA | | | | Y | Y |
| 7 | AA | | | | Y | N |
| 8 | AA | | | | Y | N |
| 9 | AA | | | | Y | N |
| 10 | AA | | | | Y | N |
| 11 | AA | | | | N | N |
| 12 | AA | | | | N | N |
| 13 | AA | | | | Y | N |
| 14 | AA | | | | N | N |
| 15 | AA | | | | Y | N |
| 16 | AA | | | | N | N |
| 17 | AA | | | | Y | Y |
| 18 | AA | | | | Y | Y |
| 19 | AA | | | | Y | N |
| 20 | AA | | | | Y | N |
| 21 | AA | | | | Y | N |
| 22 | AA | | | | Y | Y |
| 23 | AA | | | | Y | Y |
| 24 | AA | | | | Y | N |
| 25 | AA | | | | Y | N |
| 26 | AA | | | | Y | N |
| 27 | AA | | | | Y | N |
| 28 | AA | | | | Y | N |
| 29 | AA | | | | N | N |
| 30 | AA | | | | N | N |
| 31 | AA | | | | N | N |
| 32 | AA | | | | N | N |
| 33 | AA | | | | N | N |
| 34 | AA | | | | N | N |
| 35 | AA | | | | N | N |
| 36 | AA | | | | N | N |
| 37 | AA | | | | N | N |
| 38 | AA | | | | N | N |
| 39 | AA | | | | N | N |
| 40 | AA | | | | N | N |
| 41 | AA | | | | N | N |
| 42 | AA | | | | N | N |

| Case | Race | CHD1 | PTEN | ERG | BCR | Met |
|---|---|---|---|---|---|---|
| 43 | EA | | | | Y | Y |
| 44 | EA | | | | Y | Y |
| 45 | EA | | | | Y | N |
| 46 | EA | | | | N | N |
| 47 | EA | | | | Y | N |
| 48 | EA | | | | N | N |
| 49 | EA | | | | Y | Y |
| 50 | EA | | | | Y | Y |
| 51 | EA | | | | Y | Y |
| 52 | EA | | | | Y | Y |
| 53 | EA | | | | Y | N |
| 54 | EA | | | | Y | N |
| 55 | EA | | | | Y | N |
| 56 | EA | | | | Y | N |
| 57 | EA | | | | Y | N |
| 58 | EA | | | | N | N |
| 59 | EA | | | | N | N |
| 60 | EA | | | | N | N |
| 61 | EA | | | | N | N |
| 62 | EA | | | | N | N |
| 63 | EA | | | | N | N |
| 64 | EA | | | | N | N |
| 65 | EA | | | | N | N |
| 66 | EA | | | | N | N |
| 67 | EA | | | | N | N |
| 68 | EA | | | | Y | N |
| 69 | EA | | | | Y | N |
| 70 | EA | | | | Y | N |
| 71 | EA | | | | Y | N |
| 72 | EA | | | | Y | N |
| 73 | EA | | | | Y | N |
| 74 | EA | | | | Y | N |
| 75 | EA | | | | Y | N |
| 76 | EA | | | | Y | N |
| 77 | EA | | | | N | Y |
| 78 | EA | | | | N | N |
| 79 | EA | | | | N | N |
| 80 | EA | | | | N | N |
| 81 | EA | | | | N | N |
| 82 | EA | | | | Y | N |
| 83 | EA | | | | Y | N |
| 84 | EA | | | | N | N |
| 85 | EA | | | | N | N |
| 86 | EA | | | | N | N |
| 87 | EA | | | | N | N |
| 88 | EA | | | | N | N |
| 89 | EA | | | | N | N |
| 90 | EA | | | | N | N |
| 91 | EA | | | | N | N |
| 92 | EA | | | | Y | Y |
| 93 | EA | | | | Y | N |
| 94 | EA | | | | Y | N |
| 95 | EA | | | | Y | N |
| 96 | EA | | | | Y | N |
| 97 | EA | | | | Y | N |
| 98 | EA | | | | Y | N |
| 99 | EA | | | | N | N |
| 100 | EA | | | | N | N |
| 101 | EA | | | | N | N |

| | |
|---|---|
| | ***PTEN* deletion** |
| | **ERG expression** |
| | ***CHD1* deletion** |
| | **No defect** |
| | **Benign** |
| **Y** | **Yes, BCR** |
| **Y** | **Yes, Met** |
| **N** | **No BCR/Met** |

**Suppl.Fig. 5:**
**a:** The association between *CHD1* deletion and BCR in all (AA and EA, left) and EA (right) only prostate cancer patients with Kaplan-Meier curve analysis.
**b:** Univariable survival analysis of the clinical features associated with BCR (N=189, excluding 11 patients receiving neo-adjuvant therapy).
**c:** Univariable survival analysis of the clinical features associated with metastasis (N=189, excluding 11 patients receiving neo-adjuvant therapy).
**d:** The Association between *CHD1* deletion and BCR in all (AA and EA) prostate cancer patients with multivariable Cox model analysis adjusting for other key factors associated with BCR (top table with Gleason Group, bottom table with pathological Gleason score).
**e:** The association between *CHD1* deletion and metastasis in all (AA and EA, left) and EA (right) only prostate cancer patients with Kaplan-Meier curve analysis.
**f:** The association between *CHD1* deletion and metastasis in all (AA and EA) prostate cancer patients with multivariable Cox model analysis adjusting for other key factors associated with BCR (top table with Gleason Group, bottom table with pathological Gleason score).

**a**

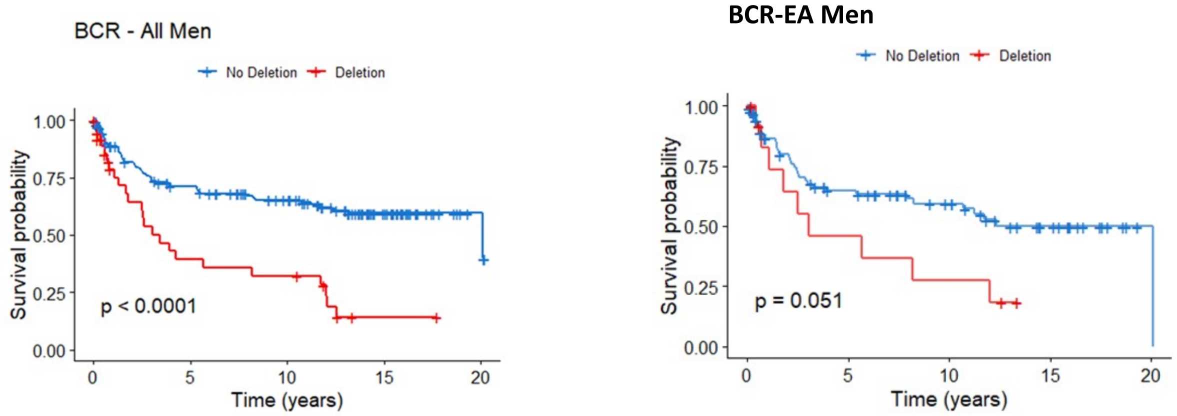

**b**

| | | | | time_RP_BCR | | |
|---|---|---|---|---|---|---|
| Variables | Level | N | Hazard Ratio (95% CI) | HR P-value | Assumption P-value | Log-rank P-value |
| ***CHD1* Deletion** | Deletion | 37 | 2.80 (1.72-4.57) | **<.001** | 0.193 | **<.001** |
| | No Deletion | 152 | - | - | | |
| RACE | European American | 101 | 1.38 (0.88-2.19) | 0.164 | 0.285 | 0.162 |
| | African American | 88 | - | - | | |
| Path T Stage | pT3-pT4 | 104 | 5.58 (3.23-9.62) | **<.001** | 0.218 | **<.001** |
| | pT2 | 85 | - | - | | |
| GG2_cat | GG4-GG5 | 40 | 2.30 (1.38-3.83) | **0.001** | 0.421 | **<.001** |
| | GG1-GG3 | 149 | - | - | | |
| Path Gleason | From 8 to 10 | 34 | 3.83 (1.92-7.66) | **<.001** | 0.449 | **<.001** |
| | 3+4 | 64 | 2.41 (1.35-4.30) | **0.003** | | |
| | 4+3 | 17 | 4.86 (2.29-10.34) | **<.001** | | |
| | 3+3 | 74 | - | - | | |
| Surgical margin | Positive | 76 | 3.80 (2.39-6.04) | **<.001** | 0.313 | **<.001** |
| | Negative | 113 | - | - | | |
| Adjuvant therapy | Yes | 47 | 10.46 (6.10-17.95) | **<.001** | 0.002 | **<.001** |
| | No | 142 | - | - | | |
| Age at Diagnosis | | 189 | 1.03 (0.99-1.06) | 0.142 | 0.154 | - |
| PSA at Diagnosis | | 178 | 1.42 (0.99-2.03) | 0.059 | 0.433 | - |

**c**

| | | | | time_Dx_mets | | |
|---|---|---|---|---|---|---|
| **Variables** | **Level** | **N** | **Hazard Ratio (95% CI)** | **HR P-value** | **Assumption P-value** | **Log-rank P-value** |
| **CHD1 Deletion** | **Deletion** | 37 | 2.99 (1.33-6.75) | **0.008** | 0.466 | **0.005** |
| | **No Deletion** | 152 | - | - | | |
| **RACE** | **European American** | 101 | 1.45 (0.65-3.22) | 0.367 | 0.199 | 0.364 |
| | **African American** | 88 | - | - | | |
| **Path T Stage** | **pT3-pT4** | 104 | 4.48 (1.53-13.05) | **0.006** | 0.405 | **0.003** |
| | **pT2** | 85 | - | - | | |
| **GG2_cat** | **GG4-GG5** | 40 | 3.04 (1.36-6.82) | **0.007** | 0.252 | **0.005** |
| | **GG1-GG3** | 149 | - | - | | |
| **Path Gleason** | **From 8 to 10** | 34 | 5.30 (1.83-15.34) | **0.002** | 0.286 | **0.002** |
| | **3+4** | 64 | 1.40 (0.44-4.40) | 0.57 | | |
| | **4+3** | 17 | 1.90 (0.37-9.85) | 0.443 | | |
| | **3+3** | 74 | - | - | | |
| **Surgical margin** | **Positive** | 76 | 2.76 (1.22-6.26) | **0.015** | 0.375 | **0.011** |
| | **Negative** | 113 | - | - | | |
| **Adjuvant therapy** | **Yes** | 47 | 3.51 (1.59-7.74) | **0.002** | 0.217 | **<.001** |
| | **No** | 142 | - | - | | |
| **Age at Diagnosis** | | 189 | 1.06 (0.99-1.13) | 0.094 | 0.213 | - |
| **PSA at Diagnosis** | | 178 | 1.03 (0.54-1.97) | 0.923 | 0.785 | - |

**d**

with Gleason Group

| | **HR** | **P value** | **CI. Lower. HR** | **CI. upper. HR** |
|---|---|---|---|---|
| ***CHD1* Deletion vs No Deletion** | 2.1 | **0.012** | 1.2 | 3.7 |
| **Age at Diagnosis** | 1.0 | 0.992 | 1.0 | 1.0 |
| **Race EA vs AA** | 1.4 | 0.194 | 0.8 | 2.4 |
| **Diagnosis PSA** | 1.4 | 0.106 | 0.9 | 2.0 |
| **Pathological Stage pT3-pT4 vs pT2** | 3.0 | 0.003 | 1.4 | 6.1 |
| **GG4-GG5 vs GG1-GG3** | 1.3 | 0.350 | 0.7 | 2.4 |
| **Surgical Margin (positive vs negative)** | 1.4 | 0.261 | 0.8 | 2.6 |

with pathological Gleason score

| | **HR** | **P value** | **CI. Lower. HR** | **CI. upper. HR** |
|---|---|---|---|---|
| ***CHD1* Deletion vs No Deletion** | 1.9 | **0.032** | 1.1 | 3.5 |
| **Age at Diagnosis** | 1.0 | 0.801 | 1.0 | 1.0 |
| **Race EA vs AA** | 1.4 | 0.260 | 0.8 | 2.3 |
| **Diagnosis PSA** | 1.4 | 0.084 | 1.0 | 2.1 |
| **Pathological Stage pT3-pT4 vs pT2** | 3.1 | 0.002 | 1.5 | 6.5 |
| **Pathological Gleason 3+4 vs 3+3** | 1.2 | 0.629 | 0.6 | 2.3 |
| **Pathological Gleason 4+3 vs 3+3** | 2.6 | 0.029 | 1.1 | 5.9 |
| **Pathological Gleason 8-10 vs 3+3** | 2 | 0.092 | 0.9 | 4.3 |
| **Surgical Margin (positive vs negative)** | 1.4 | 0.314 | 0.7 | 2.5 |

**e**

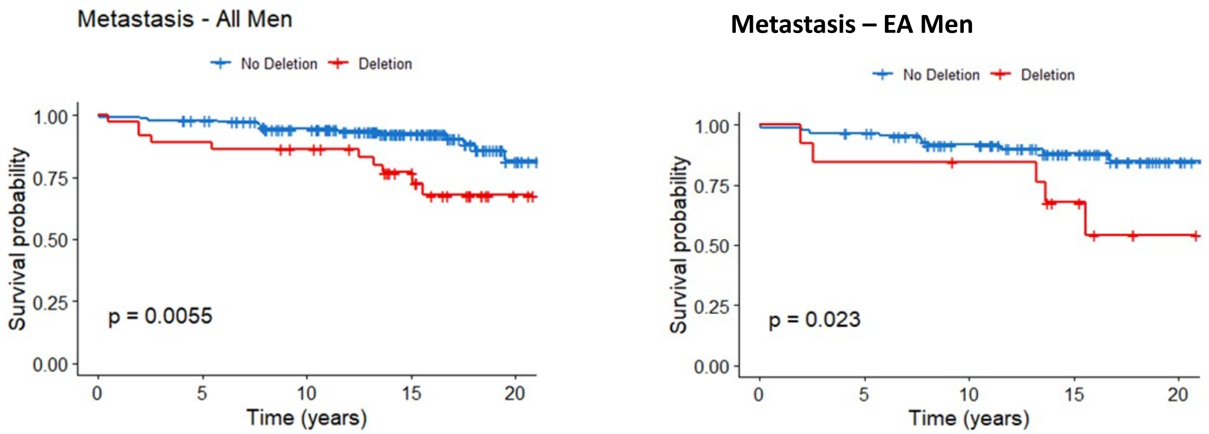

**f**

with Gleason Group

| | HR | P value | CI. Lower. HR | CI. upper. HR |
|---|---|---|---|---|
| ***CHD1* Deletion vs No Deletion** | 2.8 | **0.032** | 1.1 | 7.2 |
| **Age at Diagnosis** | 1.0 | 0.340 | 1.0 | 1.1 |
| **Race EA vs AA** | 1.6 | 0.372 | 0.6 | 4.2 |
| **Diagnosis PSA** | 0.8 | 0.452 | 0.4 | 1.6 |
| **Pathological Stage pT3-pT4 vs pT2** | 1.9 | 0.328 | 0.5 | 7.1 |
| **GG4-GG5 vs GG1-GG3** | 1.6 | 0.295 | 0.6 | 4.3 |
| **Surgical Margin (positive vs negative)** | 2.2 | 0.157 | 0.7 | 6.4 |

with pathological Gleason score

| | HR | P value | CI. Lower. HR | CI. upper. HR |
|---|---|---|---|---|
| ***CHD1* Deletion vs No Deletion** | 2.6 | **0.048** | 1.0 | 6.9 |
| **Age at Diagnosis** | 1.0 | 0.341 | 1.0 | 1.1 |
| **Race CA vs AA** | 1.2 | 0.671 | 0.5 | 3.4 |
| **Diagnosis PSA** | 0.7 | 0.306 | 0.3 | 1.4 |
| **Pathological Stage pT3-pT4 vs pT2** | 1.8 | 0.380 | 0.5 | 6.9 |
| **Pathological Gleason 3+4 vs 3+3** | 0.9 | 0.849 | 0.2 | 3.4 |
| **Pathological Gleason 4+3 vs 3+3** | 0.9 | 0.911 | 0.1 | 5.5 |
| **Pathological Gleason 8-10 vs 3+3** | 2.7 | 0.125 | 0.8 | 10.1 |
| **Surgical Margin (positive vs negative)** | 1.9 | 0.239 | 0.6 | 5.6 |

#### 2.1 Cohort selection and Tissue Microarray (TMA) generation

The aggregate cohort was composed of 2 independently selected cohort samples from Bio-specimen bank of Center for Prostate Disease Research and the Joint Pathology Center. Wholemount prostates were collected from 1996 to 2008 with minimal follow-up time of 10 years. The first cohort of 42 AA and 59 EA cases was described before [2, 3]. Similarly, the second cohort of 50 AA and 50 EA cases was selected based on the tissue availability (>1.0 cm tumor tissue) and tissue differentiation status (1/3 well differentiated, 1/3 moderately differentiated and 1/3 poorly differentiated). All the selected cases had the signed patient consent forms for tissue research applications. Patients who have donated tissue for this study also contributed to the long term follow-up data (mean 14.5 years). Our study was reviewed and approved by institutional review board (IRB) of WRNMMC and Uniformed Services University of the Health Sciences, Bethesda, MD. TMA block was assigned as 10 cases each slide and each case with 2 benign tissue cores, 2 Prostatic intraepithelial neoplasia (PIN) cores if available and 4-10 tumor cores covering the index and non-index tumors from formalin fixed paraffin embedded (FFPE) wholemount blocks. The description of numbers of patients, tumors and tumor cores of combined cohort was in Supplementary table 1c. All the blocks were sectioned into 8 $\mu$M tissue slides for FISH staining.

#### 2.2 Fluorescence in situ hybridization (FISH) assay

A gene-specific FISH probe for *CHD1* was generated by selecting a combination of bacterial artificial chromosome (BAC) clones (Thermo Fisher Scientific, Waltham, MA) within the region of observed deletions near 5q15-q21.1, resulting in a probe matching ca. 430 kbp covering the *CHD1* gene as well as some upstream and downstream adjacent genomic sequences including the complete repulsive guidance molecule B (RGMB) gene. Due to the high degree of homology of chromosome 5-specific alpha satellite centromeric DNA to the centromere repeat sequences on other chromosomes, and the resulting potential for cross-hybridization to other centromere sequences, particularly on human chromosomes 1 and 19, a control probe matching a stable genomic region on the short arm of chromosome 5 – instead of a centromere 5 probe - was used for chromosome 5 counting (supplementary figure 3). The FISH assay of *CHD1* was performed on TMA as previously described [2]. The green signal was from probe detecting control chromosome 5 short arm and the red signal was from probe detecting *CHD1* gene copy. The FISH-stained TMA slides were scanned with Leica Aperio VERSA digital pathology scanner for further evaluation. The criteria for *CHD1* deletion were that in over 50% of counted cancer cells (with at least 2 copies of chromosome 5 short arm detected in one tumor cell) more than one copy of *CHD1* gene had to be undetected. Focal deletions were called when more than 25% of evaluable tumor cells showed loss of allele or when more than 50% evaluable tumor cells in each gland of a cluster of two or three tumor glands showed loss of allele. Benign prostatic glands and stroma served as built-in control.

The sub-clonality of *CHD1* deletion was presented with a heatmap showing *CHD1* deletion status in all the given tumors sampled from whole-mount sections of each patient. The color designations were denoted as: red color (full deletion) meaning all the tumor cores carrying *CHD1* deletion within a given tumor, yellow color (sub-clonal deletion) meaning only partial tumor cores carrying *CHD1* deletion within a given tumor and green color (no deletion) meaning no tumor core carry *CHD1* deletion (Figure 1b).

#### 2.3 Statistical Analysis

The correlations of *CHD1* deletion and clinic-pathological features, including pathological stages, Gleason score sums, Grade groups, margin status, and therapy status were calculated using an unpaired t-test or chi-square test. Gleason Grade Groups were derived from the Gleason patterns for cohort from Grade group 1 to Grade group 5. Due to the small sample sizes within each Grade group, Grade group 1 through Grade group 3 were categorized as one level as well as Grade group 4 through Grade group 5. A BCR was defined as either two successive post-RP PSAs of $\geq$0.2 ng/mL or the initiation of salvage therapy after a rising PSA of $\geq$0.1 ng/mL. A metastatic event was defined by a review of each patient's radiographic scan history with a positive metastatic event defined as the date of a positive CT scan, bone scan, or MRI in their record. The associations of *CHD1* deletion and clinical outcomes with time to event outcomes, including BCR and metastasis, were analyzed by a Kaplan–Meier survival curves and tested using a log-rank test. Multivariable Cox proportional hazards models were used to estimated hazard ratios (HR) and 95% confidence intervals (Cis) to adjust for age at diagnosis, PSA at diagnosis, race, pathological tumor stage, grade group, and surgical margins. We checked the proportional hazards assumption by plotting the log-log survival curves. A P-value < 0.05 was considered statistically significant. Analyses were performed in R version 4.0.2.

#### 2.4 Immunohistochemistry for ERG

ERG immunohistochemistry was performed as previously described [4]. Briefly, four micrometer TMA sections were dehydrated and blocked in 0.6% hydrogen peroxide in methanol for 20 min. and were processed for antigen retrieval in EDTA (pH 9.0) for 30 min in a microwave followed by 30 min of cooling in EDTA buffer. Sections were then blocked in 1% horse serum for 40 min and were incubated with the ERG-MAb mouse monoclonal antibody developed at CPDR (9FY, Biocare Medical Inc.) at a dilution of 1:1280 for 60 min at room temperature. Sections were incubated with the biotinylated horse anti-mouse antibody at a dilution of 1:200 (Vector Laboratories) for 30 min followed by treatment with the ABC Kit (Vector Laboratories) for 30 min. The color was developed by VIP (Vector Laboratories,) treatment for 5 minutes, and the sections were counter stained by hematoxylin. ERG expression was reported as positive or negative. ERG protein expression was correlated with clinico-pathologic features.

### 3 Samples involved in the NGS part of the study

Altogether, 498 whole exomes (WES) and 63 whole genomes (WGS) were considered in the study.

#### 3.1 Whole exomes

The whole exomes comprised the entire TCGA PRAD-US cohort with available WES bams, and were downloaded from the GDC data portal. In case a patient had multiple normal and/or tumor bams, the more recent sample was considered only, therefore we ended up exactly 1 normal and 1 tumor sample for every patient. These samples were aligned to the grch38 reference genome.

The self-declared ancestries of the cohort:

- African American: $N_{AA} = 52$
- European American: $N_{EA} = 387$
- Asian $N_{AS} = 12$
- American or Alaska native $N_{AN} = 1$
- Not reported $N_{\text{not reported}} = 46$

#### 3.2 Whole genomes

The whole genomes were aligned to grch37, and collected from 4 different sources. These were the following:

- **TCGA PRAD-US** WGS (N = 20)
  Available from the ICGC data portal
  Self declared ancestries:
  - African American: $N_{AA} = 2$
  - European American: $N_{EA} = 18$
- Specimens from the **Dana Farber Cancer Institute, US** (DFCI, N = 19)
  Self declared ancestries:
  - African American: $N_{AA} = 9$
  - European American: $N_{EA} = 10$
- Specimens from the **Center for Prostate Disease Research, US** (CPDR, N = 14)
  Self declared ancestries:
  - African American: $N_{AA} = 7$
  - European American: $N_{EA} = 7$
- Specimens from the **Decker et al. article [5], Mayo Clinic, US** (N = 10)
  Available at the dbGaP website, under accession number phs001105.v1.p1
  Self declared ancestries:
  - African American: $N_{AA} = 0$
  - European American: $N_{EA} = 10$

### 4 Evaluating the validity of the self-declared ancestries

#### 4.1 Ancestries of the whole exomes

The following ancestry-terms and abbreviations are used interchangeably throughout the supplementary materials, and the same color-code is used for every ancestry group in the figures:

- African American: AA, african
- European American: EA, caucasian, CA, white
- American or Alaska Native: AN, American Native
- Asian American: AS, asian

In addition, for patients, who have not reported their ancestries, a gray color-code is used.
Out of the 498 PRAD-US patients 452 reported their ancestry ($N_{AA} = 52$, $N_{EA} = 387$, $N_{AS} = 12$, $N_{AN} = 1$). The remaining patients $N_{nr} = 46$ have not reported anything about their race, or the data was missing (Suppl.Fig. 24).

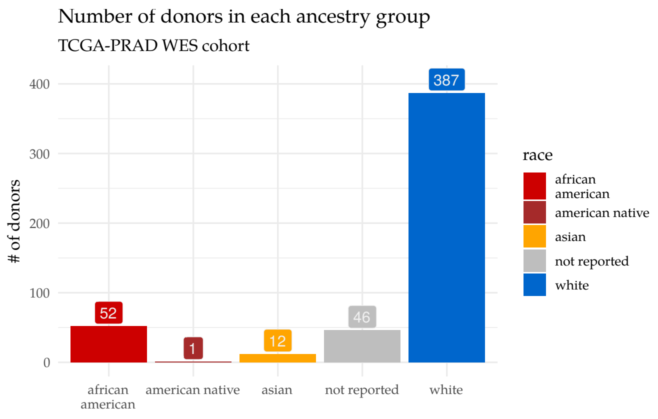

**Suppl.Fig. 6:** Distribution of the self-declared ancestries within the TCGA WES dataset.

Since patients report their ancestries based on their best believes, the collected information about their genetic background is oftentimes incorrect [6]. In order to alleviate all discrepancies, and to keep the 46 patients who haven't reported anything in the study, we have decided to re-classify the donors based on their germline genotypes.

##### 4.1.1 Targeted SNPs

Our strategy was to determine the genotypes of every sample at key genomic SNP-coordinates, that are significantly more abundant in any of the three common (EA, AA, and AS) ancestry groups, then use a Bayes Classifier to identify the most probable ancestries of the "not reported" cases. Furthermore, we could use the genotypes to identify the outliers among the cases that had self-declared ancestries.

We used the database of the Exome Aggregation Consortium (ExAC) [7] to select a group of variants that are significantly more common in the three ancestries. In order to increase the accuracy in the population allele frequency of each SNP, we only considered variants that were supported at least 4,000 donors within the AA, and 10,000 donors within the AS and EA populations (Suppl.Fig.7).

Suppl.fig.9 illustrates this selection process, by showing the 10 most common single nucleotide variants in each group, that are almost absent in the other two. Since the reference genome itself is biased towards the Caucasian genome, it is not surprising, that we saw significantly higher allele frequencies in African Americans than in European Americans. To achieve robust results, the 1000-1000 most common variants were selected as predictors in each ancestry group, and their genomic coordinates with their immediate surrondings (+5000 bp in both the 3' and 5' directions) were collected into bedGraph format.

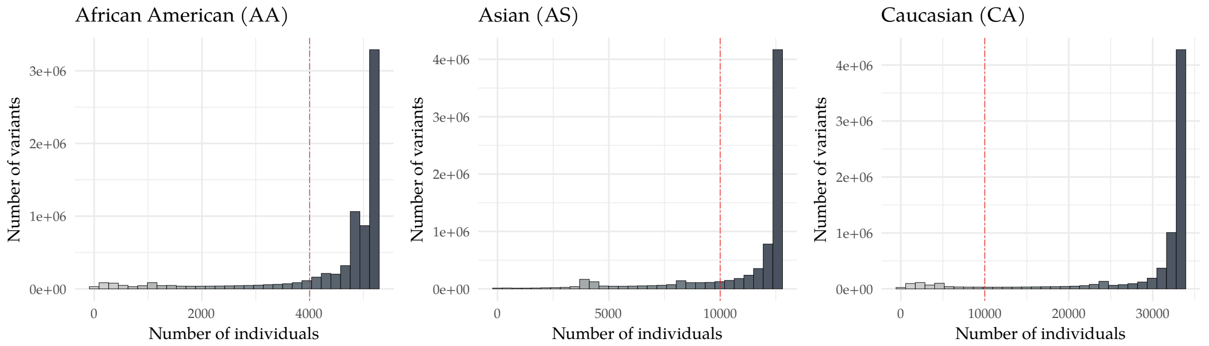

**Suppl.Fig. 7:** Distributions of the number of individuals supporting SNPs in the ExAC database in each ancestry group. The red dashed vertical lines represent a threshold, below which the variants were excluded from the search.

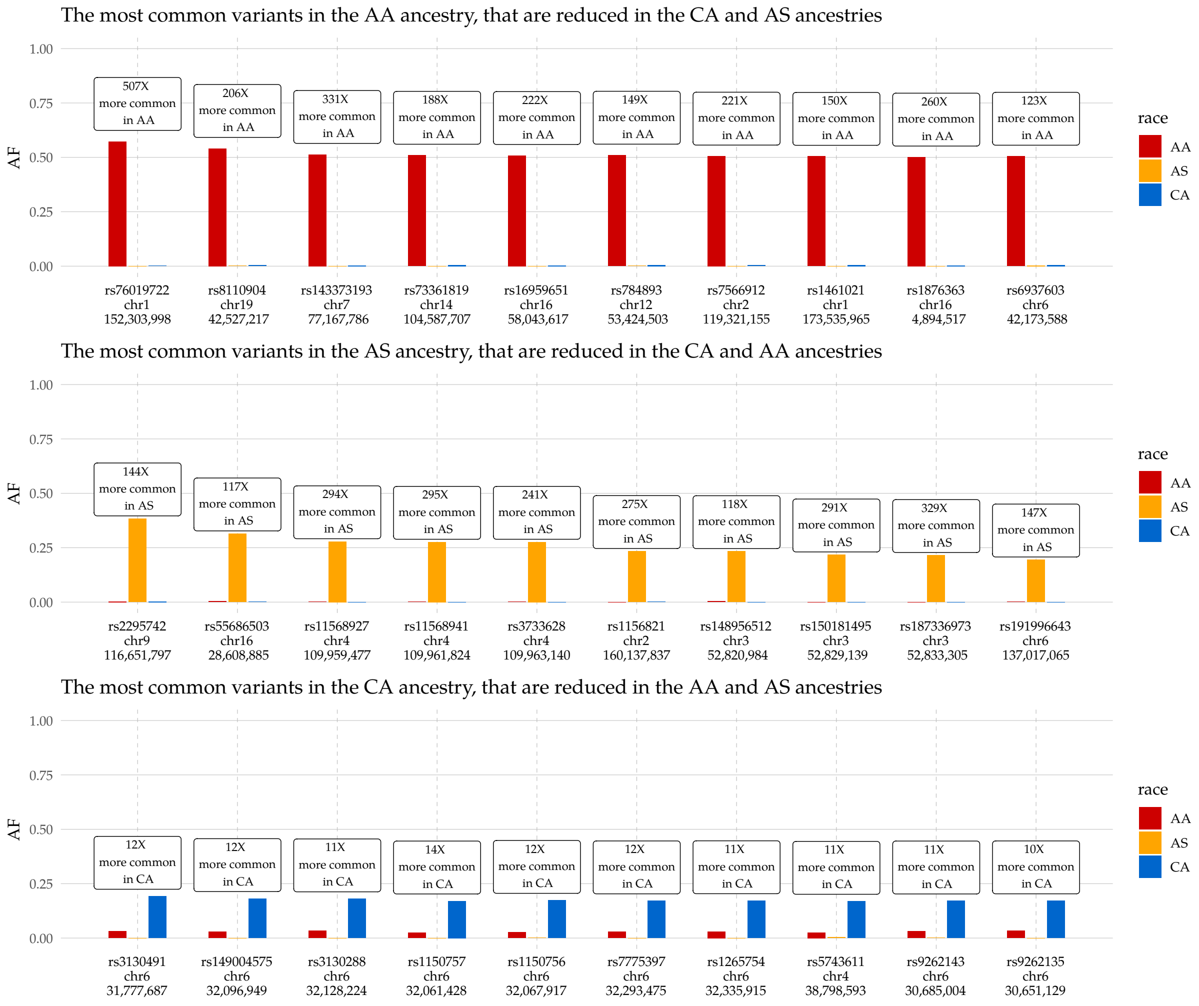

**Suppl.Fig. 8:** The ten most ancestry-specific SNPs of the three most frequent WES ancestry groups. The relative commonnes of each SNP was compared to the mean of the population allele frequencies (AF) of the other two ancestries.

---

The bedGraph file then was forwarded as a list of targeted regions to GATK HaplotypeCaller [8] (v4.1.0) to call germline mutations in each sample, and to extract the genotypes of the targeted SNPs. The surrounding regions were only evaluated, because we also wanted to see the degree of the difference in the number of germline mutations among the three ancestries. Not surprisingly, we found significant differences between the African American and

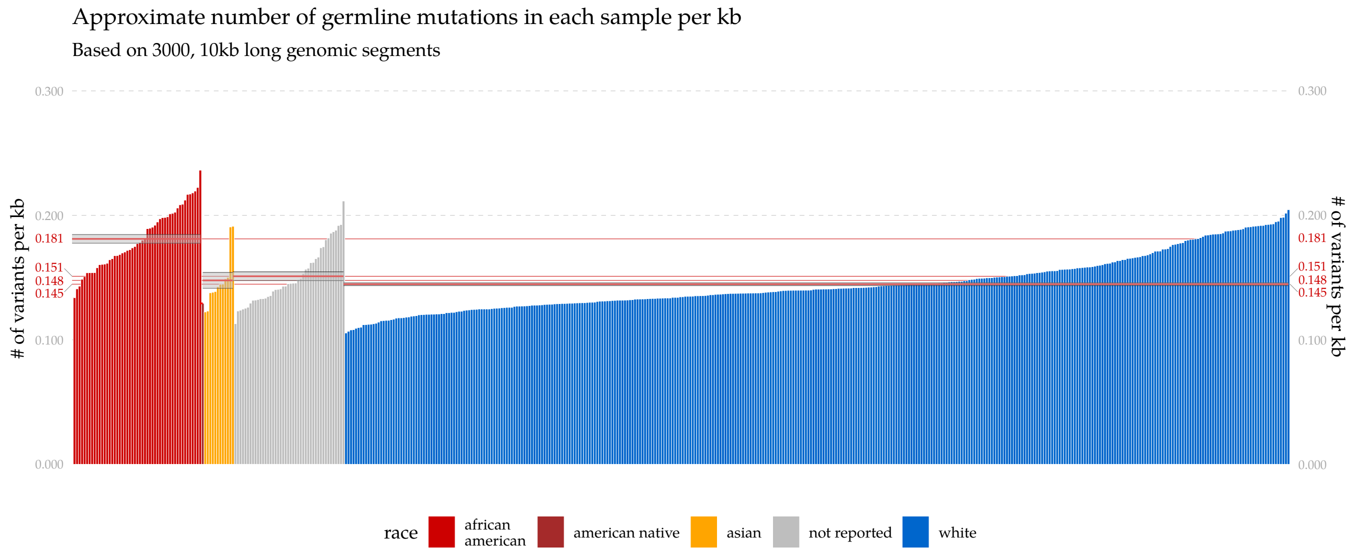

**Suppl.Fig. 9:** Frequencies of the exonic germline mutations within the PRAD-US whole exomes, grouped by the self-reported or not-reported ancestries.

European American samples (Suppl.Fig. 8). The mean numbers of germline variants in the different ancestries were the following:

- AA: $0.181 \pm 0.003 \frac{1}{kb}$
- EA: $0.145 \pm 0.001 \frac{1}{kb}$
- AS: $0.148 \pm 0.008 \frac{1}{kb}$
- not reported: $0.151 \pm 0.001 \frac{1}{kb}$

These results suggested, that the majority of the "not reported" cases belong to the asian or white ancestries, however a few cases might be likely African American (based on these numbers we could assume that we find approximately 8 AA, 37 EA and 1 AS patients among them).

###### 4.1.2 Genotyping and PCA

The collected 3000 SNPs then were used to create a single genotype matrix (**G**) with 498 rows (patients) and 3000 columns (genotypes). When the genotype of SNP $j$ of patient $i$ was REF/REF, the corresponding element of the matrix ($G[i, j]$) was set to 0, for heterozygous ALT/REF variants it was set to 1, and it was set to 2 for ALT/ALT homozygotes. Next, we executed a singular value decomposition on matrix **G** and determined its eigenvalues and eigenvectors, i.e. its principal components (PC, Suppl.Fig. 10). The projections of the data onto 2-dimensional plains set by the first few principal components is shown on Suppl.Fig. 11. It was clear, that the first principal component explained a cardinally greater proportion of the variants than any other PC, and it was the component that separated the AA patients from the EA patients the best. PC2, while representing a small fraction of the variance, separated the asian samples from the other two ancestries.

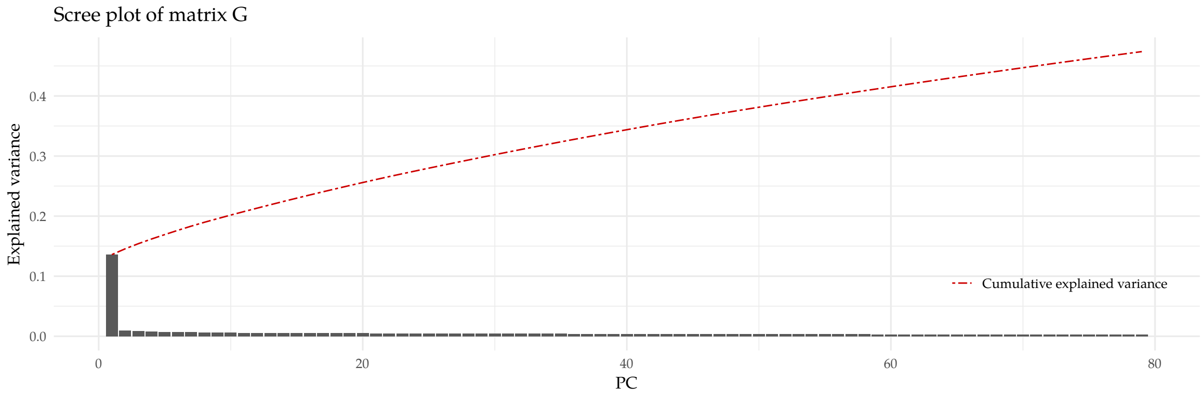

**Suppl.Fig. 10:** Scree plot of the ancestry-genotype matrix **G**

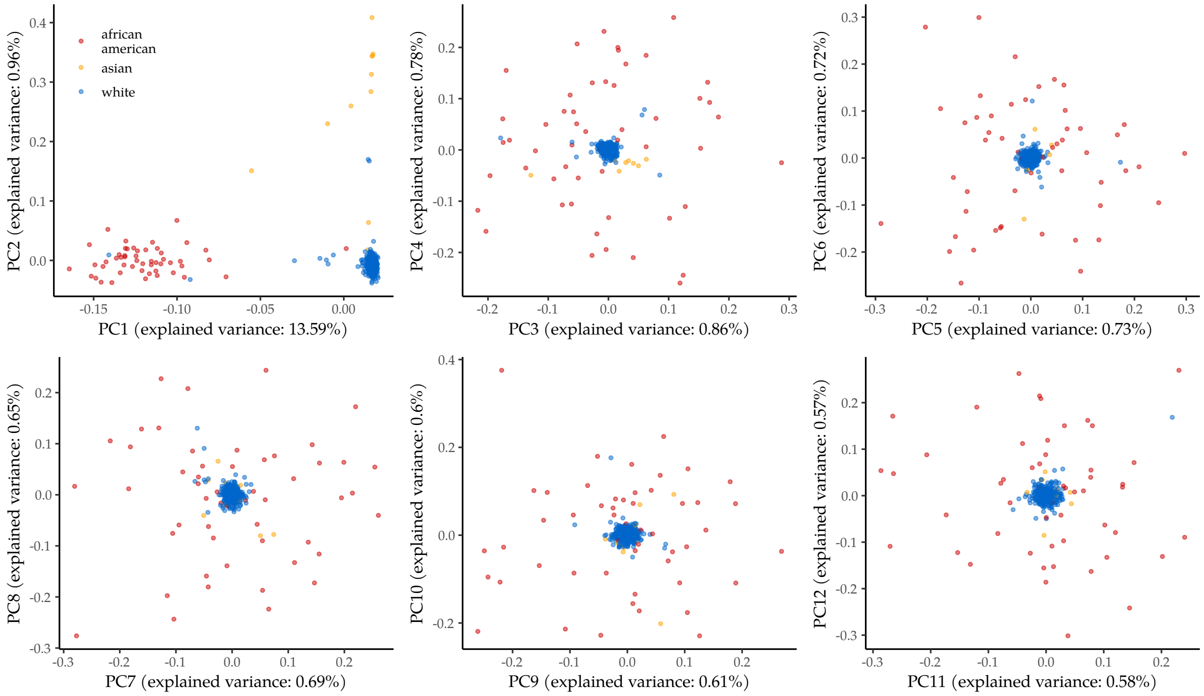

**Suppl.Fig. 11:** Projection of the ancestry genotype matrix onto its first few principal components.

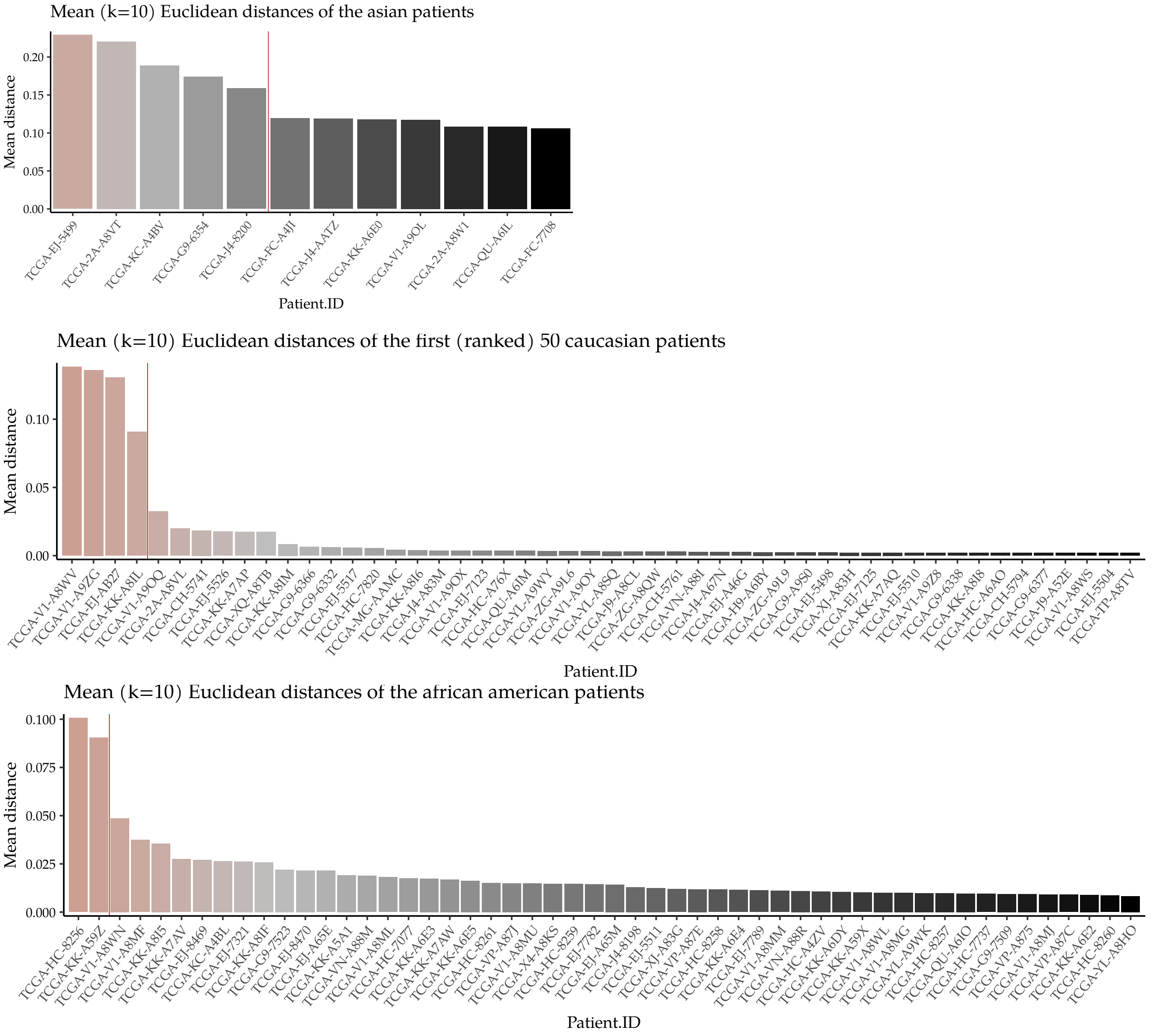

**Suppl.Fig. 12:** Euclidean mean distances in the PC1-PC2 space. Large mean distances indicate, that the sample is located further from its cluster than the rest of the members of that group. Red vertical lines indicate the thresholds that were used for outlier detection. The ancestry of samples left from the threshold were reclassified as "outliers". These samples were excluded from the training process introduced in the next section.

##### 4.1.3 Outlier Detection

From Suppl.Fig. 11 it was clear, that some of the patients were positioned further from the cluster of their own ancestry than one of the other ancestries', which is likely contributable to the problems in the nature of the self ancestry declaration process. We have decided therefore, to filter out these "outliers" from the remaining samples, by using an Euclidean mean difference-based outlier filtering strategy, during which we focused only on the PC1-PC2 space, and measured the mean distance of every point from their 10 closest neighbors that belonged to the same ancestry. The resulting ranked distribution of the mean distances of the samples is illustrated in Suppl.Fig. 12.

The outlier-filtering thresholds were set at ranked positions, where there were significant jumps in the first order derivatives of the ranked mean distances. These thresholds are also illustrated in Suppl.Fig. 12, while Suppl.Fig. 13

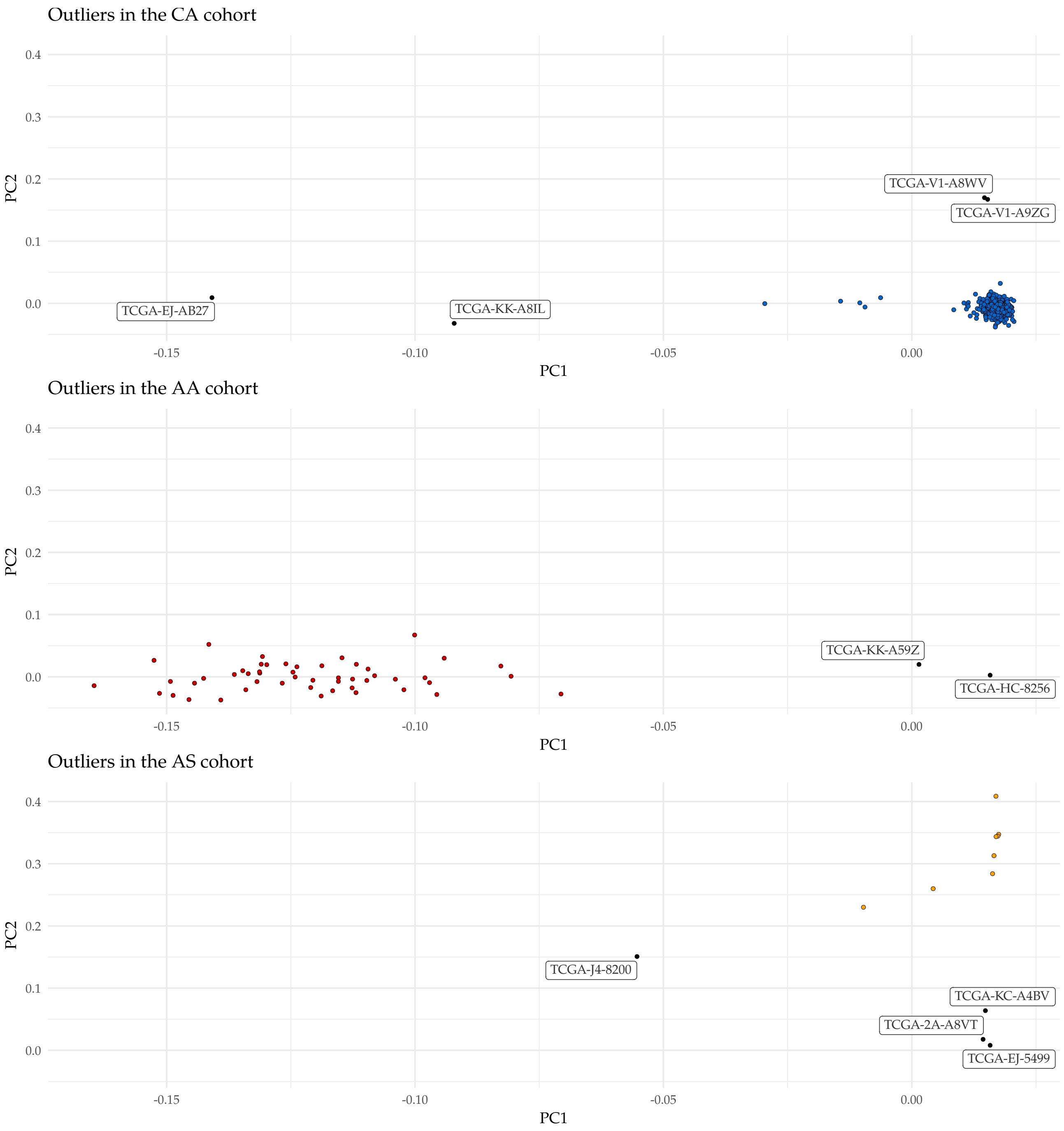

**Suppl.Fig. 13:** Outliers of the three ancestries. From top panel to the bottom, European American, African American, and Asian American ancestries. Samples that have not deviated from their main cluster significantly are colored according to the color-code of their ancestry, while clear outliers are colored black. The TCGA submitter IDs of these samples are also indicated on the figure.

shows the resulting distribution of the three clusters of the PC1-PC2 space, with the samples classified as outliers, highlighted. The ancestries of these samples were reclassified as "outliers", and were treated the same way as samples with "not reported" ancestries throughout the remaining part of this section.

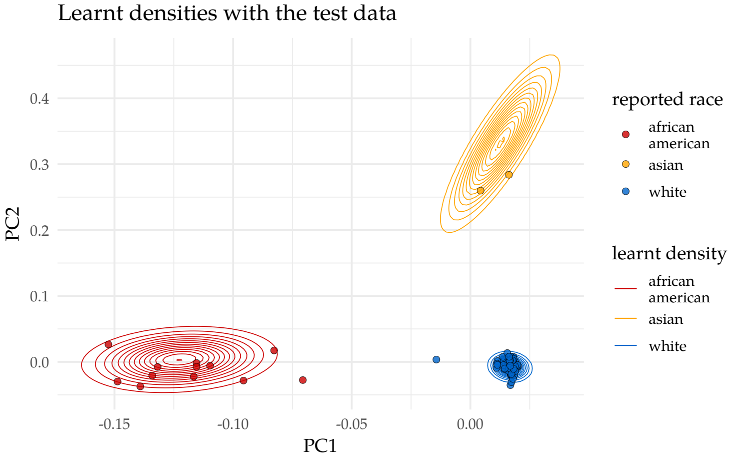

**Suppl.Fig. 14:** Learnt distributions of the ancestries in the PC1-PC2 space of the genotype matrix. Samples of the test set are also represented in the figure, colored by their original, self-declared ancestries.

##### 4.1.4 Classification of the ancestries - The (non-Naïve) Bayes Classifer

Our strategy was to teach a model how the points of the ancestries are distributed in the PC1-PC2 space, and use the learnt densities of these distributions to determine which ancestry the "not reported" and "outlier" cases most likely to belong to based on their genotypes. The three classes $C = 0, 1, 2$ were encoded the following way:

- White: 0
- African American: 1
- Asian: 2

Before the calculation of the principal components, the columns of matrix $\mathbf{G}$ were standardized according to:

$$G[i,j]^* = \frac{G[i,j] - \mathbb{E}_k G[k,j]}{\sigma_k(G[k,j])}. \tag{4.1}$$

The probability that sample $\mathbf{x}_i = \mathbf{G}[\mathbf{i}, \cdot]$ belongs to ancestry group $c_a$ is the following:

$$P(y_i = c_a | \mathbf{x}_i) = \frac{P(\mathbf{x}_i | y_i = c_a) P(y_i = c_a)}{\sum_{c' \in C} P(\mathbf{x}_i | y_i = c') P(y_i = c')} \tag{4.2}$$

The likelihood $P(\mathbf{x}_j | y_j = c_i)$ is provided by a multivariate normal density $\mathcal{N}(\mathbf{x}_j | \boldsymbol{\mu}_{c_i}, \boldsymbol{\Sigma}_{c_i})$, the maximum a posteriori (MAP) estimates of the parameters of which were calculated by the classifier algorithm. $P(y_j = c_i)$ is the relative sample size (number of patients) of ancestry $c_j$, i.e. $P(y_j = c_i) = n_{c_i} / \sum_k n_{c_k}$, where $n_{c_k}$ is the number of patients belonging to ancestry $c_k$.

##### 4.1.5 Training and Prediction

The 441 AA, EA, and AS samples were randomly separated into training ($N_{training} = 352$ - 80%) and test ($N_{test} = 89$ - 20%) sets, and the classifier was programmed in R. The learnt distributions along with the test data are illustrated in Suppl.Fig.14. The probabilities representing a sample's connection to any of the ancesty groups were calculated according to equation (4.2), and the results are illustrated in Suppl.Fig. 15. Based on these probabilities, the accuracy of the resulting classifier was estimated to be within the [0.949, 0.999] range (Suppl.Fig 16).

##### 4.1.6 Predictions of the "not reported" and "outlier" cases

The predicted ancestries of the 46 "not reported" and 10 "outlier" samples are visualized in Suppl.Fig. 17 and 18. The final number of patients per ancestry groups is illustrated in Suppl.Fig. 17.

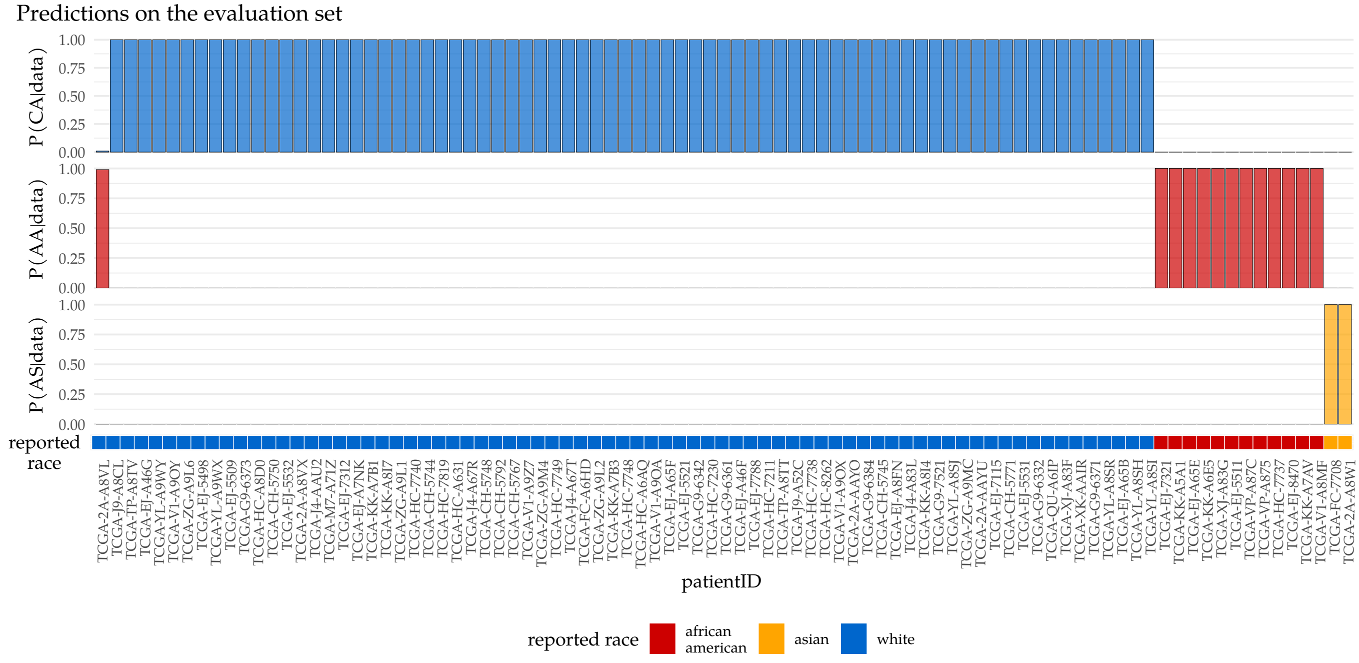

**Suppl.Fig. 15:** Predicted probabilities of samples of the test set. The reported, i.e. self-declared ancestries are also illustrated under the bar plots.

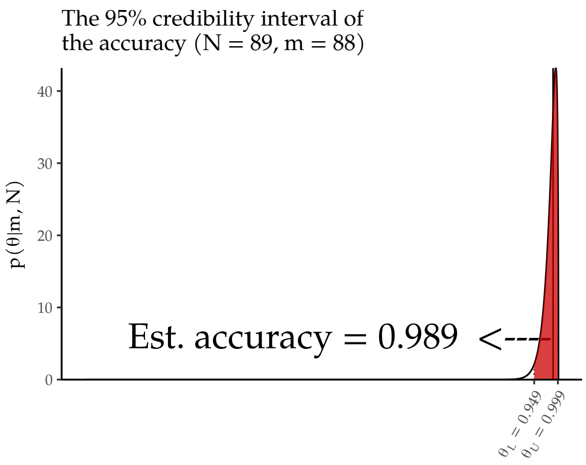

**Suppl.Fig. 16:** Estimated Accuracy ($\theta$) of the Bayes classifier. The 95% credibility interval was estimated using a binomial distribution of $N = 89$ trials and $m = 88$ successes.

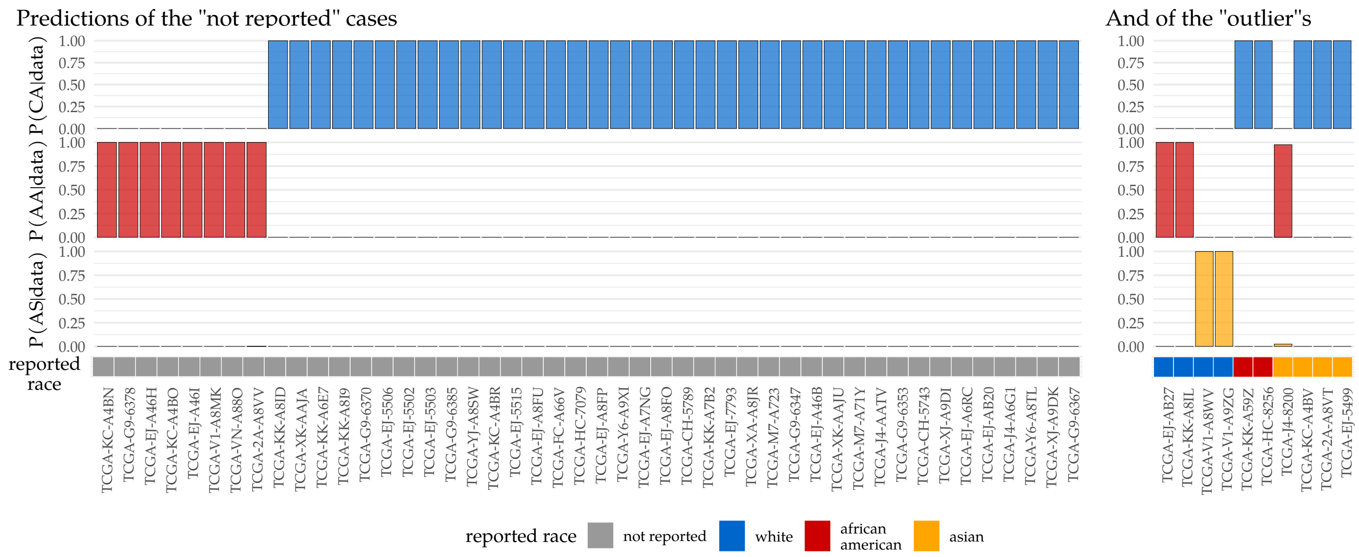

**Suppl.Fig. 17:** Predicted probabilities of samples of the "outlier" and "not reported" samples. The reported, i.e. self-declared ancestries are also illustrated under the bar plots.

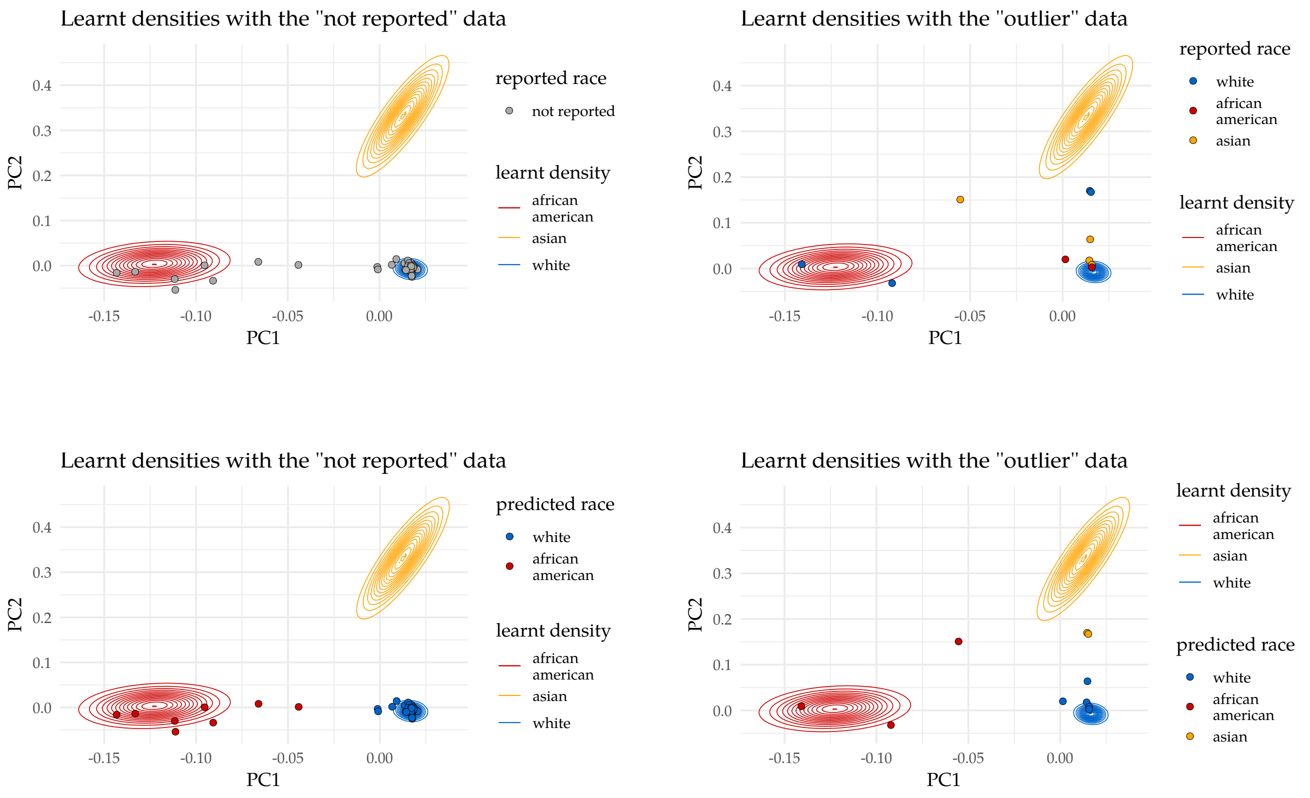

**Suppl.Fig. 18:** Learnt distributions of the ancestries in the PC1-PC2 space of the genotype matrix. Samples of the "not reported" and "outlier" sets are also represented in the figures, in the top two figures they are colored according to their original, self-declared ancestries, while in the bottom two figures they are colored according to their reclassified ancestries.

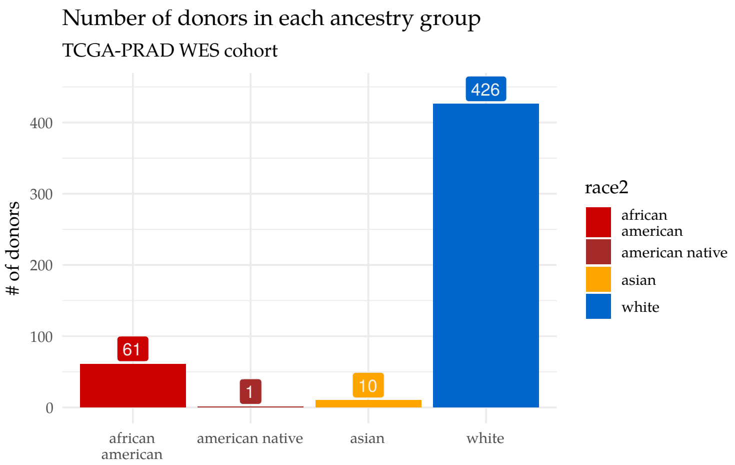

**Suppl.Fig. 19:** Final distribution of the validated ancestries in the TCGA WES dataset

#### 4.2 Ancestries of the whole genomes

The evaluation of the validity of the self-declared ancestries within the whole genome cohorts followed the same strategy we used for the whole exomes. The same set of 3000 SNPs were used as targets, and the resulting genotypes were collected into a similar **G** matrix to that of the whole exomes. Initially, the number of ancestries were the following:

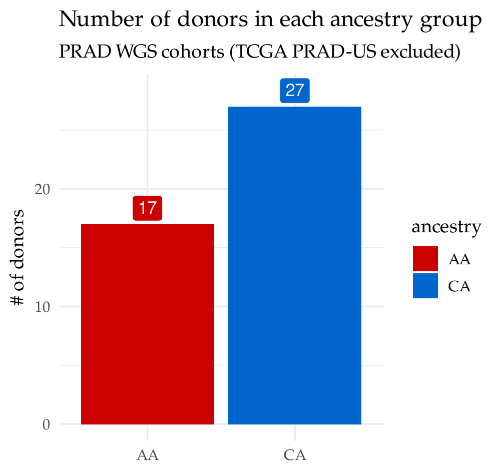

**Suppl.Fig. 20:** Number of patients with AA/CA self-declared ancestries among the PRAD WGS samples. As the 20 TCGA donors have been analyzed in the whole exome section, the were not involved in the WGS section.

The collected **G** genotypes were standardized according to equation (4.1), and the MAP-estimates of the ancestry densities determined in the previous section were used to identify the two populations. The distribution of the points in the PC1-PC2 space is shown in Suppl.Fig.21, which was immediately enough to determine that there were no discrepancies in the self-declared ancestries in any of the WGS samples. This is further proved by the predicted probabilities that are shown in Suppl.Fig.22.

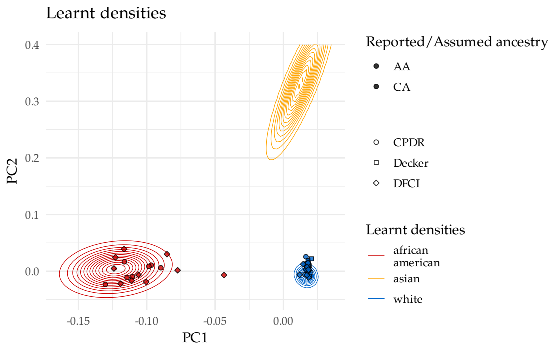

**Suppl.Fig. 21:** Distribution of the self-declared ancestries of the PRAD WGS samples. As the 20 TCGA donors have been already analyzed in the whole exome section, they were not included in the WGS section.

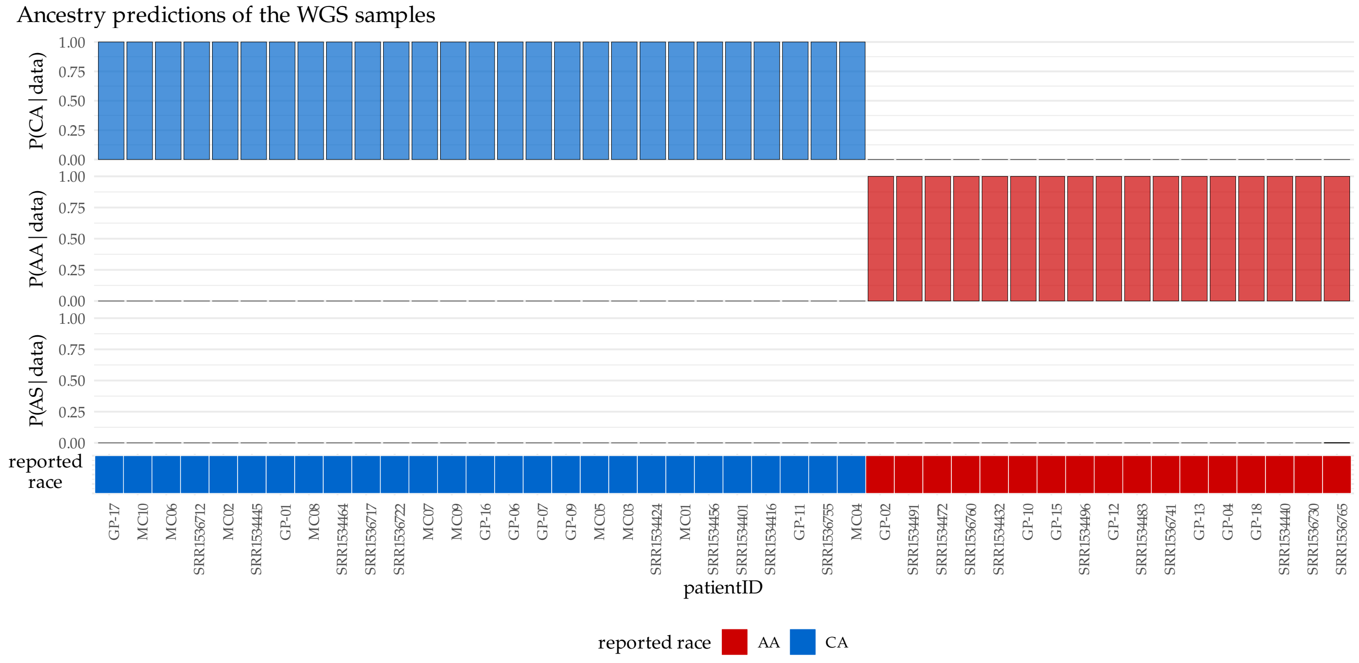

**Suppl.Fig. 22:** Predicted probabilities of samples of the "outlier" and "not reported" samples. The reported, i.e. self-declared ancestries are also illustrated under the bar plots.

The ancestry-classification model identified 100% of the samples in accordance with their declared ancestries. All the self-declared ancestries of the whole genomes appeared to be accurate.

### 5 Determining the sub-clonal loss of *CHD1*

**Summary:** In this section of the supplementary materials we explain the strategy we used to identify the subclonal loss of *CHD1* in the TCGA whole exomes, and in the various cohorts of whole genomes. Briefly, a linear model was fitted on the corresponding normalized germline and tumor coverages, and the fitted slope, after it was corrected for the cellularity of the tumor, was used to infer the approximate level of loss in the tumor.

The paired germline and tumor binary alignment (bam) files were analyzed using bedtools genomcecov (v2.28.0) [9], and their mean sequencing depths were determined. The coverage above and within the direct vicinity of *CHD1* (chr5:98,853,485-98,930,272 in grch38 and chr5:98,190,408-98,262,740 in grch37) was collected in 50 bp wide bins into d-dimensional vectors ($d_{grch37}$ = 1447, $d_{grch38}$ = 1536) using an in-house tool and samtools (v1.6) [10], and were normalized using their corresponding mean sequencing depths.

We aimed to develop a reliable measure that reflects the level of loss in the vicinity of *CHD1* on a continuous scale. On a simplistic level, this could be realized by fitting a linear curve on the corresponding normalized tumor ($d_t$) and normal coverages ($d_n$) in the following form:

$$d_t = \alpha + \beta_0 d_n. \tag{5.1}$$

Here, $\beta_0$, i.e. the slope of the curve is a rough measure of the level of *CHD1* in the tumor. When it is close to 1, it indicates that neither of the copies of *CHD1* are lost in the tumor. When $\beta_0 \in [0, 1)$, then there might be a loss in *CHD1*, when $\beta_0 > 1$, there might be an amplification in *CHD1*. However, such a model would be inaccurate, as it does not consider the degree of normal contamination (1-cellularity) in the tumor sample, and the estimate would be a subject of minor fluctuations in the coverage when the loss only affects a small subclone. Therefore, the simple model described by equation (5.1) was extended by three additional steps.

First, in order to correct for minor fluctuations and outliers, instead of fitting a curve on the single position coverage-pairs, the corresponding depths were collected into 50 bp long bins, and were normalized by the mean coverage of the entire genome or exome. These bins were used to estimate $\beta_0$.

Second, we estimated the significance of the presumed deletions in every tumor by fitting similar patient-wise linear curves on the corresponding tumor normal coverages of 14 housekeeping genes (G6PD, IPO8, PGK1, PP1A, HMBS, GUSB, UBC, YWHAZ, GAPDH, HPRT1, ACTB, B2M, TBP, and TFRC). The 14 estimated slopes were standardized using their mean and standard deviation (i.e. they were transformed into their z-scores). The estimated slopes of *CHD1* then were also translated into z-scores using the previously determined parameters of their donors, and their p-values were calculated. When a resulting p-value was larger than 0.1 for whole genomes or 0.05 for whole exomes, the corresponding sample was marked as "*CHD1* intact", otherwise it was categorized as "*CHD1* loss" (Suppl.Fig. **??** and Suppl.Fig. **??**).

Third, the rough levels of the estimated loss ($1 - \beta_0$) in *CHD1* was corrected for the cellularities ($c$) of the tumors, which themselves were estimated after the rigorous selection of the most reliable cellularity-ploidy pair offered by the somatic copynumber caller sequenza [11] in the form of alternative solutions. The following formula was used:

$$\beta_t \sim \frac{\beta_0 - 1 + c}{c}, \tag{5.2}$$

which was derived from:

$$\beta_0 = \underbrace{1 - c}_{\text{normal contamination}} + \underbrace{c \cdot \beta_t}_{\text{real contribution of the tumor}}.$$

The $\sim$ operator in equation (5.2) suggests that the true level of *CHD1* can only be determined to a certain degree of accuracy that depends on the uncertainty of $\beta_0$ and $c$. The former comes from the fitted linear model itself:

$$\begin{aligned} d_t &\sim \text{Normal}(\mu, \sigma) \\ \mu &= \alpha + \beta_0 d_n \\ \alpha &\sim \text{Normal}(0, 5) \\ \beta_0 &\sim \text{Normal}^+(0, 5) \\ \sigma &\sim \text{Normal}^+(0, 5) \end{aligned}$$

(The priors are simple, non-informative priors, $\beta_0$ and $\sigma$ are confined to positive numbers only.) Therefore, in order to gain information about the uncertainty of $\beta_0$, the easiest way is to take samples from its marginal posterior.

Since sequenza determines the joint posterior of the plodidy and cellularity of the tumor using a grid approximation strategy, we took samples from the peak of the discretized marginal posterior of the cellularity that corresponded to the accepted solution of the final copynumber segments. In order to translate these discrete values into a continuous scale, a beta distribution was fitted onto collection of the cellularity samples:

$$c \sim \text{Beta}(\alpha_c, \beta_c),$$

where $\alpha_c$ and $\beta_c$ are the two shape-parameters of the distribution. Using the distributions of $\beta_0$ and $c$, we could estimate the uncertainty in the true level of loss:

$$\text{True level of loss in CHD1} = 1 - \beta_t.$$

The summary figures that illustrate these calculations are available for the TCGA exomes that were identified with *CHD1* loss (Suppl.Fig.) and for all the whole genomes (Suppl.Fig.) on the following pages.

### 6 *CHD1* loss summary - NGS samples

Altogether 530 PRAD NGS cases have been analyzed. Out of the 77 AA patients, 20 were identified with *CHD1* loss (26%), while out of the 453 CA cases, 73 showed clear signs of decreased coverage in the vicinity of the gene (16.1%).

| | AA | CA |
|---|---|---|
| ***CHD1* intact** | 57 | 380 |
| ***CHD1* loss** | 20 | 73 |

The probability of obtaining the observed frequencies for our data assuming no difference between the groups was assessed with a Fisher exact test:

$$\boxed{p = 0.029,}$$

and with a simple Bayesian model, using two independent binomial densities to model the inferred distributions of the likely frequencies of *CHD1* loss in the two ancestry groups, based on the data. The model description:

$$\begin{aligned} N^{CA}_{loss} &\sim \text{Binomial}(N_{CA}, \text{Prop}_{CA}) \\ N^{AA}_{loss} &\sim \text{Binomial}(N_{AA}, \text{Prop}_{AA}) \end{aligned},$$

where $N^{CA}_{loss}$ and $N^{AA}_{loss}$ are the number patients identified with *CHD1* loss in the CA and AA ancestry groups respectively, $N_{CA}$ and $N_{AA}$ are the total number of patients in the two ancestry groups, while $Prop_{AA}$ and $Prop_{CA}$ are the inferred proportions. The posterior distributions, and the difference between the two, are visualized on Suppl.Fig.23.

Based on our data, the model above estimates, that there's a 0.981 probability, that *CHD1* is more frequently lost in the prostate adenocarcinomas of African American men, than in patients with European ancestry.

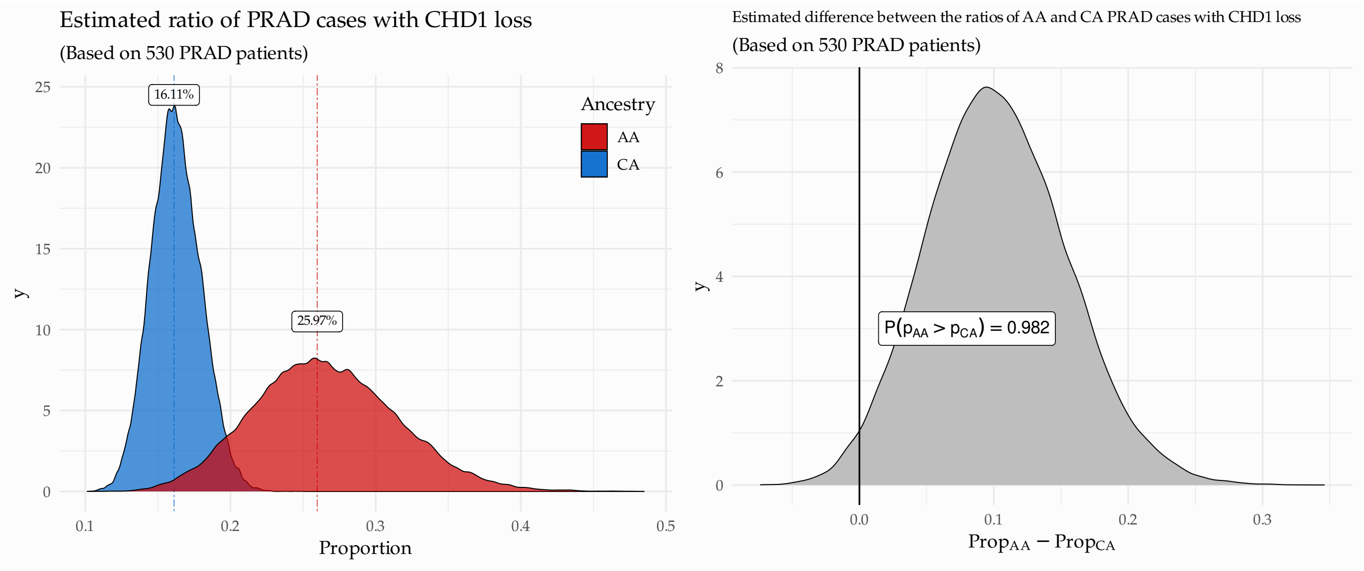

**Suppl.Fig. 23:** On the left: The estimated distributions of the prevalence of *CHD1* loss cases in AA and CA PRAD cases. The figure illustrates the likely range of the true ratio of *CHD1*-loss in AA and CA tumors in the greater population.
On the right: The difference of the two posteriors $Prop_{AA} - PropCA$.

### 7 Genotyping

Variant (small-scale and large-scale) and copynumber calling were performed identically to the study by Sztupinszki et al. [12]. Genotypes were categorized as wild type: +|+ if no pathogenic (or likely pathogenic) variants (according to intervar) were found in the gene, monoallelic: +|- when at least one pathogenic germline or somatic variant or an LOH was identified in the gene, and biallelic: -|- when a pathogenic variant was accompanied by an LOH, or a deep deletion was found.

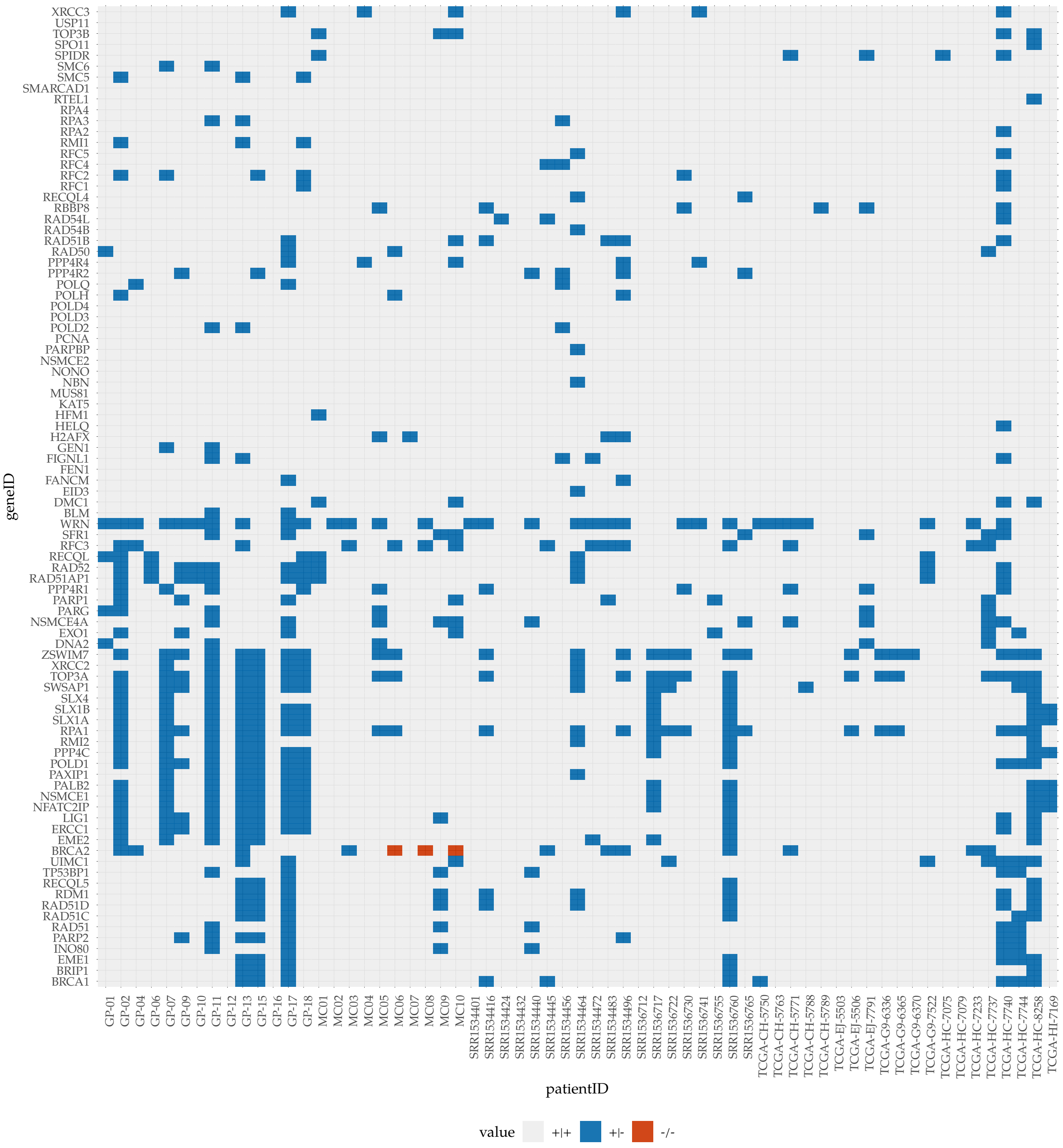

**Suppl.Fig. 24:** Genotyping of genes involved in DNA repair in the whole genomes

### 8 Assessing the local loss of heterozygosity

**Summary:** Here we describe the methodology of our strategy to assess the local heterozygosity of a loss in a tumor subclone, in this case in *CHD1*. Briefly, we collect the SNP ALT allele-frequencies in the close vicinity of the gene in the tumor, carefully focusing on regions that have suffered the most serious loss (e.g., if only a part of the gene is lost), and by using the tumor cellularity and the estimated level of loss calculated in the previous section, we assess whether a heterozygous or a homozygous subclonal deletion is most likely to result in the observed frequency pattern.

#### 8.1 Modeling the effects of an LOH on the allele-frequencies

The SNP variant allele frequencies (VAF) in the close vicinity of *CHD1* in the tumor were collected with GATK HaplotypeCaller (v4.1.0) [8]. The coverage and VAF data were carefully analyzed in order to ensure that we were strictly focusing on regions that had suffered the most serious loss (e.g., if only a part of the gene was lost, the unaffected region was excluded from the analysis). By using the tumor cellularity (c) and the estimated level of loss in the tumor ($\beta_t$), we assessed whether a heterozygous or a homozygous subclonal deletion is more likely to result in the observed frequency pattern.

The observed distribution of the SNP ALT allele frequencies in the tumor sample ($AF_{obs}$) can be thought of as stochastic variables that are generated by the following process:

$$AF_{obs} \sim \underbrace{(1-c)\cdot AF_{normal}}_{\text{normal contamination}} + c \cdot [\underbrace{L_{true}\cdot AF_{normal}}_{\text{Tumor cells with the normal phenotype}} + \underbrace{(1-L_{true})AF_{tumor\,subclone}}_{\text{Tumor cells with a loss in CHD1}}],$$

where $c$ is the cellularity of tumor sample (also a stochastic random variable, approximated by a beta-process, see in the previous section), and $L_{true}$ is the approximate level of the true level of *CHD1* within the tumor sample (i.e. the proportion of cancer cells with intact *CHD1*), after we compensate for the normal contamination. $AF_{normal}$ is the distribution of the allele frequencies of the heterozygous SNPs in the normal sample, which can also be modeled with a beta process:

$$AF_{normal} \sim \text{Beta}(\alpha_n, \beta_n),$$

centered on 0.5, i.e. $\alpha_n \simeq \beta_n$. When the loss in the tumor is homozygous, than all the reads originate from either the normal cells or the tumor cells that exhibit the normal phenotype, i.e. with intact *CHD1*. In order to ensure that $(1 - cellularity) + cellularity$ always adds up to 1, we will say that in that case, $AF_{tumor\,subclone} = AF_{normal}$, in which case the observed allele-frequency is the same as the normal allele-frequency (only in the vicinity of the target gene), i.e.:

$$\boxed{AF_{obs}^{homozygous} \sim AF_{normal}.}$$

Naturally, the same distribution occurs without a deletion in the targeted gene, in which case the only sign of the loss is a drop in the coverage in the tumor.

A heterozygous deletion can arise through either the loss of the ALT allele (in which case $AF_{tumor\,subclone} = 0$) or the REF allele (in which case $AF_{tumor\,subclone} = 1$):

$$AF_{tumor\,subclone} = \begin{cases} 0, & \text{if the ALT allele is lost} \\ 1, & \text{if the REF allele is lost.} \end{cases}$$

In case of an LOH, the distribution of the observable allele frequencies in the tumor becomes bimodal. First, let's simplify the original formula:

$$AF_{obs} = \underbrace{(1-c+cL_{true})}_{w_1}\cdot AF_{normal} + \underbrace{c(1-L_{true})}_{w_2}\cdot AF_{tumor\,subclone} = w_1 \cdot AF_{normal} + w_2 \cdot AF_{tumor\,subclone},$$

where both $w_1$ and $w_2$ are stochastic variables that only depend on the accepted cellularity and the fitted slope on the corresponding coverage segments, and

$$\sum_{i=1}^{2} w_i = 1.$$

In a heterozygous model therefore, the observable allele-frequencies will be generated by the following stochastic process:

$$\boxed{AF_{obs}^{LOH} \sim \frac{1}{2}\left(\underbrace{w_1 \cdot AF_{normal} + w_2 \cdot 1}_{\text{In case the REF allele is lost}}\right) + \frac{1}{2}\left(\underbrace{w_1 \cdot AF_{normal} + w_2 \cdot 0}_{\text{In case the ALT allele is lost}}\right).}$$

The LHS will result in variants with higher AFs, the RHS in lower AFs. The distance between the two modes depends on $w_2$. The larger that weight is, the more the modes will approach AF = 0 and AF = 1.

The final task is to compare the likelihoods that the homozygous or the heterozygous processes have generated the data, and determine their respective probabilities:

$$\mathcal{L}(AF_{obs}|\textbf{heterozygous deletion}) = \prod_{i=1}^{N}(AF_{obs_i}|AF_{obs}^{LOH})$$

and,

$$\mathcal{L}(AF_{obs}|\textbf{homozygous deletion}) = \prod_{i=1}^{N}(AF_{obs_i}|AF_{obs}^{homozygous}),$$

The probability that the deletion affects only one of the alleles, i.e. it is heterozygous, can be calculated from the likelihoods above by following the formula below:

$$P(\textbf{heterozygous deletion}) = \frac{\mathcal{L}(AF_{obs}|\textbf{heterozygous deletion})}{\mathcal{L}(AF_{obs}|\textbf{heterozygous deletion}) + \mathcal{L}(AF_{obs}|\textbf{homozygous deletion})}$$

And naturally,

$$P(\textbf{homozygous deletion}) = \frac{\mathcal{L}(AF_{obs}|\textbf{homozygous deletion})}{\mathcal{L}(AF_{obs}|\textbf{heterozygous deletion}) + \mathcal{L}(AF_{obs}|\textbf{homozygous deletion})} = 1 - P(\textbf{heterozygous deletion})$$

#### 8.2 Illustrating the process on the whole genomes

The following figures (Suppl.Fig. 25) will illustrate the process described above on each of the whole genomes that were identified with *CHD1* loss. In order, to maximize the predictive accuracy of the model, we have tried to include the surrounding SNPs as well in the analysis. However, when the coverage was significantly different in *CHD1*, than what we saw in its vicinity, we had to confine our attention to a smaller region, in the most severe case only to small part of the gene (TCGA-CH-5788).

The predictive accuracy and error of the model was also estimated by bootstrapping, in which 90% of the available AFs were sampled with replacement 1000 times. The model was unable to assess the probability of the presence of an LOH when a small subclonal ratio of tumor cell with *CHD1* loss was coupled with a low cellularity. In such cases the density functions of the heterozygous and homozygous processes could not be distinguished precisely. From the *CHD1*-loss whole genomes, these samples are GP-02, SRR1536760, SRR1534432, TCGA-CH-5750, and TCGA-HC-7737, as illustrated on their respective figures.

For the whole exomes, a 1MB wide surrounding region had to be evaluated due to the restricted number of interrogatable heterozygous SNPs. The resulting predicted probabilities are available in Suppl. Table 2 and 3.

**Suppl.Fig. 25:** Illustrations of the LOH calling procedure on the *CHD1*-loss whole genomes. Each sample has its own figure set: the top panel shows the coverage and spatial distribution of the heterozygous SNPs around *CHD1* in the sample, with a grey box around those that were included in the LOH analysis. The figure also indicates the mean coverage over the entire tumor genome (blue line) and the average coverage in the displayed region (red line). The bottom panel shows the various observed AF-distribution as histograms, and the theoretical distributions that are expected if the deletion is homozygous or heterozygous. The final probabilities that infer whether the observed AF-distribution in the tumor above and around *CHD1* result from an LOH or not are also displayed. The error bars indicate the 95% percentile intervals of 1000 bootstrap iterations.

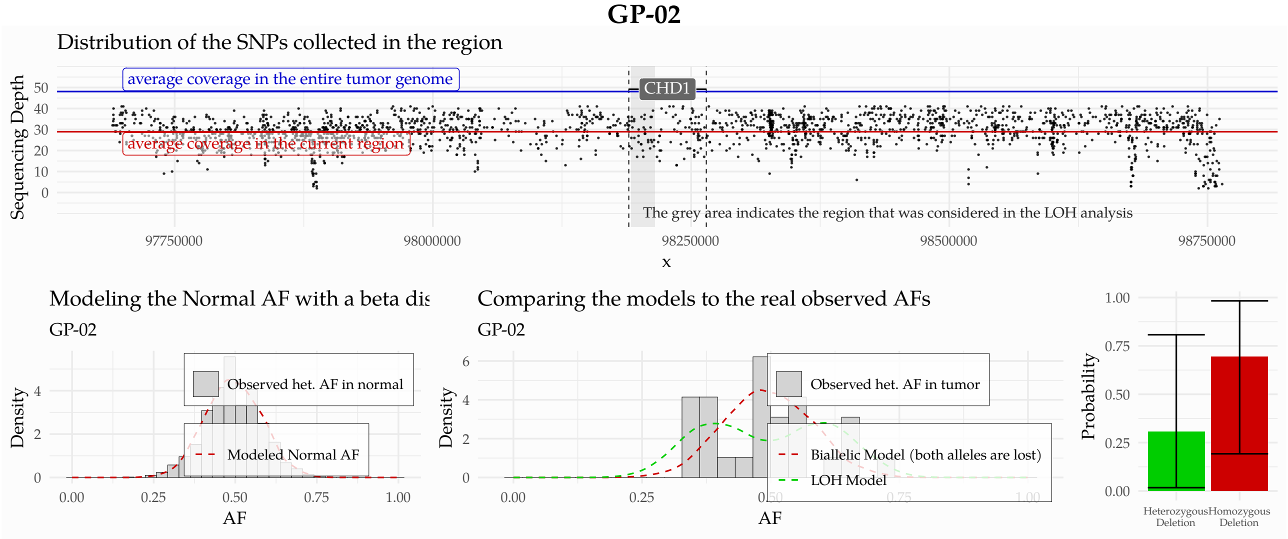

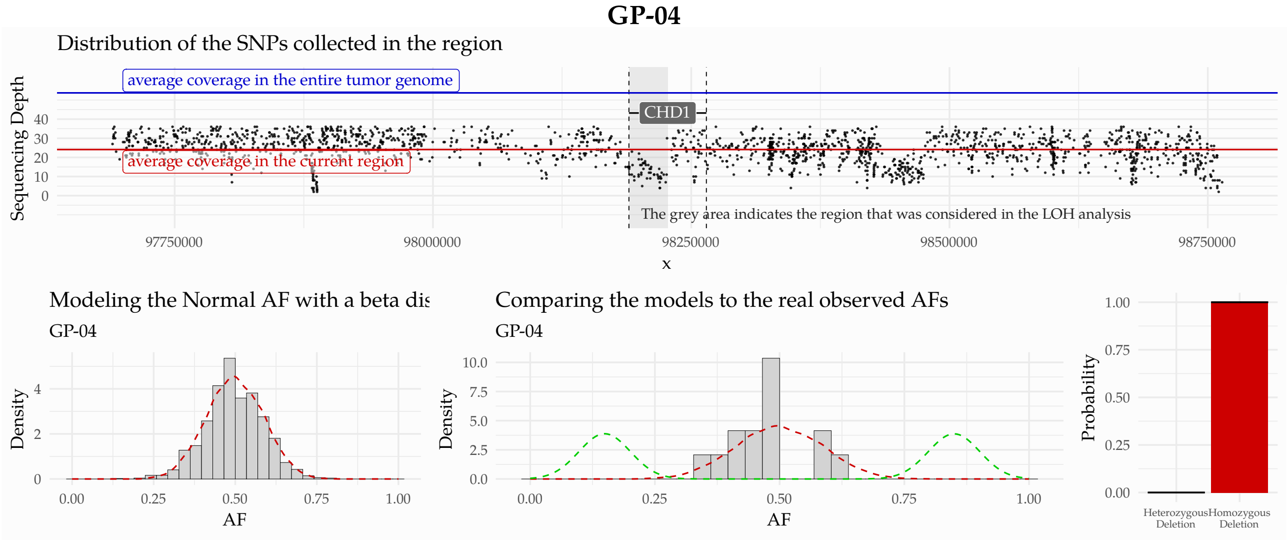

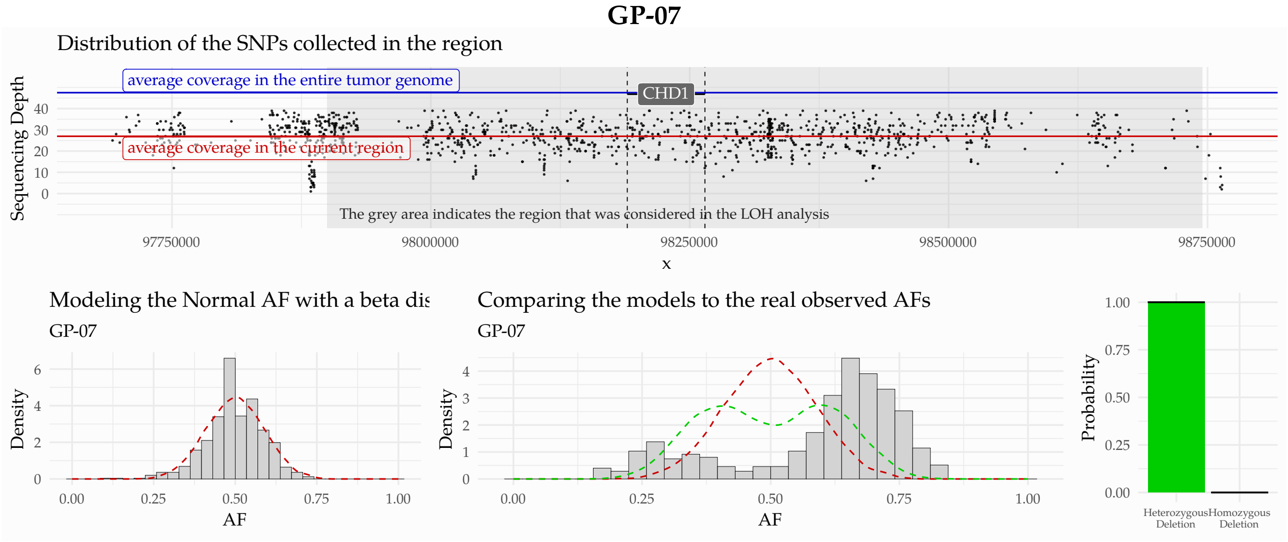
GP-07
Distribution of the SNPs collected in the region
average coverage in the entire tumor genome
average coverage in the current region
CHD1
Sequencing Depth
The grey area indicates the region that was considered in the LOH analysis
97750000
98000000
98250000
98500000
98750000
x
Modeling the Normal AF with a beta dis
GP-07
Comparing the models to the real observed AFs
GP-07
Density
AF
Probability
Heterozygous Deletion
Homozygous Deletion

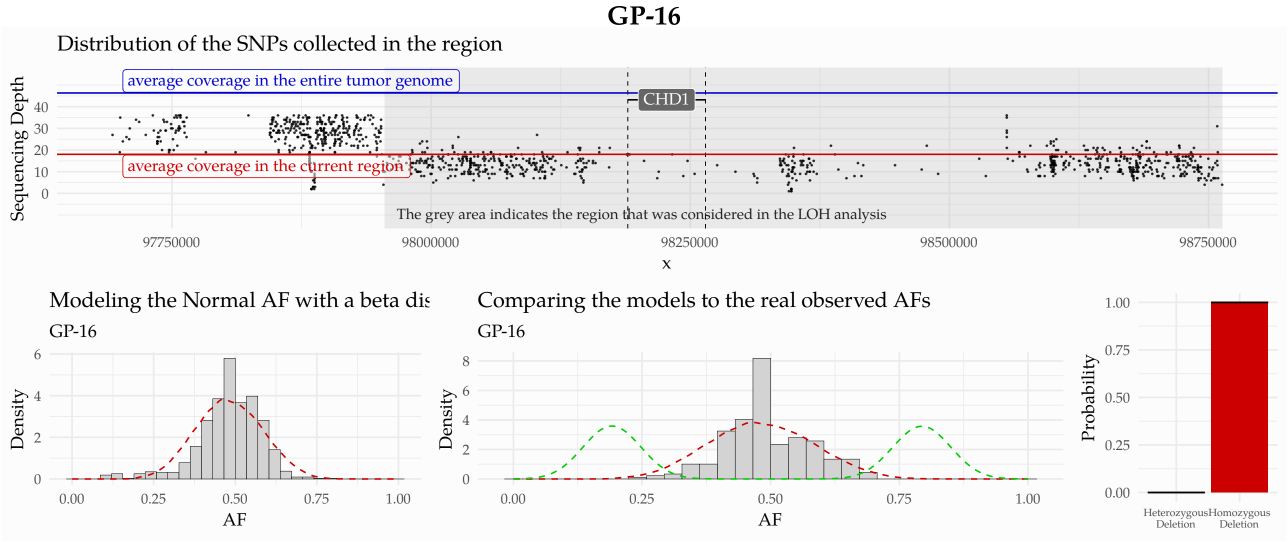
GP-16
Distribution of the SNPs collected in the region
average coverage in the entire tumor genome
average coverage in the current region
CHD1
Sequencing Depth
The grey area indicates the region that was considered in the LOH analysis
97750000
98000000
98250000
98500000
98750000
x
Modeling the Normal AF with a beta dis
GP-16
Comparing the models to the real observed AFs
GP-16
Density
AF
Probability
Heterozygous Deletion
Homozygous Deletion

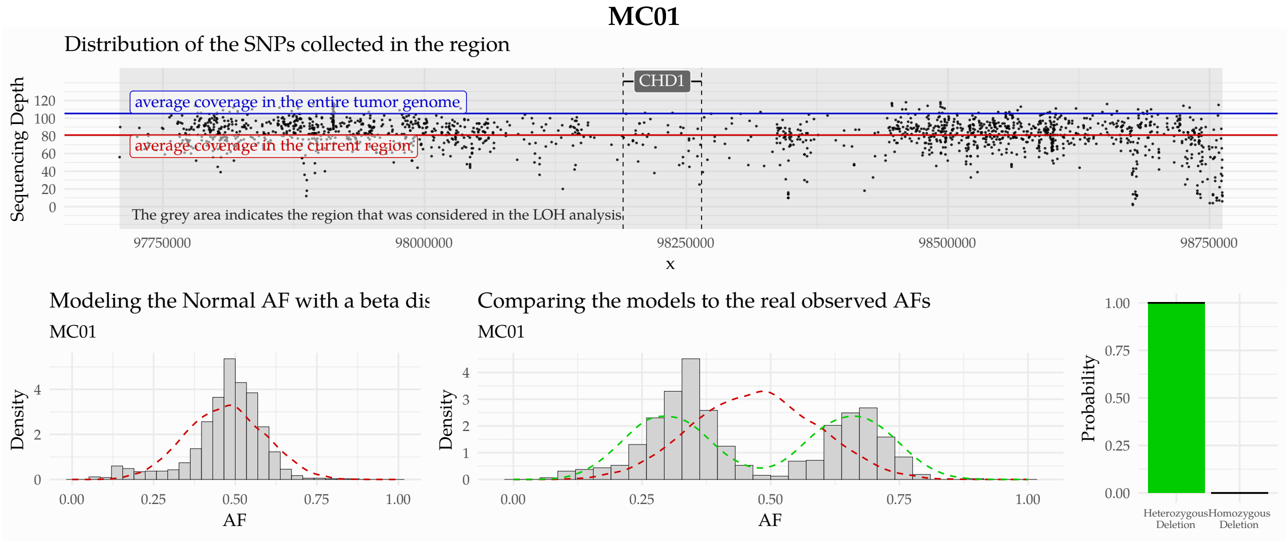
MC01
Distribution of the SNPs collected in the region
average coverage in the entire tumor genome
average coverage in the current region
CHD1
Sequencing Depth
The grey area indicates the region that was considered in the LOH analysis
97750000
98000000
98250000
98500000
98750000
x
Modeling the Normal AF with a beta dis
MC01
Comparing the models to the real observed AFs
MC01
Density
AF
Probability
Heterozygous Deletion
Homozygous Deletion

**MC03**

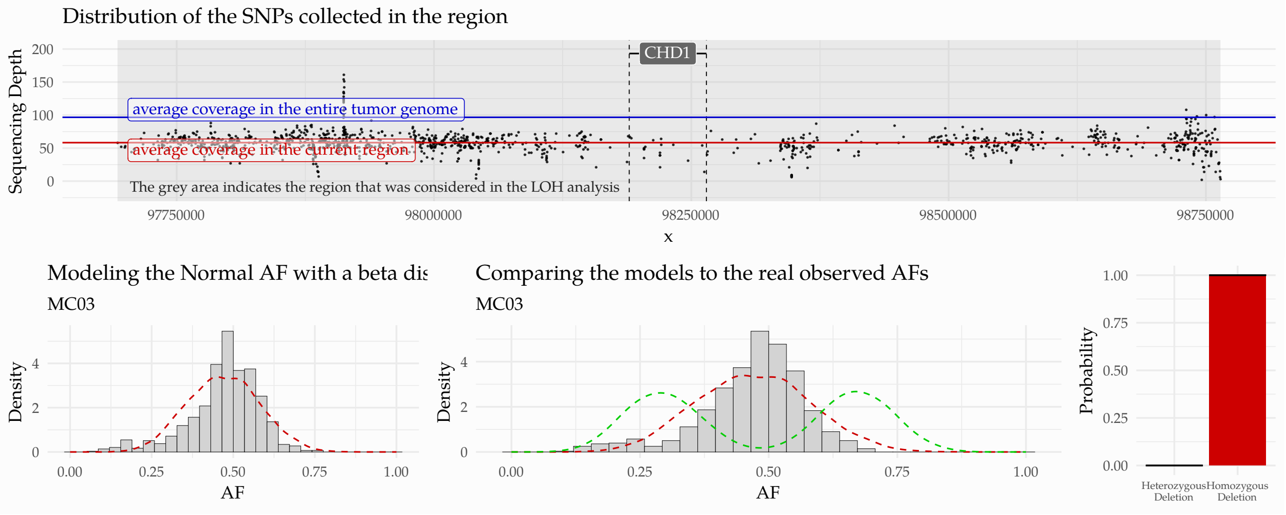

**MC06**

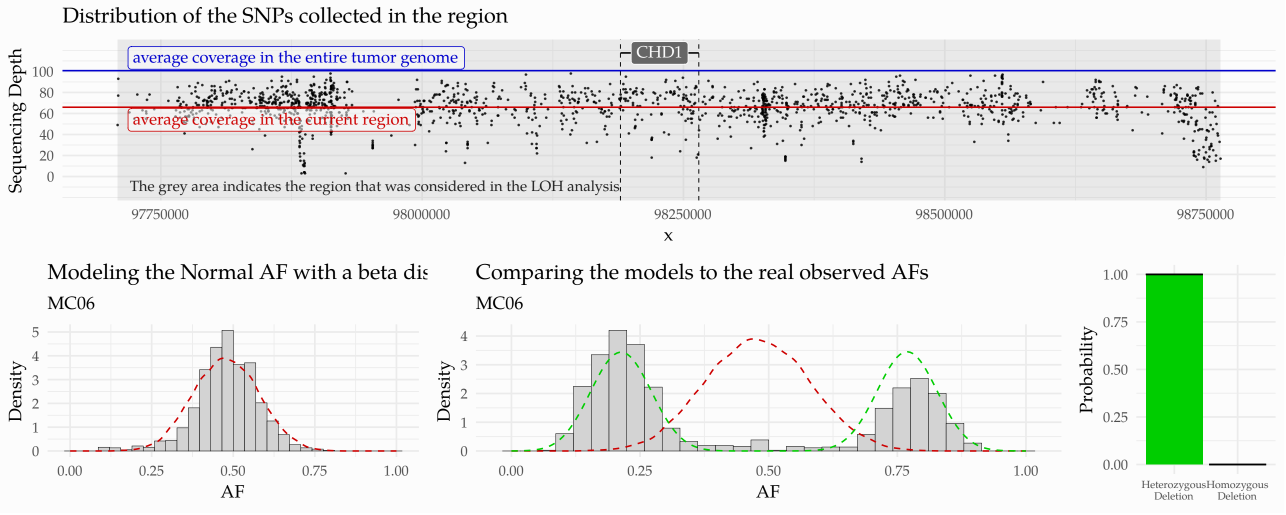

**MC07**

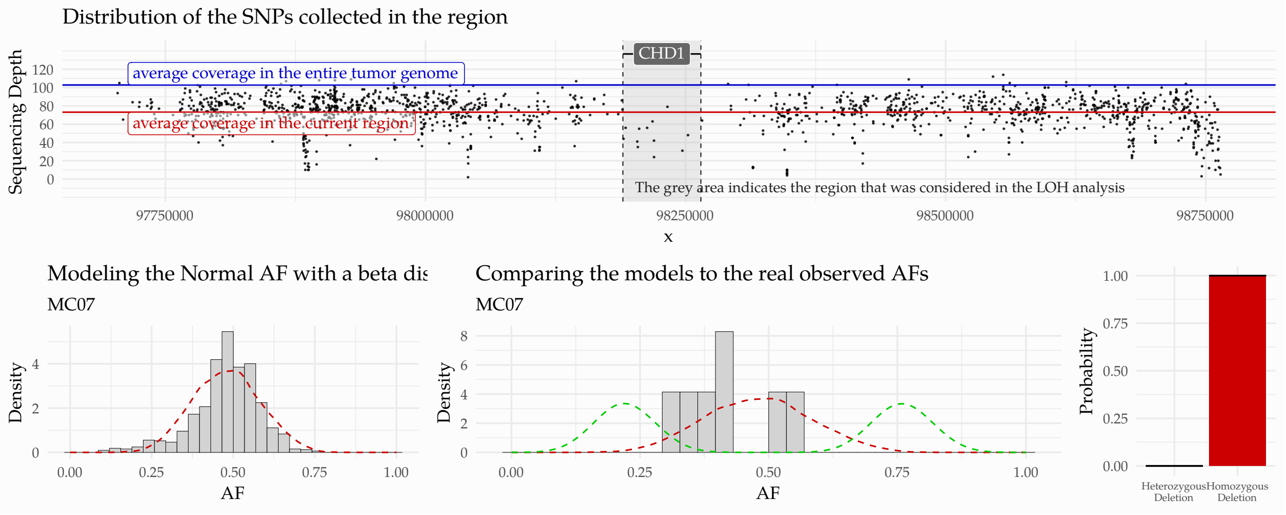

##### SRR1534432

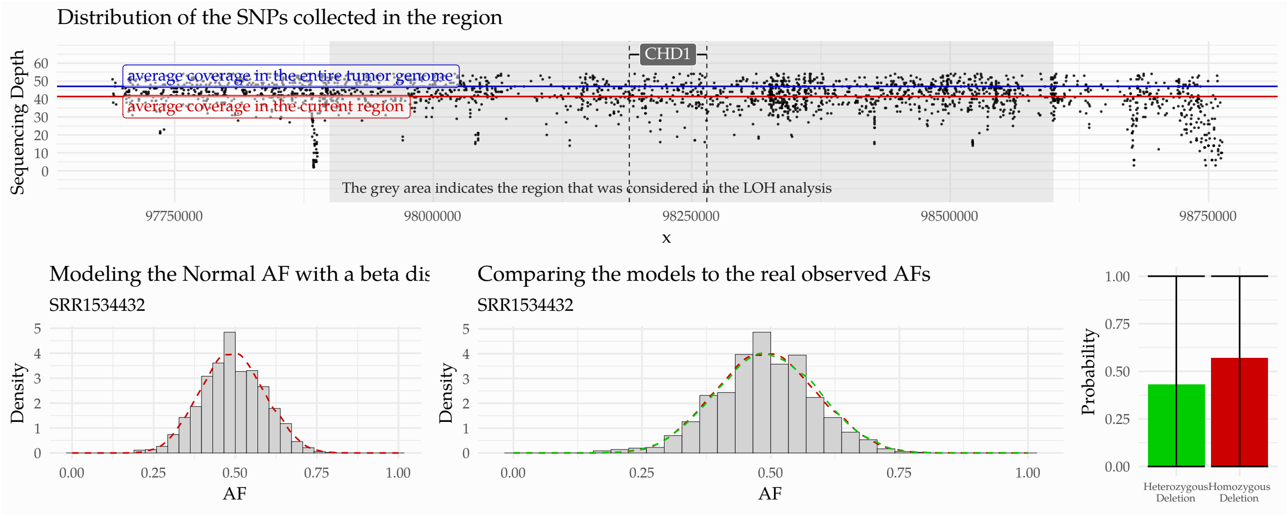

##### SRR1536730

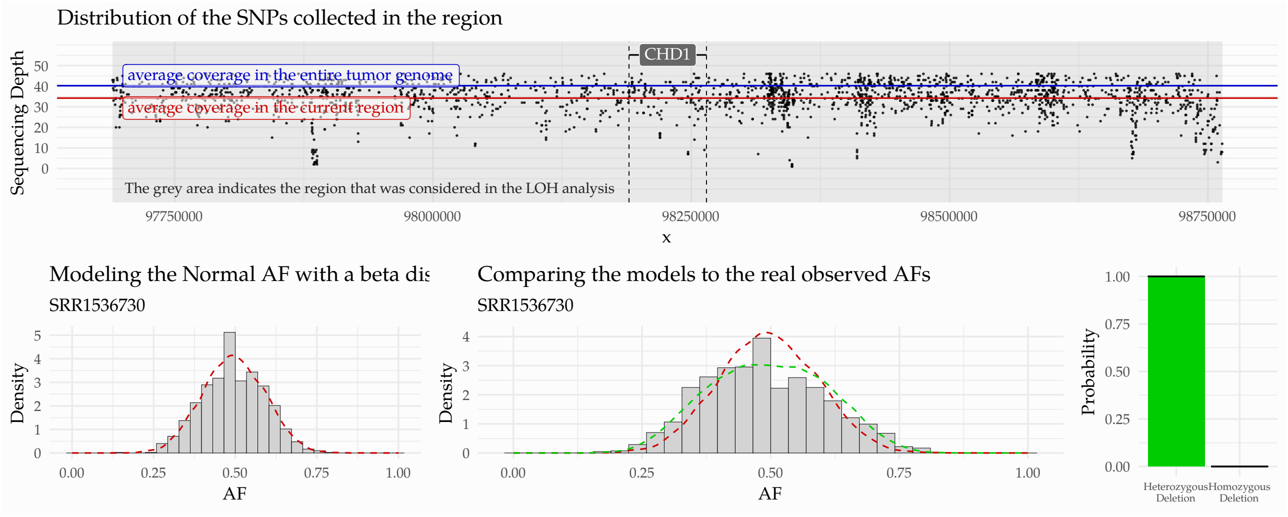

##### SRR1536760

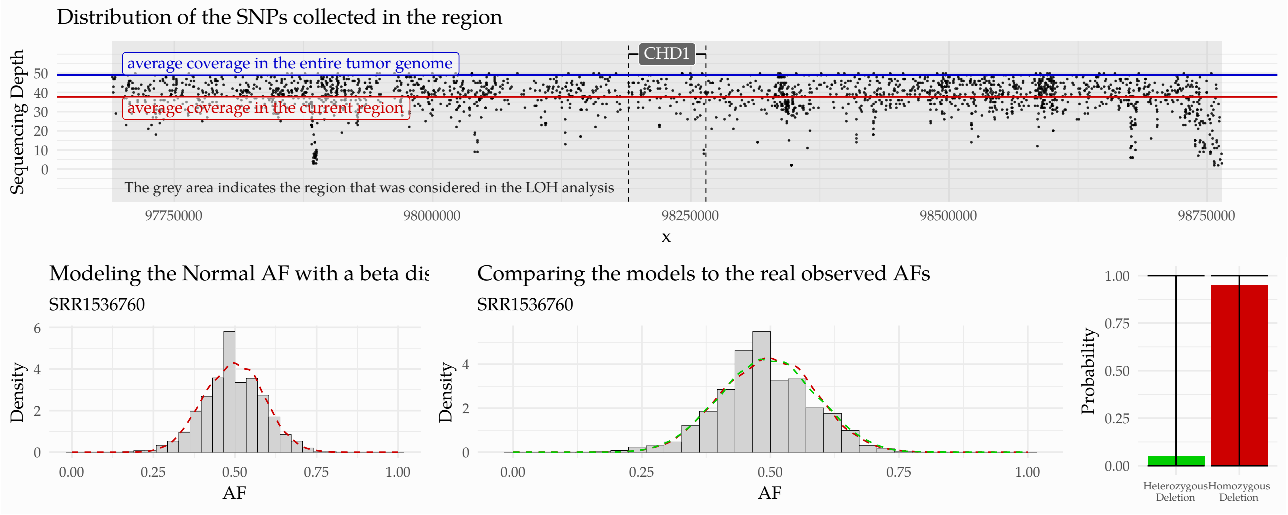

**DO36248**

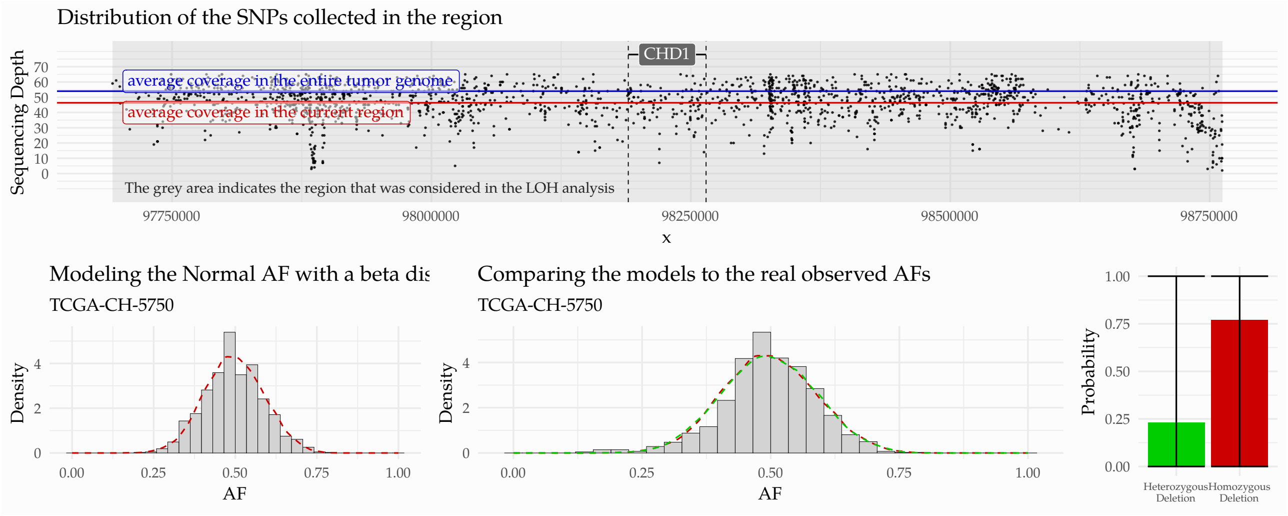

**DO36285**

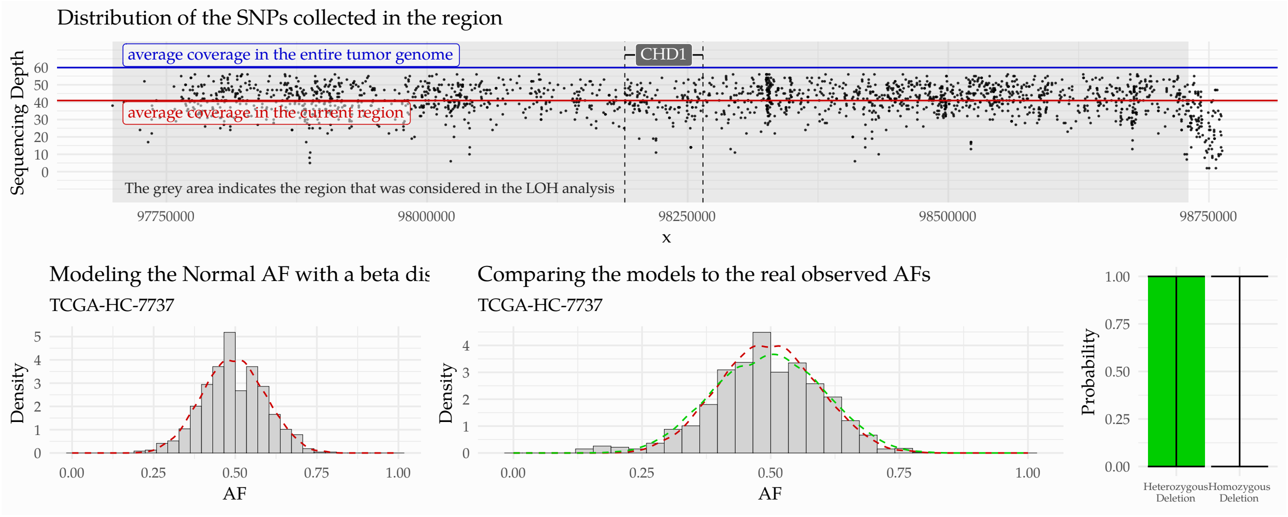

**DO36253**

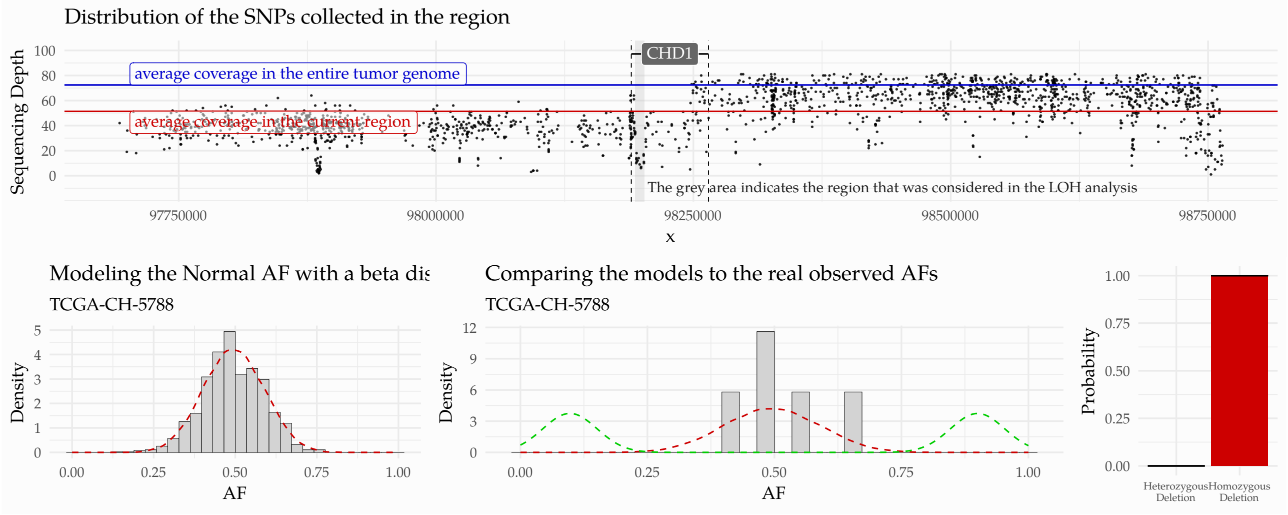

### 9 Genomic Features of the Whole Genomes

Apart of the third generation of somatic signatures, the genomic features described in the article were all determined in our earlier publication: Sztupinszki et al. 2020 [12]. All the values are available in the supplementary materials of the article.

#### 9.1 Genomic Scars

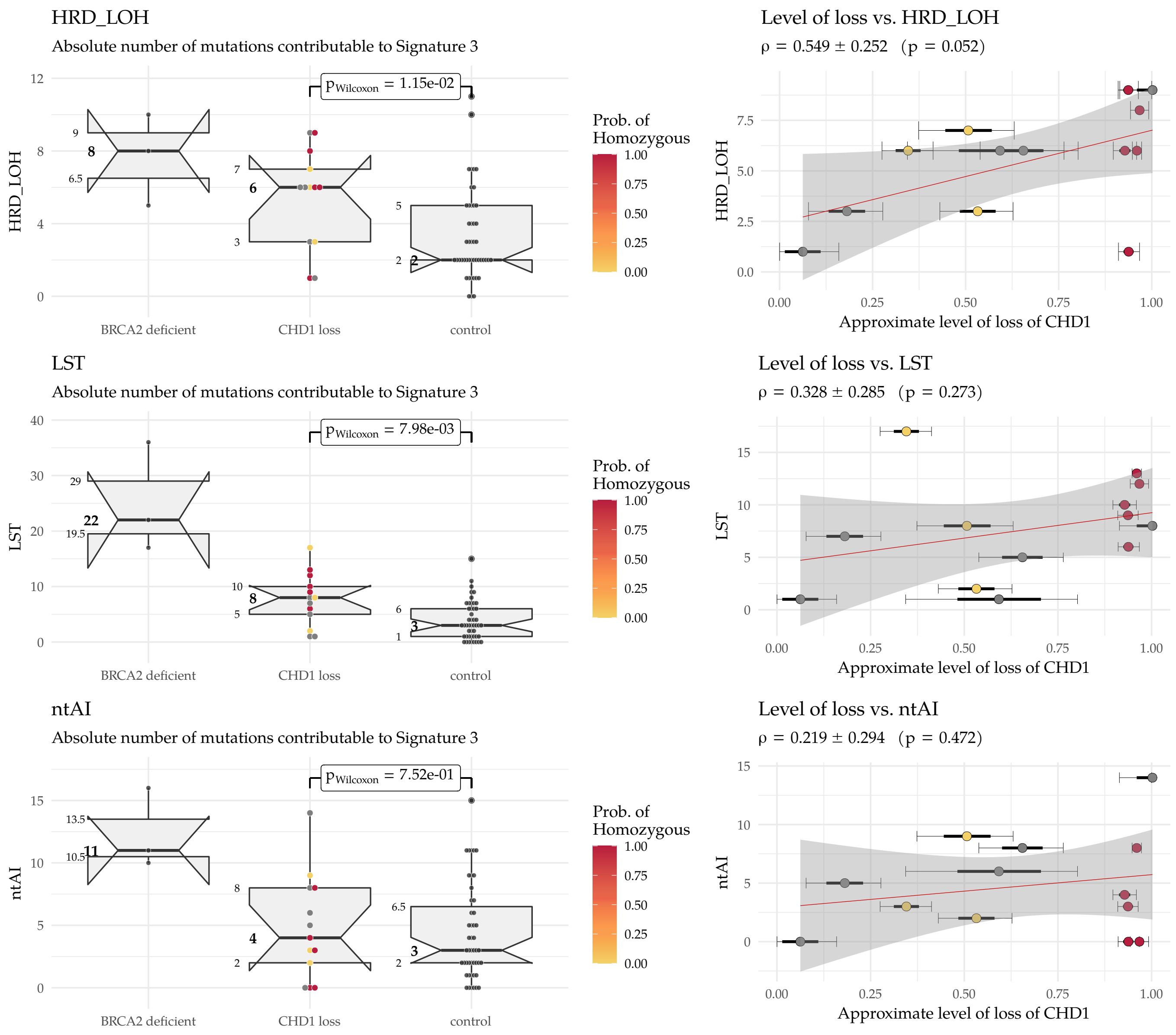

**Suppl.Fig. 26:** HRD-related genomic scars in the whole genomes. **HRD_LOH**: HRD Loss of Heterozygosity, **LST**: Large Scale Transition, **ntAI**: number of telomeric Allelic Imbalance

#### 9.2 Somatic Signatures

Since the publication of the third generation of somatic signatures [13], it has become standard to use the new set to describe the underlying mutational processes in each tumor sample. The HRDetect model[14], however, was developed on the second generation of these signatures [15], therefore they cannot be abandoned completely. In the following, both the second and third generation single nucleotide signatures will be discussed, and the extraction of the third generation signatures will be explained as well.

The single nucleotide and indel spectra were generated using the ICAMs R package [16] (Suppl.Fig. **??**). In order to minimize the signal to noise ratio, only those signatures were considered that have significantly improved the reconstruction similarities (cosine-similarity). This was realized by adopting a dynamic signature extraction technique, in which originally we only considered the HRD-related signatures (e.g. Signature.3 and Signature.8 for the second generation signatures) [1] as core contributors, and every additional signature was analyzed one by one, throughout multiple iterations. In each iteration only one of the signatures was appended to the list of core signatures, the one that maximized the mean reconstruction similarity between the original spectras and their reconstructions. Such iterations were executed, until the increment in the similarity fall below 0.001.

##### 9.2.1 Second-generation Signatures

**Suppl.Fig. 27:** Number of variants contributable ot the 2nd generation single nucleotide signatures. Signatures 1,2,3,5,6,8 and the total number of SNVs in the sample (numSNV)

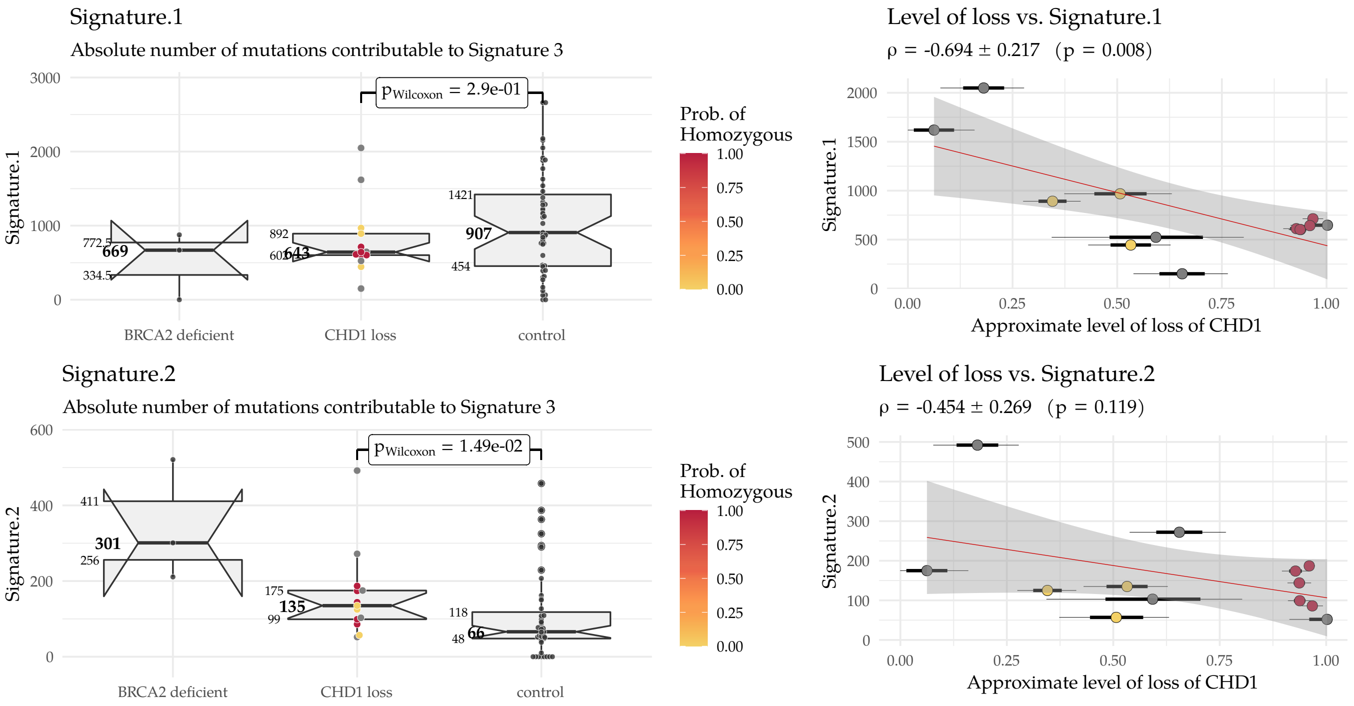

[1] Forcing the model to consider Signature.3 and Signature.8 does not necessarily mean, than the final contributions of these signatures will be non-zero at the end of the dynamic extraction process. If an additional signature can better represent the observed mutational patterns, it can simply set the contribution of these signatures to zero. (In fact, the contribution of Signature.8 ended up zero.)

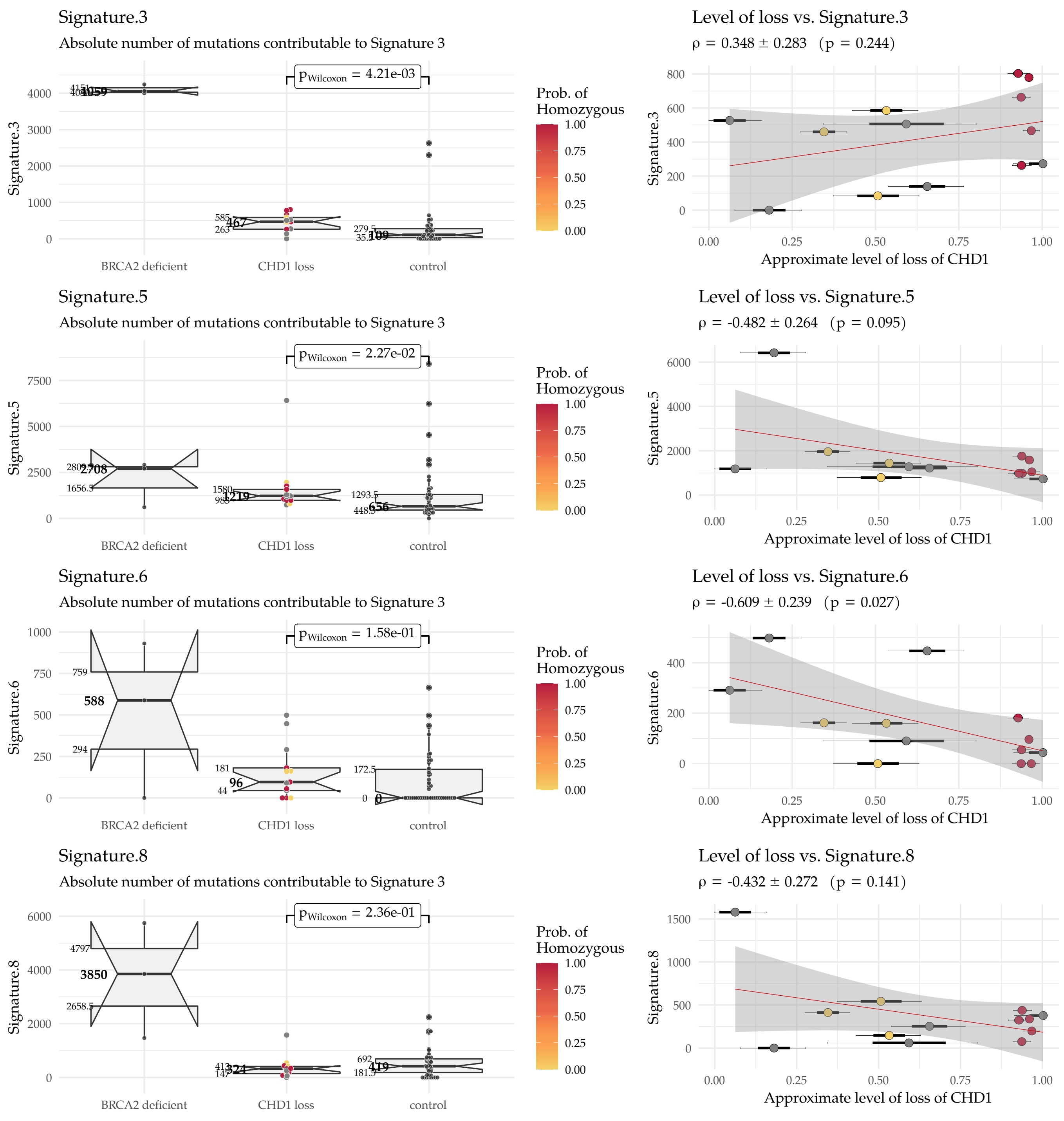

##### 9.2.2 Third-generation Signatures

The third-generation 96-dimensional mutational catalogs (a.k.a spectra) of the single base substitutions were collected into the columns of matrix **M** using the `ICAMs` R package [16], from the somatic vcfs returned by `Mutect2` (GATK v4.1.0) [8]. The **P** matrix of SBS-signatures were downloaded from the following link:

- https://www.synapse.org/#!Synapse:syn12009743

The two matrices were used in a non-negative least-squares problem to estimate the matrix of exposures **E**:

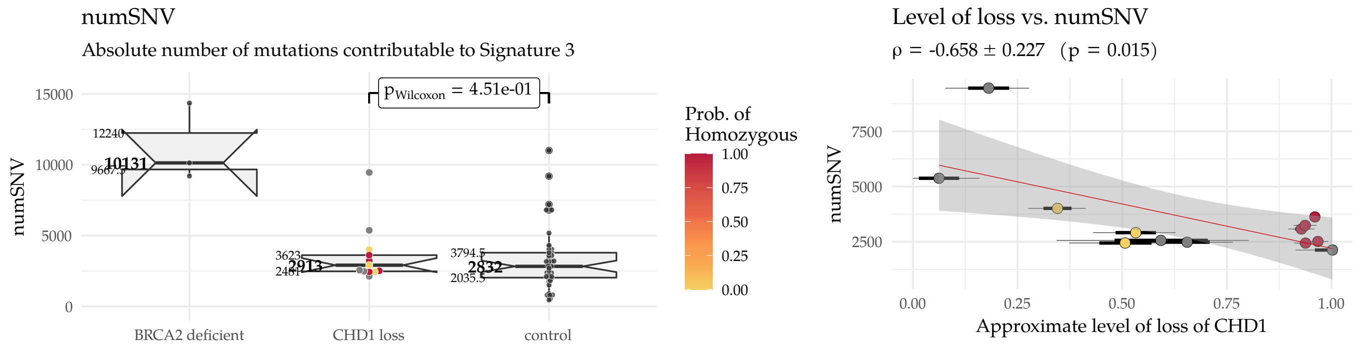

$$\min_{\mathbf{E}_i} ||\mathbf{P}\mathbf{E}_i - \mathbf{M}_i||^2, \quad ,\text{subject to} \quad \mathbf{E}_i \geq 0, \text{ for all } i = 1, \dots, N,$$

where $i$ is a given sample. The similarity between the reconstructed and original mutational was evaluated by using the cosine similarity between the two (Suppl.Fig. 28).

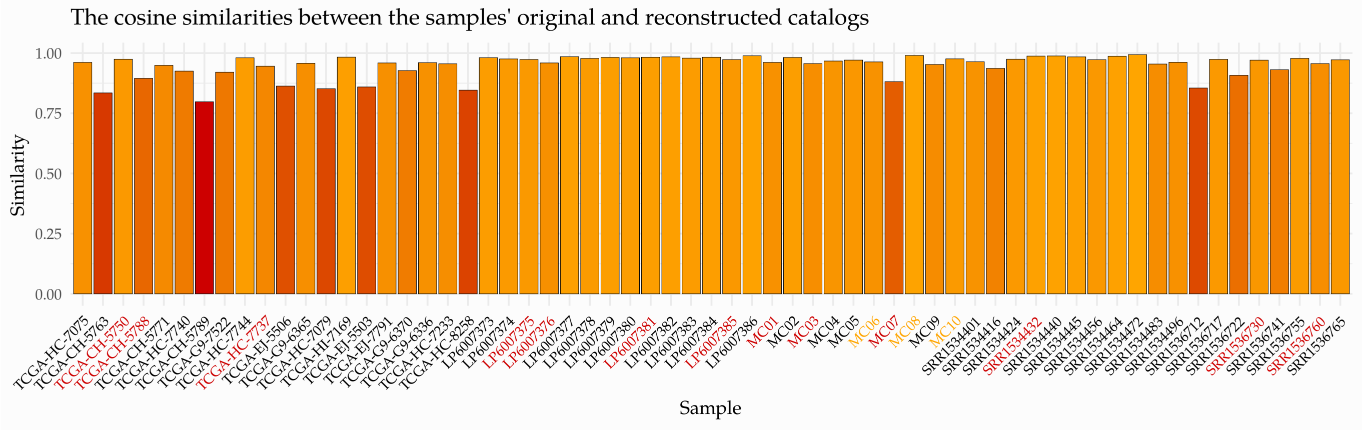

**Suppl.Fig. 28:** The cosine similarity between the original and reconstructed mutational SBS spectra in the whole genomes.

The signature composition of the whole genomes is displayed in Suppl.Fig.

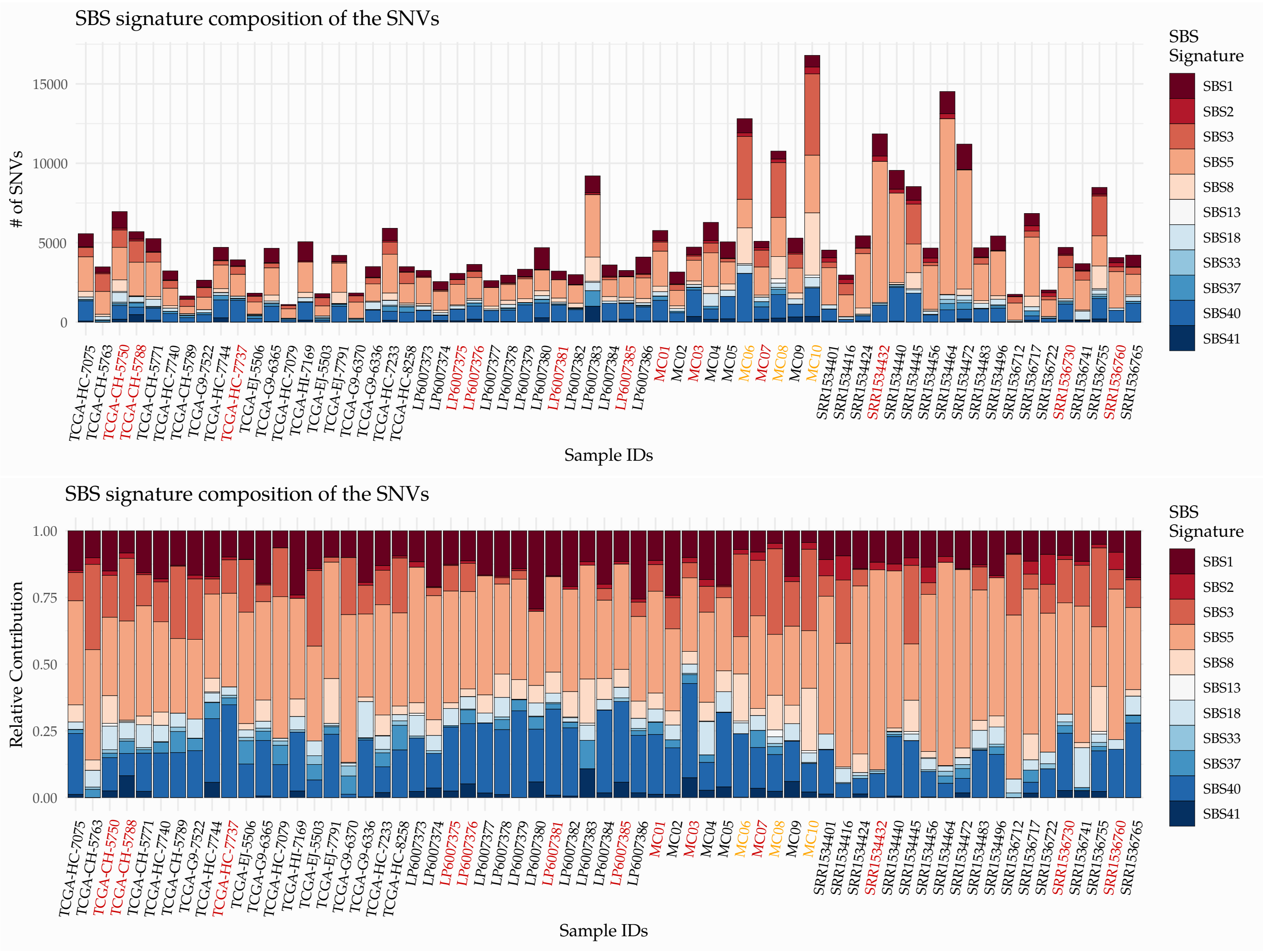

**Suppl.Fig. 29:** The SBS signature compositon of the whole genomes. Above are the absolute numbers, below the relative ratios.

**Suppl.Fig. 30:** Third generation single nucleotide (SBS) signatures.

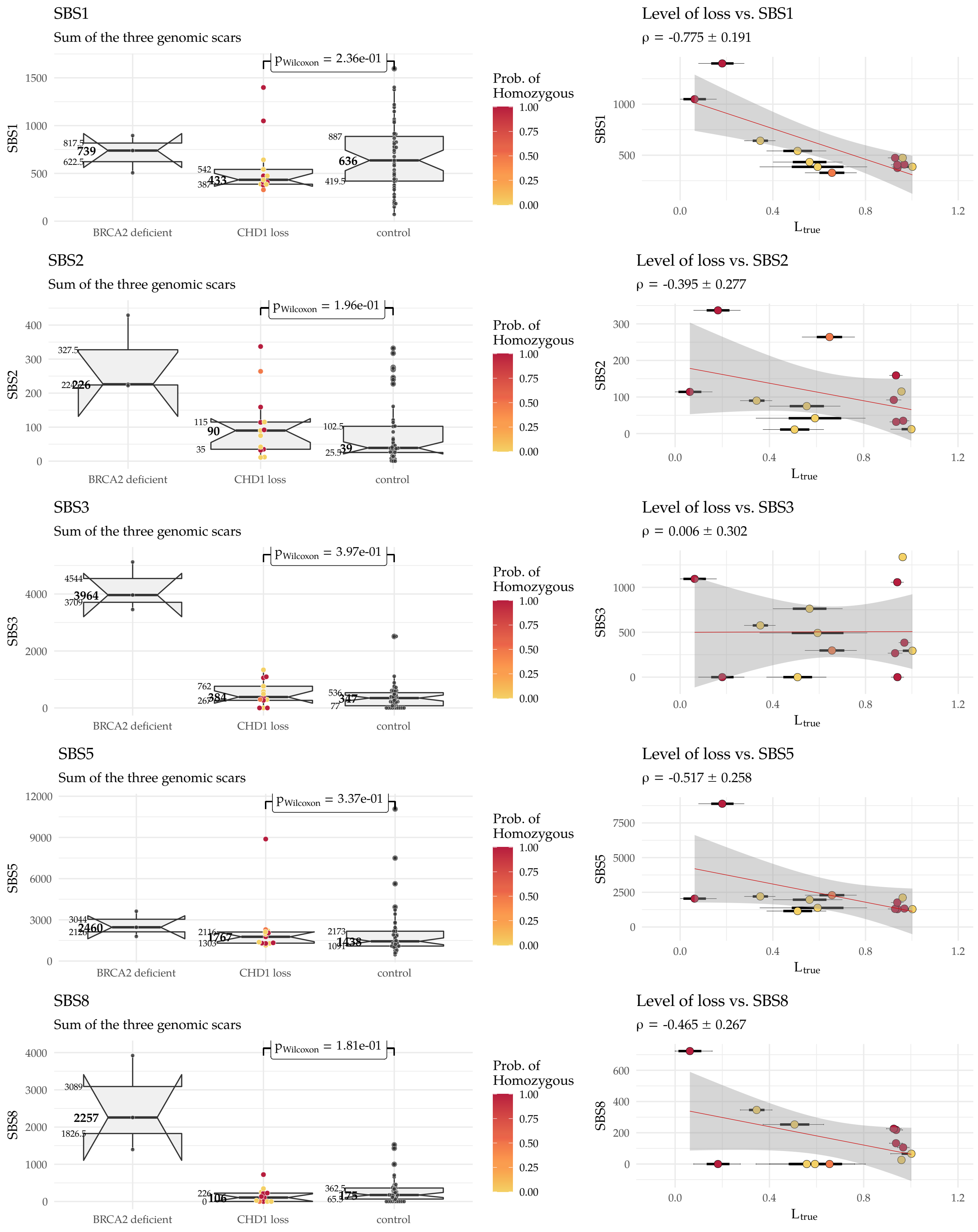

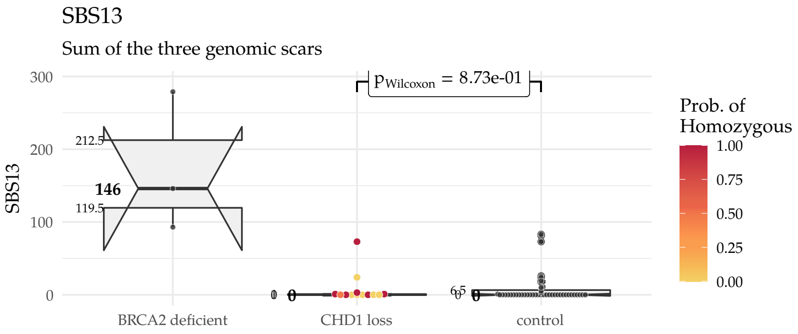
SBS13
Sum of the three genomic scars
$p_{Wilcoxon}$ = 8.73e-01
SBS13
BRCA2 deficient
CHD1 loss
control
Prob. of Homozygous

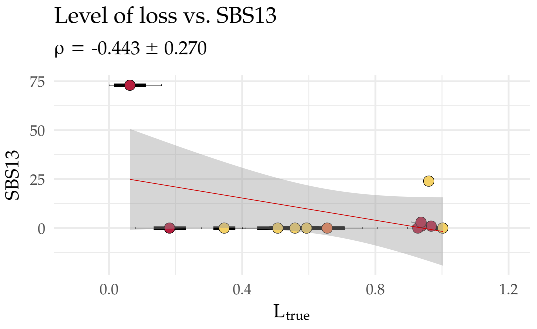
Level of loss vs. SBS13
ρ = -0.443 ± 0.270
SBS13
$L_{true}$

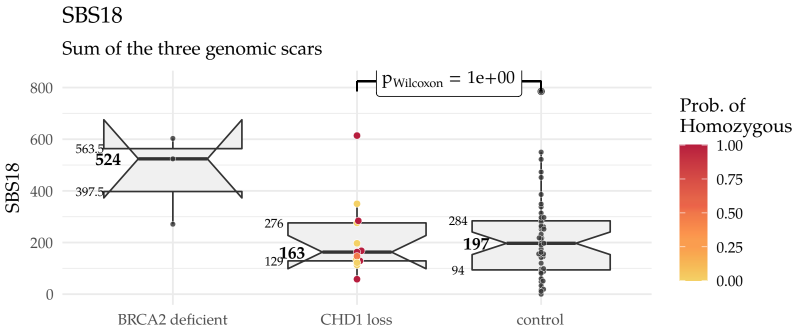
SBS18
Sum of the three genomic scars
$p_{Wilcoxon}$ = 1e+00
SBS18
BRCA2 deficient
CHD1 loss
control
Prob. of Homozygous

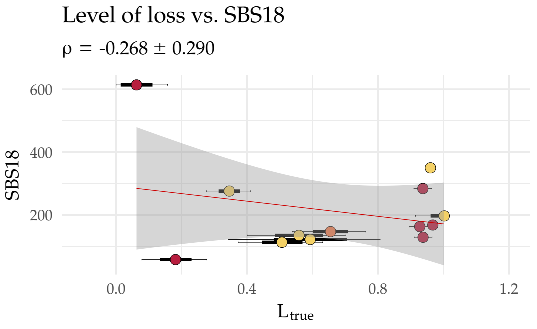
Level of loss vs. SBS18
ρ = -0.268 ± 0.290
SBS18
$L_{true}$

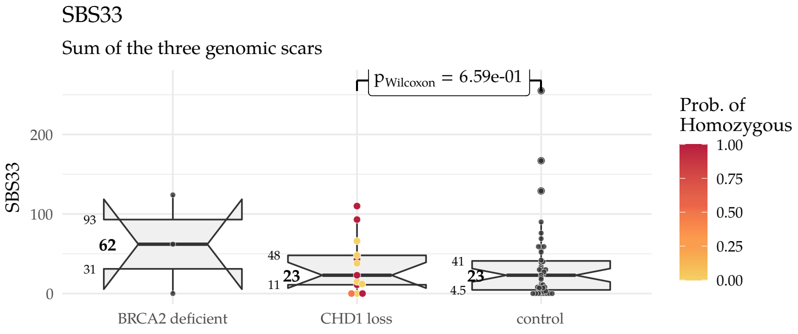
SBS33
Sum of the three genomic scars
$p_{Wilcoxon}$ = 6.59e-01
SBS33
BRCA2 deficient
CHD1 loss
control
Prob. of Homozygous

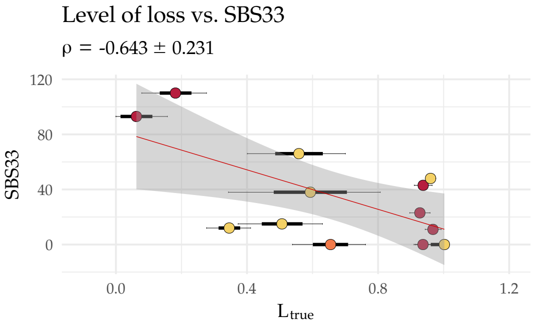
Level of loss vs. SBS33
ρ = -0.643 ± 0.231
SBS33
$L_{true}$

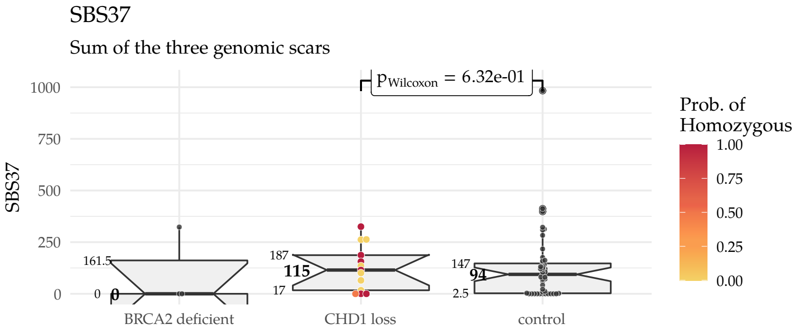
SBS37
Sum of the three genomic scars
$p_{Wilcoxon}$ = 6.32e-01
SBS37
BRCA2 deficient
CHD1 loss
control
Prob. of Homozygous

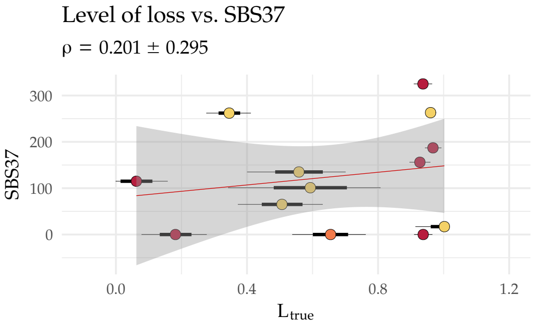
Level of loss vs. SBS37
ρ = 0.201 ± 0.295
SBS37
$L_{true}$

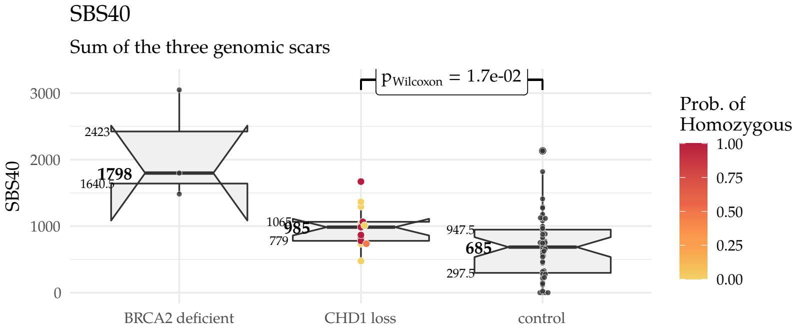
SBS40
Sum of the three genomic scars
$p_{Wilcoxon}$ = 1.7e-02
SBS40
BRCA2 deficient
CHD1 loss
control
Prob. of Homozygous

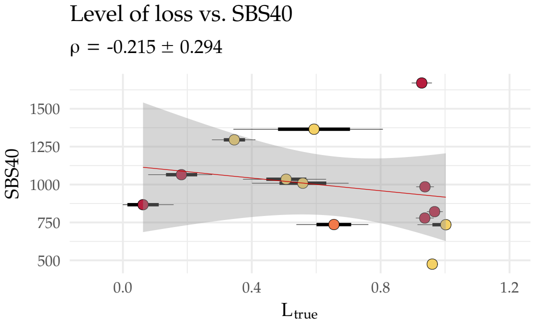
Level of loss vs. SBS40
ρ = -0.215 ± 0.294
SBS40
$L_{true}$

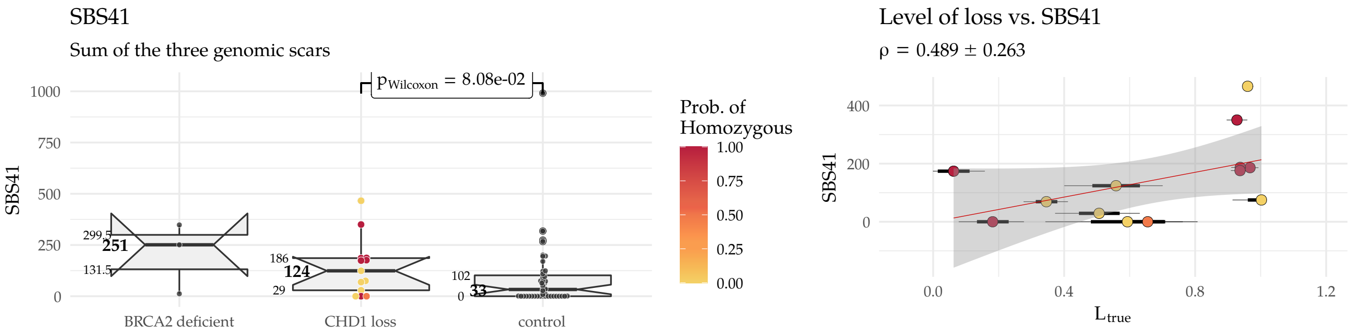

##### 9.2.3 3rd generation indel (ID) signatures

The controbutions of the 3rd generation indel signatures were identified by following the pipeline discussed in the previous subsection.

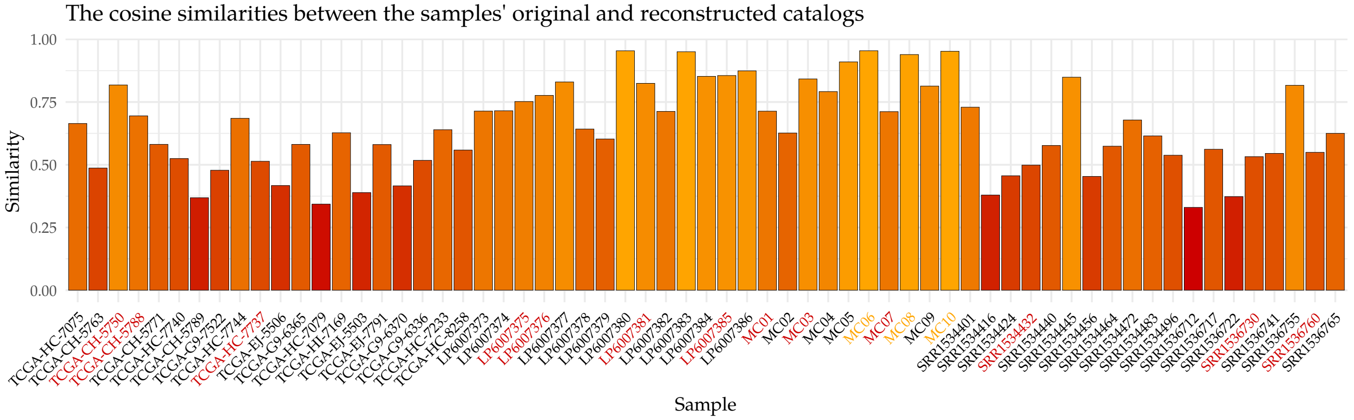

**Suppl.Fig. 31:** ↑ The cosine similarity between the original and reconstructed mutational ID spectra in the whole genomes.

**Suppl.Fig. 32:** ↓The ID signature compositon of the whole genomes. Above are the absolute numbers, below the relative ratios.

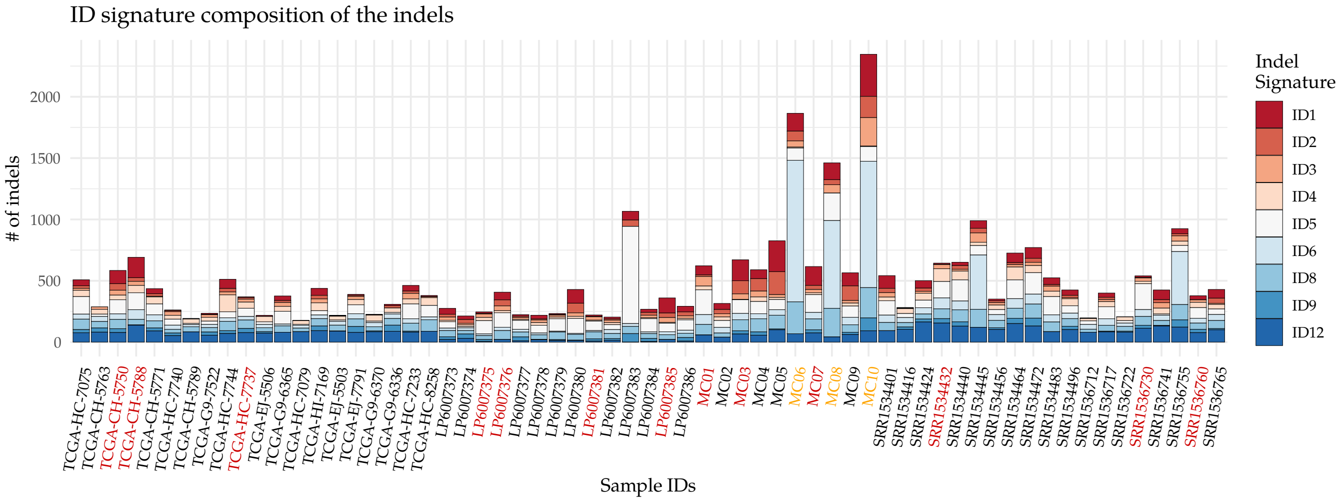

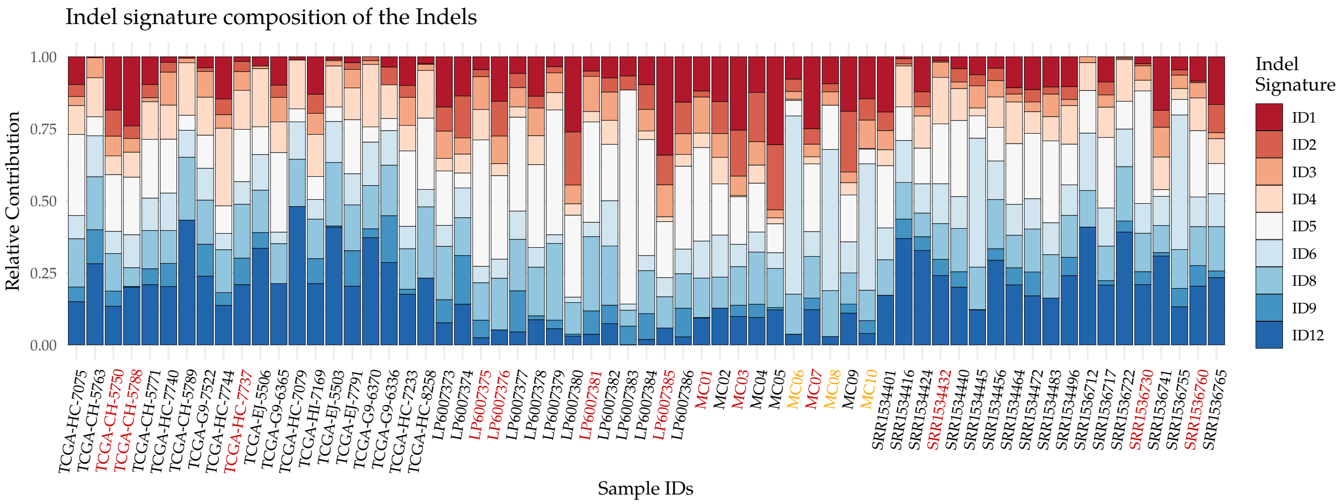

**Suppl.Fig. 33:** ↓ Number of variants contributable to the the third generation indel (ID) signatures.

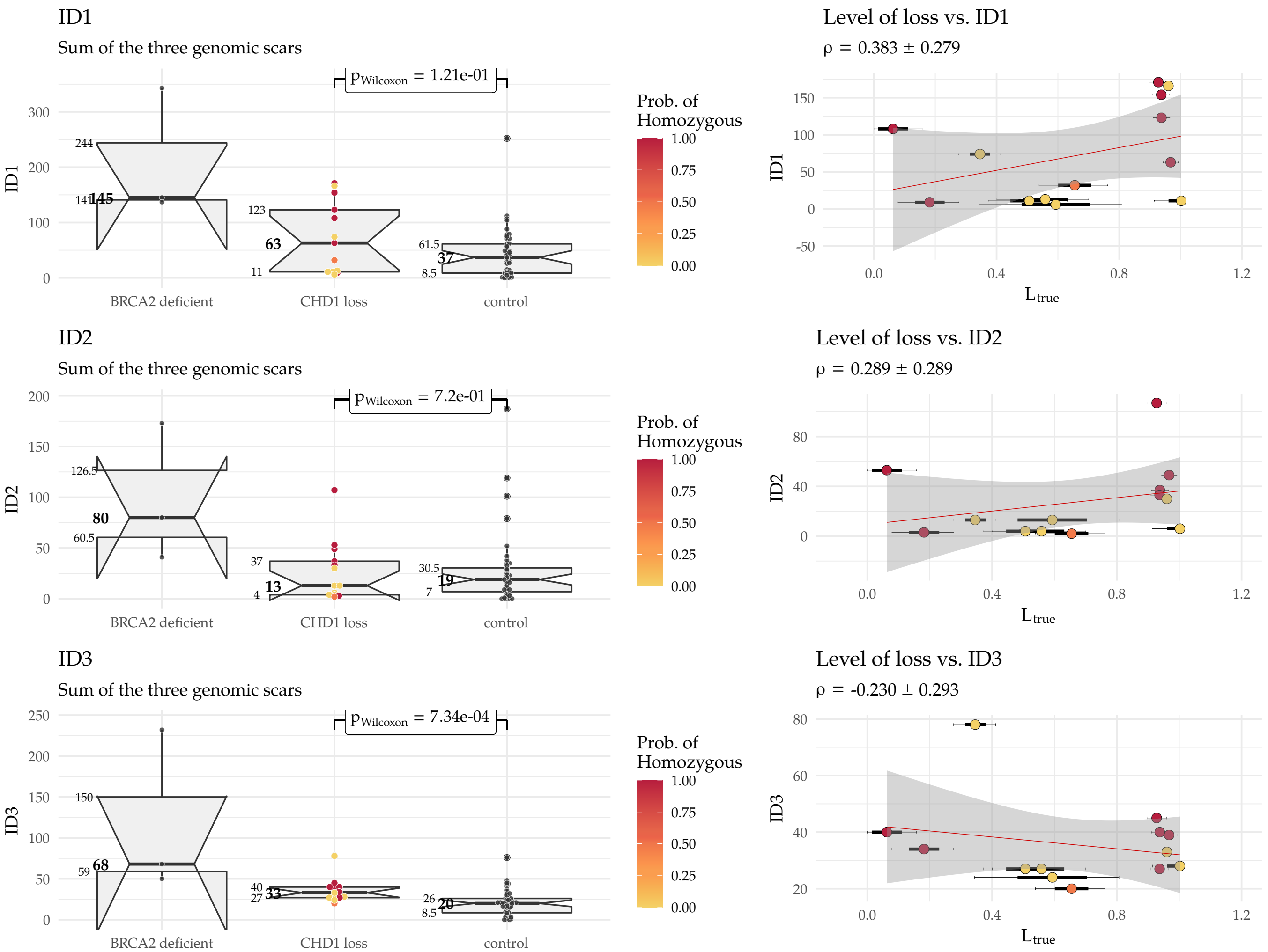

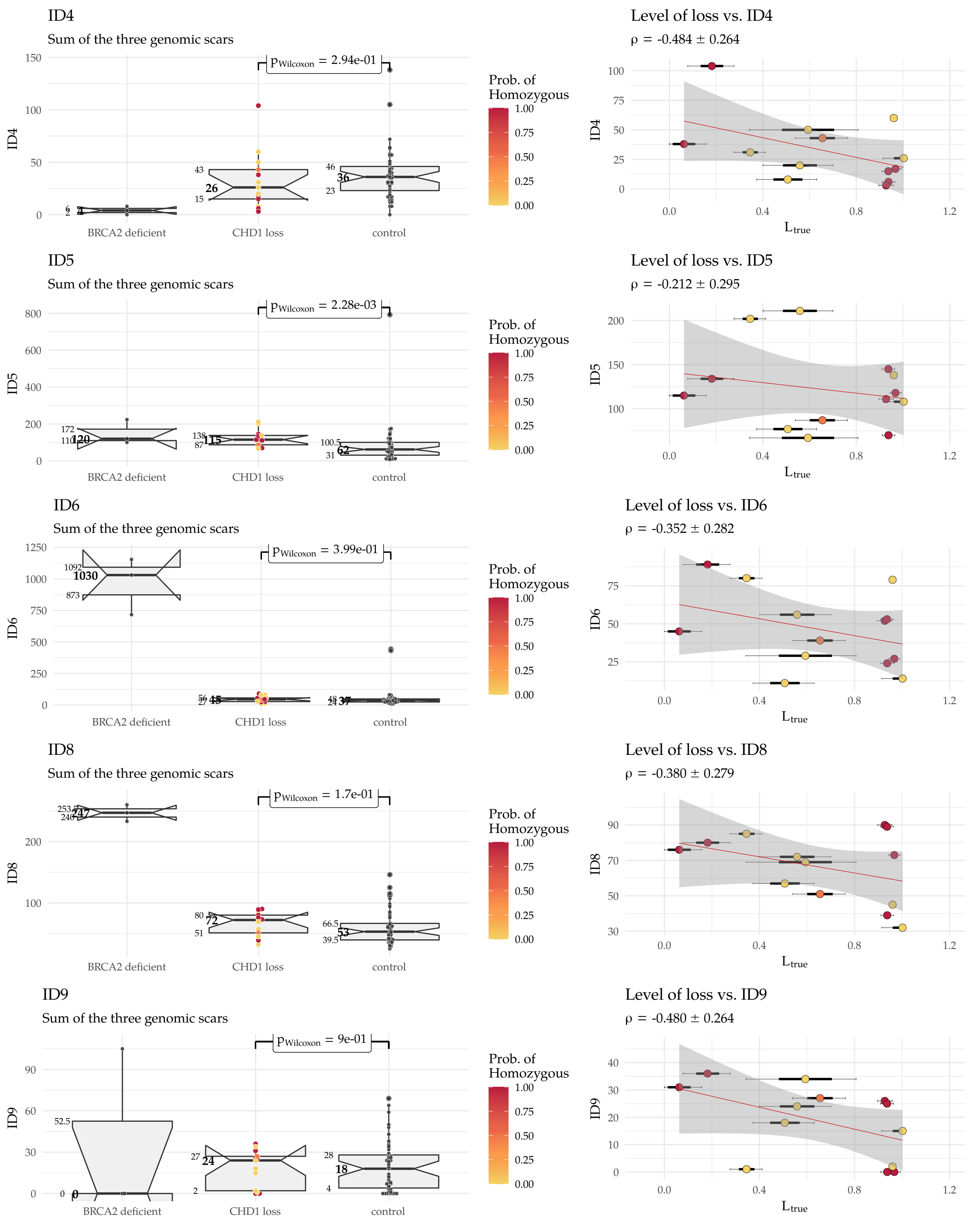
ID4
Sum of the three genomic scars
$p_{Wilcoxon}$ = 2.94e-01
Prob. of Homozygous
BRCA2 deficient
CHD1 loss
control
Level of loss vs. ID4
ρ = -0.484 ± 0.264
$L_{true}$
ID5
Sum of the three genomic scars
$p_{Wilcoxon}$ = 2.28e-03
Level of loss vs. ID5
ρ = -0.212 ± 0.295
ID6
Sum of the three genomic scars
$p_{Wilcoxon}$ = 3.99e-01
Level of loss vs. ID6
ρ = -0.352 ± 0.282
ID8
Sum of the three genomic scars
$p_{Wilcoxon}$ = 1.7e-01
Level of loss vs. ID8
ρ = -0.380 ± 0.279
ID9
Sum of the three genomic scars
$p_{Wilcoxon}$ = 9e-01
Level of loss vs. ID9
ρ = -0.480 ± 0.264

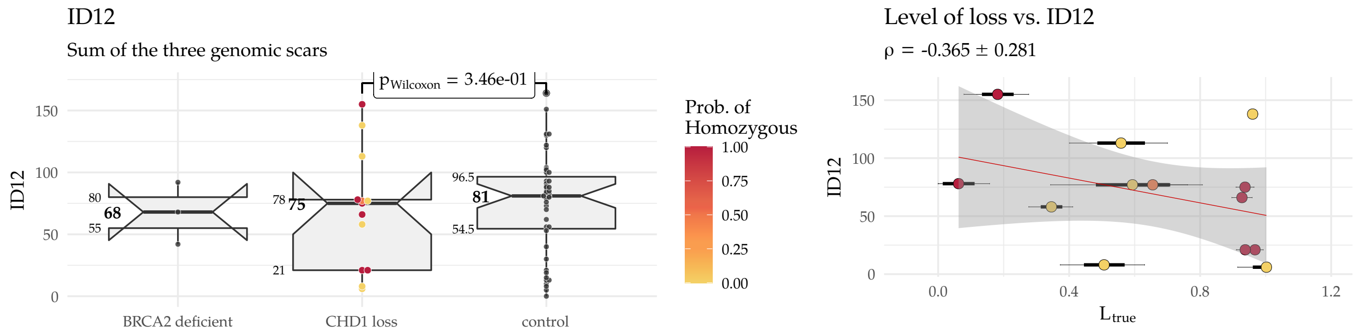

#### 9.3 Deletions - traditional approach

The "traditional approach" of deletions simply distinguishes **microhomology-mediated deletions**, **simple repeats**, and **deletions with no microhomology**, a.k.a. **unique deletions**, by following the definitions below:

- **Simple Repeats:** When the entire deletion is repeated after or before the deletion.
- **Microhomology-mediated deletion of size n:** When only the first $n$ bases are repeated after the deletion or the last $n$ nucleotides preceed it directly.
- **Unique deletion:** When the sequence of the deletion has no resemblance to the sequences that directly follow or preceed it.

**Suppl.Fig. 34:** The "traditional" classification of deletions. ndel: number of deletions, del_ins_ratio: deletion insertion ratio, microhomology: number of microhomology mediated deletions, microhomology_del_ratio: ratio of microhomology mediated deletions relative to the number of deletions

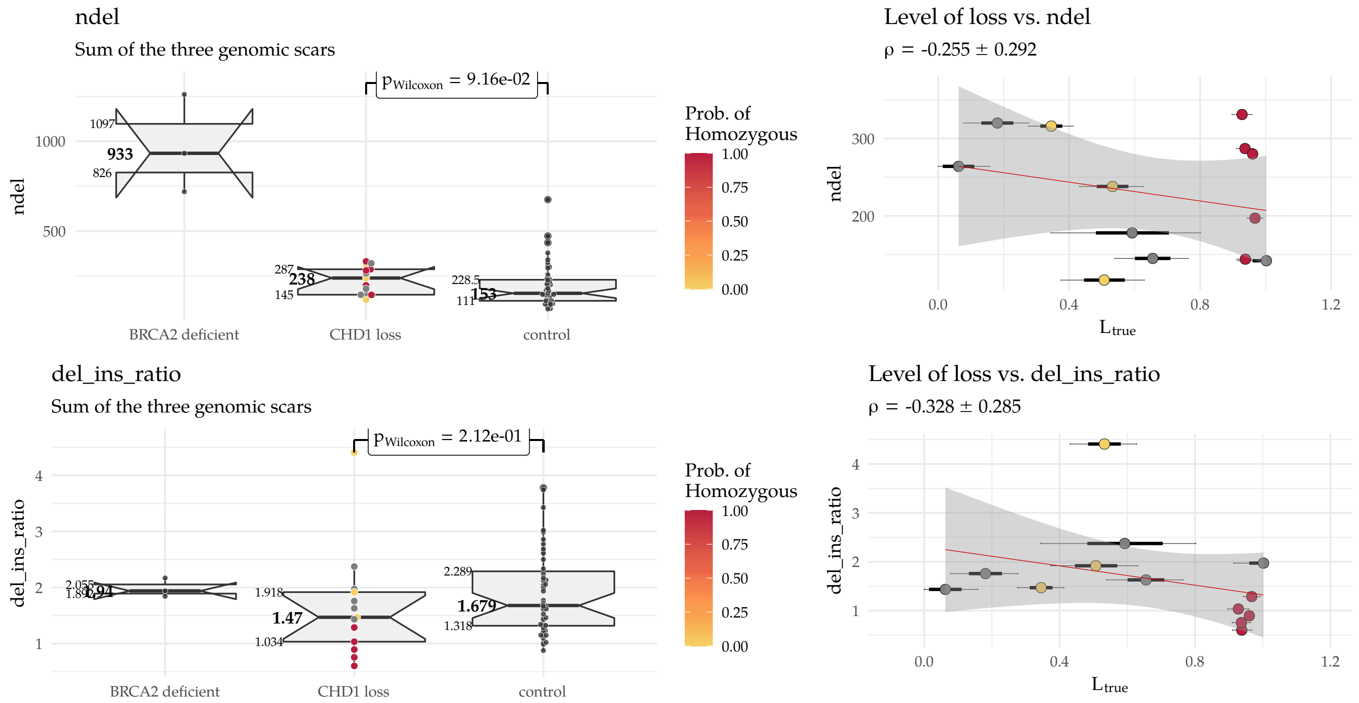

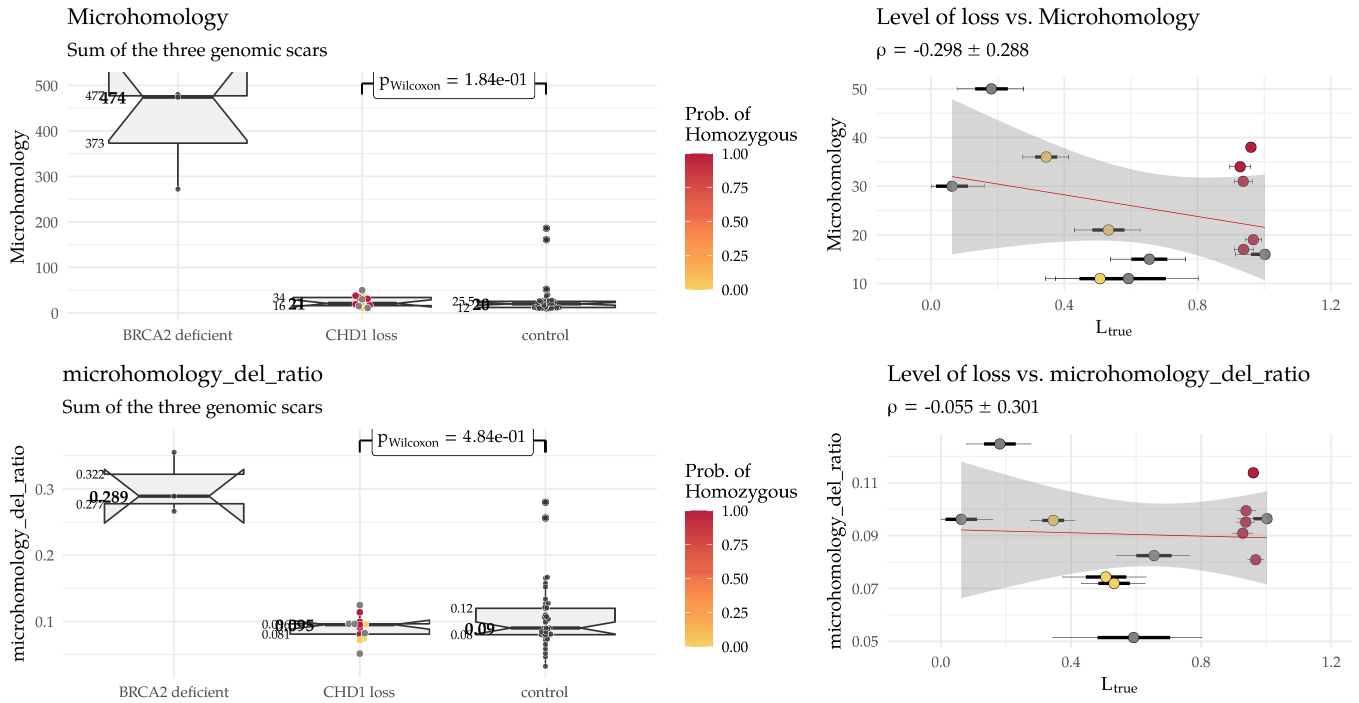

#### 9.4 Rearrangement Signatures

**Suppl.Fig. 35:** The six structural-variant-based rearrangement signatures

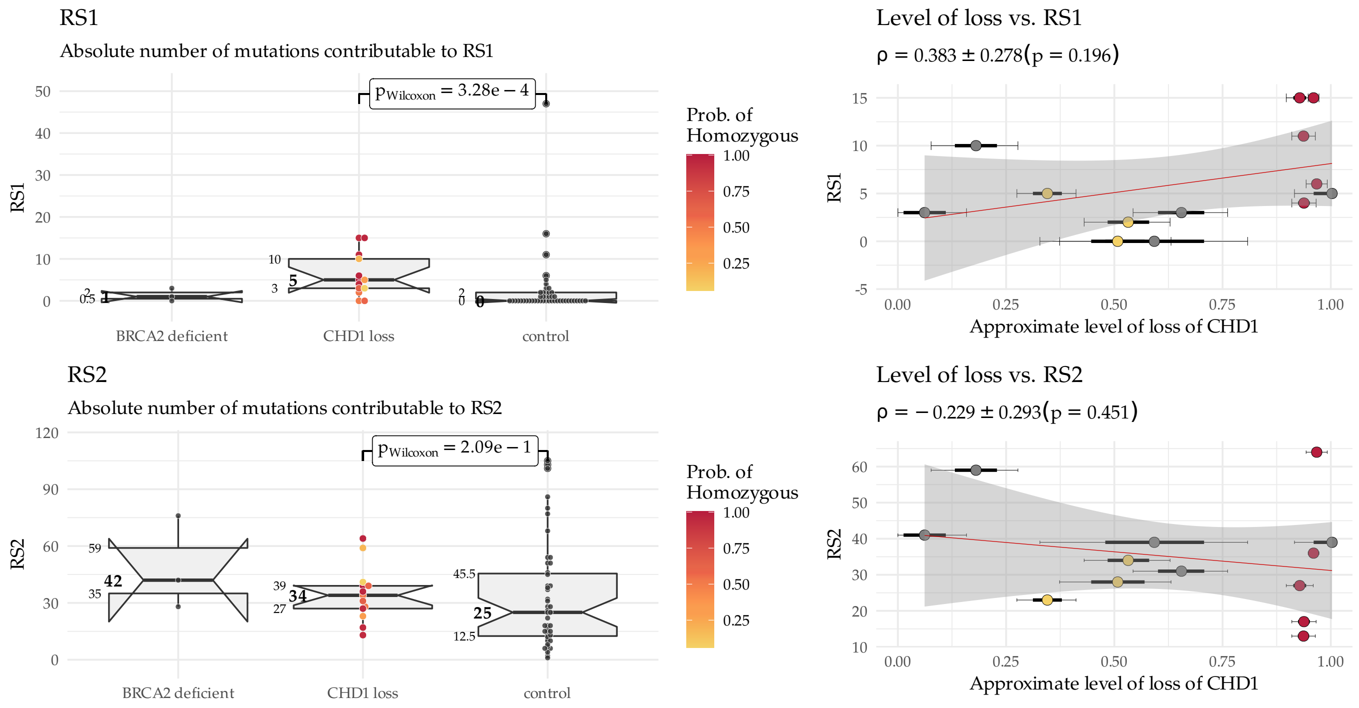

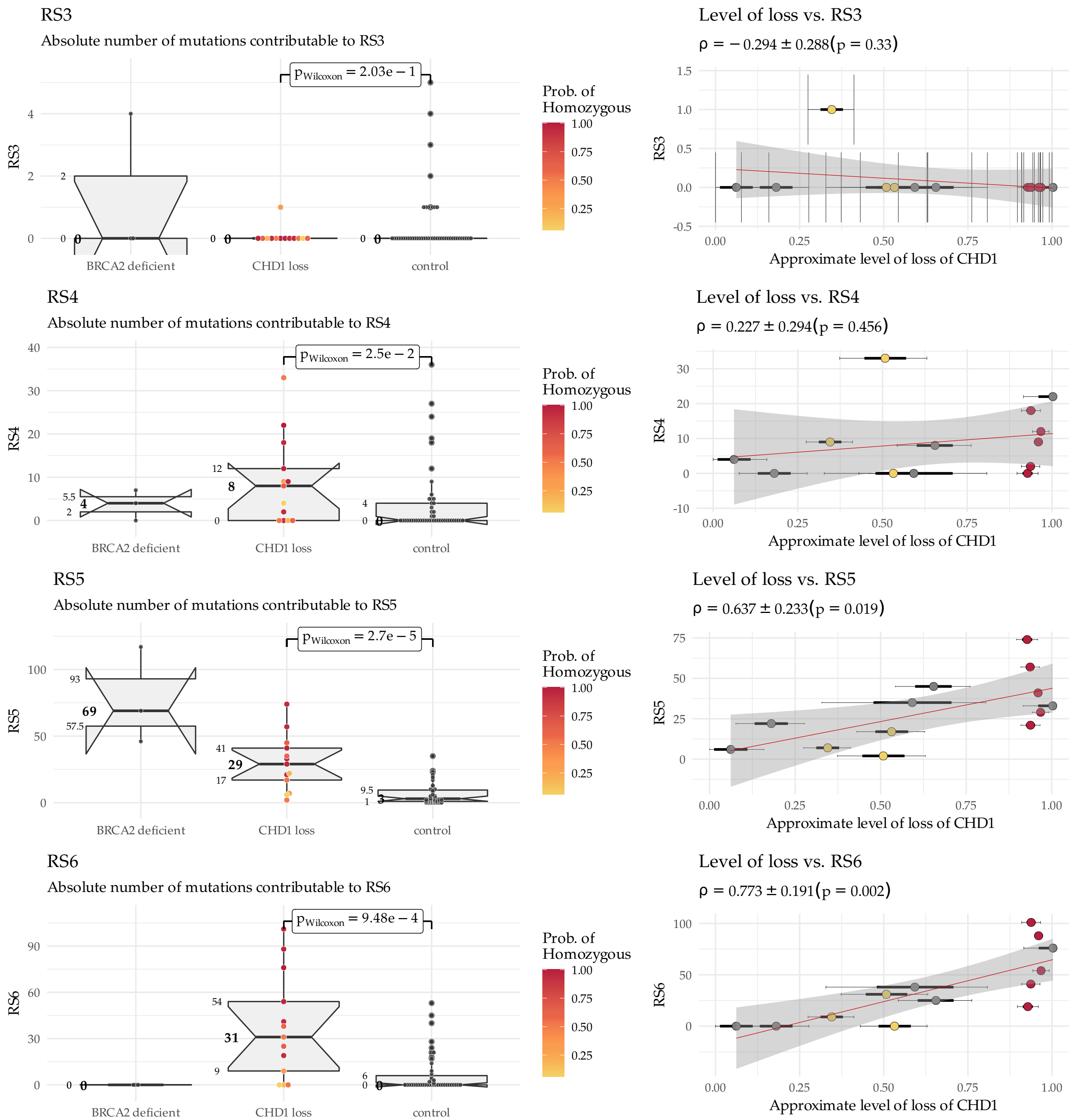
RS3
Absolute number of mutations contributable to RS3
$p_{Wilcoxon} = 2.03e-1$
Prob. of Homozygous
1.00
0.75
0.50
0.25
RS3
BRCA2 deficient
CHD1 loss
control
Level of loss vs. RS3
$\rho = -0.294 \pm 0.288 (p = 0.33)$
Approximate level of loss of CHD1
RS4
Absolute number of mutations contributable to RS4
$p_{Wilcoxon} = 2.5e-2$
Level of loss vs. RS4
$\rho = 0.227 \pm 0.294 (p = 0.456)$
RS5
Absolute number of mutations contributable to RS5
$p_{Wilcoxon} = 2.7e-5$
Level of loss vs. RS5
$\rho = 0.637 \pm 0.233 (p = 0.019)$
RS6
Absolute number of mutations contributable to RS6
$p_{Wilcoxon} = 9.48e-4$
Level of loss vs. RS6
$\rho = 0.773 \pm 0.191 (p = 0.002)$

#### 9.5 HRDetect

##### 9.5.1 Original HRDetect scores

The HRDetect scores were calculated by using the original HRDetect weights from Davies et al. [14]:

$$\begin{aligned} \text{intercept} &= -3.3642 \\ \text{Signature.8} &= 0.09062 \\ \text{HRD-LOH} &= 0.6666 \\ \text{RS5} &= 0.8467 \\ \text{RS3} &= 1.1532 \\ \text{Signature.3} &= 1.6114 \\ \text{mhm.del.ratio} &= 2.3977 \end{aligned}$$

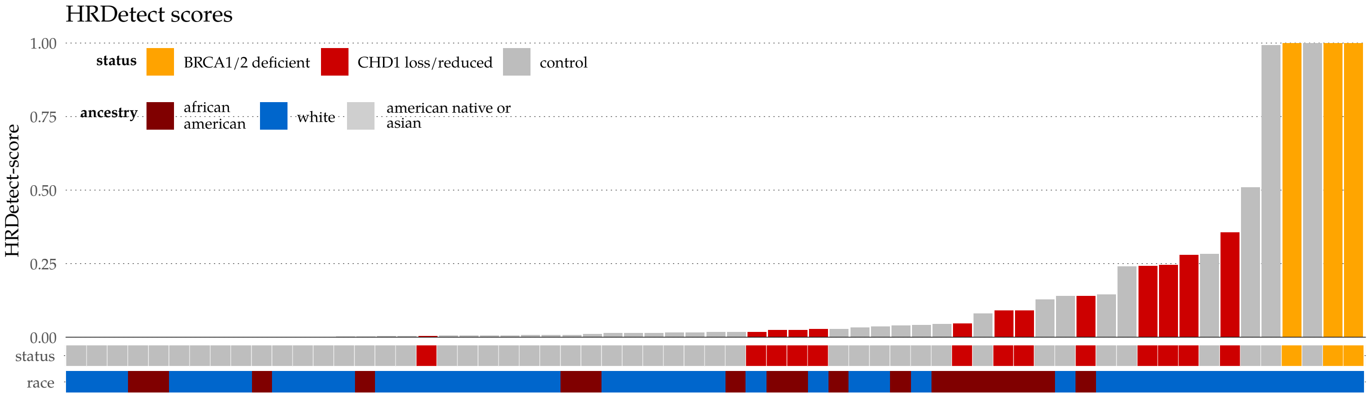

**Suppl.Fig. 36:** HRDetect score summary - WGS

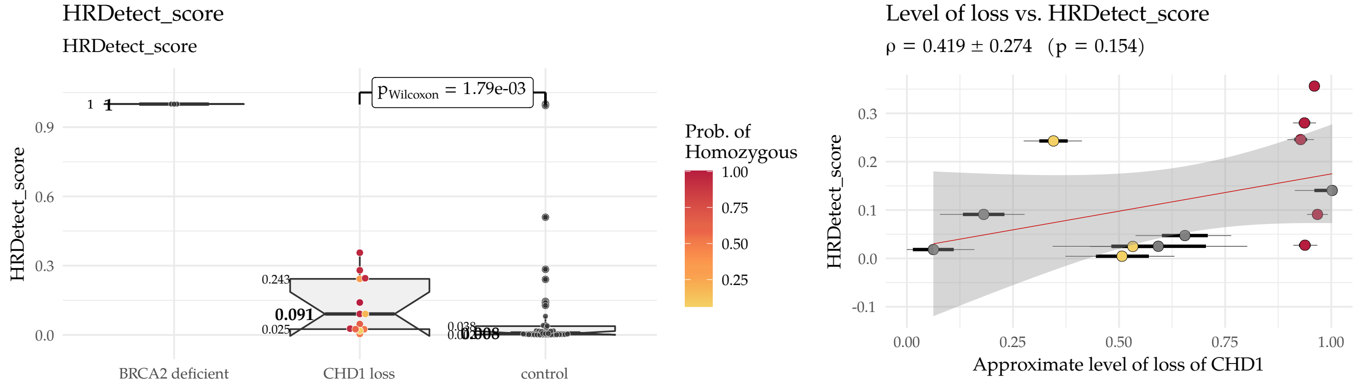

**Suppl.Fig. 37:** HRDetect scores in samples with *CHD1* loss vs. *CHD1* intact samples

##### 9.5.2 Linearized HRDetect scores

The HRDetect model is a logistic regression model, in which the ordinary linear regression scores ($\mathbf{x}$) are transformed into the range of [0,1] by putting them through a nonlinear logistic function:

$$\text{HRDetect}_{\text{probability}} = \mathbf{s} = \frac{1}{1 + e^{-\mathbf{x}}}.$$

The model is trained in a way, that these transformed scores ($\mathbf{s}$) correspond to the true probabilities that a given set of samples have a BRCA-like HR deficiency. Due to the non-linear nature of the transformation, however, the same difference in the linear scores at different regions of their domain will correspond to different changes in their corresponding HRDetect scores. In order to show that the difference between the BRCA2 mutants and the samples with *CHD1* loss is less substantial in the linear domain, we have used the logit transformation to extract the linearized, non-probabilistic HRDetect scores from the formula above:

$$\mathbf{x} = \ln\left(\frac{\mathbf{s}}{1 - \mathbf{s}}\right).$$

In addition for visualization purposes, the resulting scores were min-max normalized (mmn), i.e. their values were restricted into the [0,1] range just as before, but the linear relationship between the scores and their corresponding attributes were kept:

$$\mathbf{x}_{mmn} = \frac{\mathbf{x} - min(\mathbf{x})}{max(\mathbf{x}) - min(\mathbf{x})}.$$

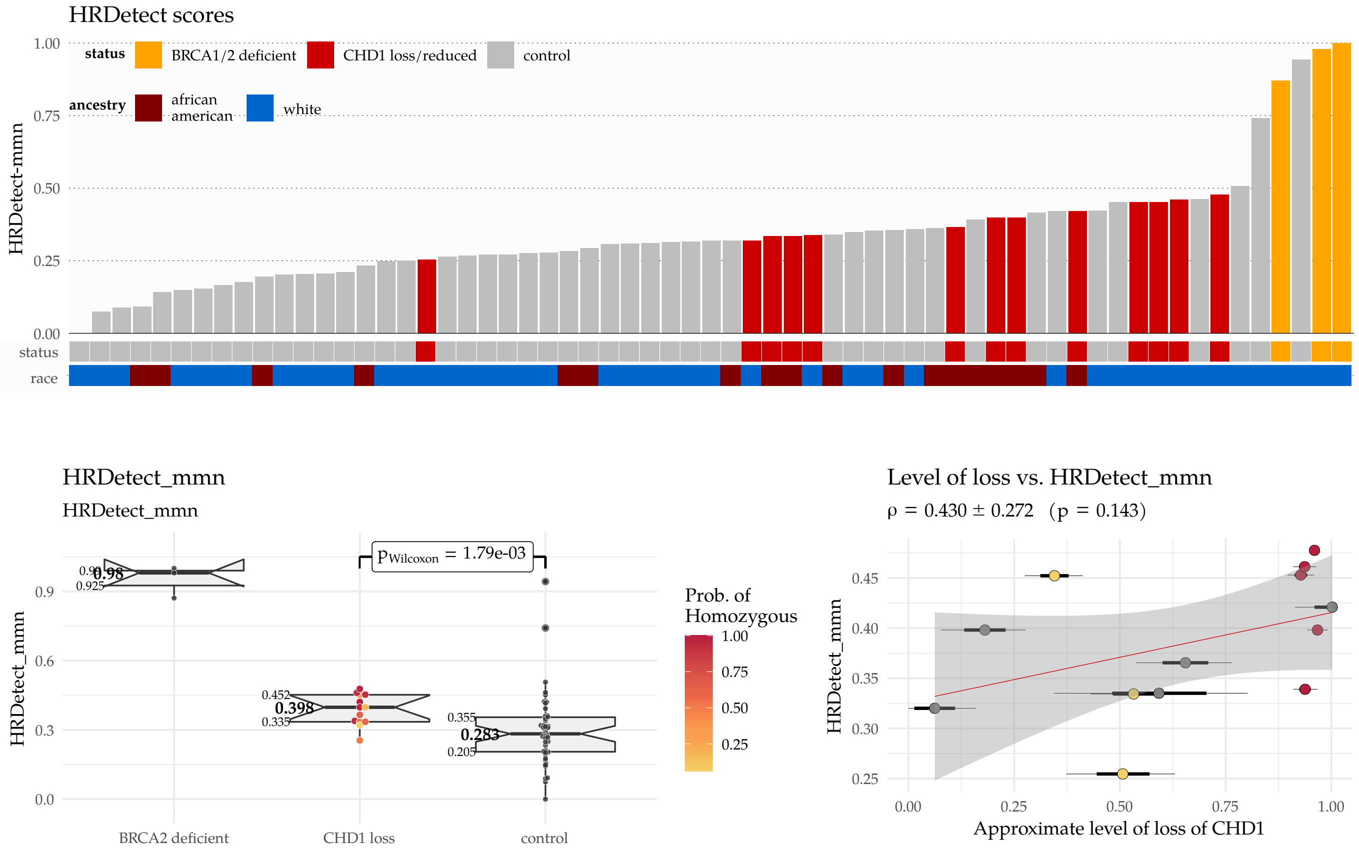

**Suppl.Fig. 38:** Linearized HRDetect score summary - WGS and the linearized HRDetect scores in samples with *CHD1* loss vs. *CHD1* intact cases

### 10 Genomic Features of the Whole Exomes

#### 10.1 Genomic Scars

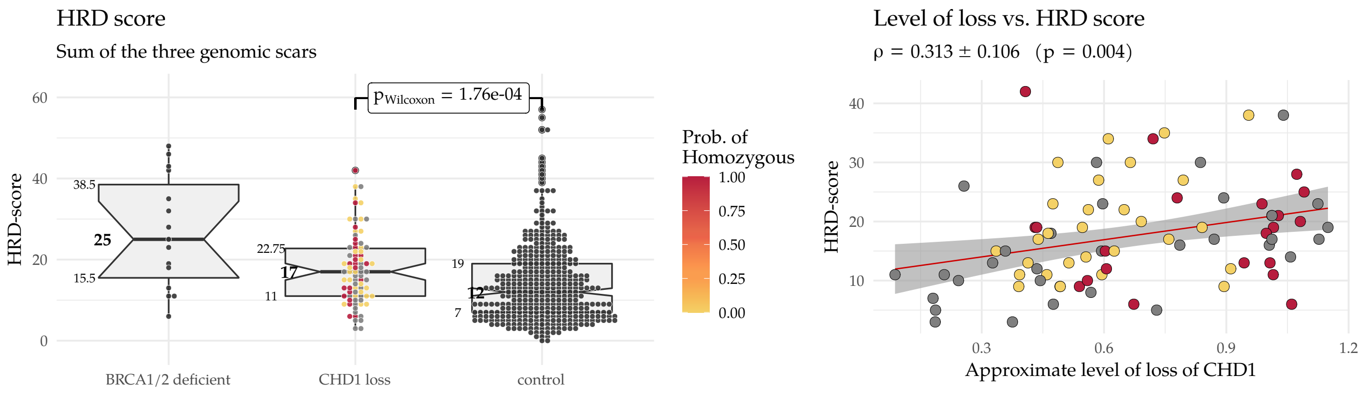

**Suppl.Fig. 39:** ↑ HRD-scores (sum of the three genomic scars) of the whole exomes

**Suppl.Fig. 40:** ↓ HRD-related genomic scars in the whole exomes. **HRD_LOH**: HRD Loss of Heterozygosity, **LST**: Large Scale Transition, **ntAI**: number of telomeric Allelic Imbalance

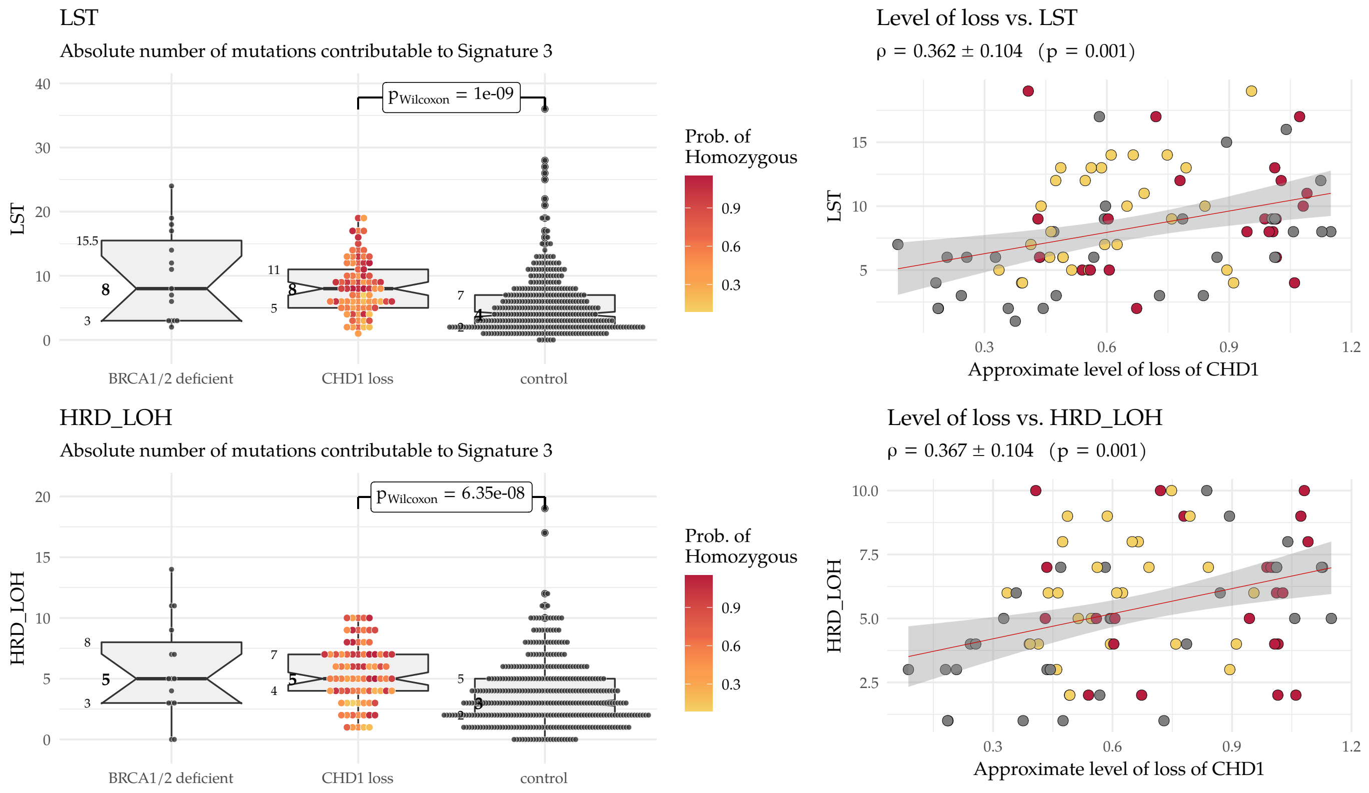

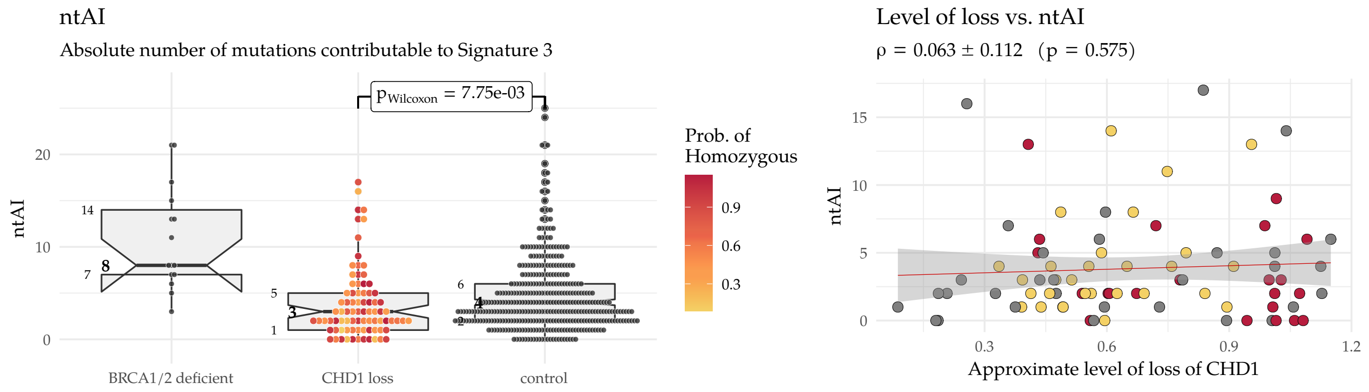

#### 10.2 Somatic SNV Signatures

**Suppl.Fig. 41:** Number of variants contributable ot the 2nd generation single nucleotide signatures – WES. Signatures 1,2,3,5,6,8 and the total number of SNVs in the sample (numSNV)

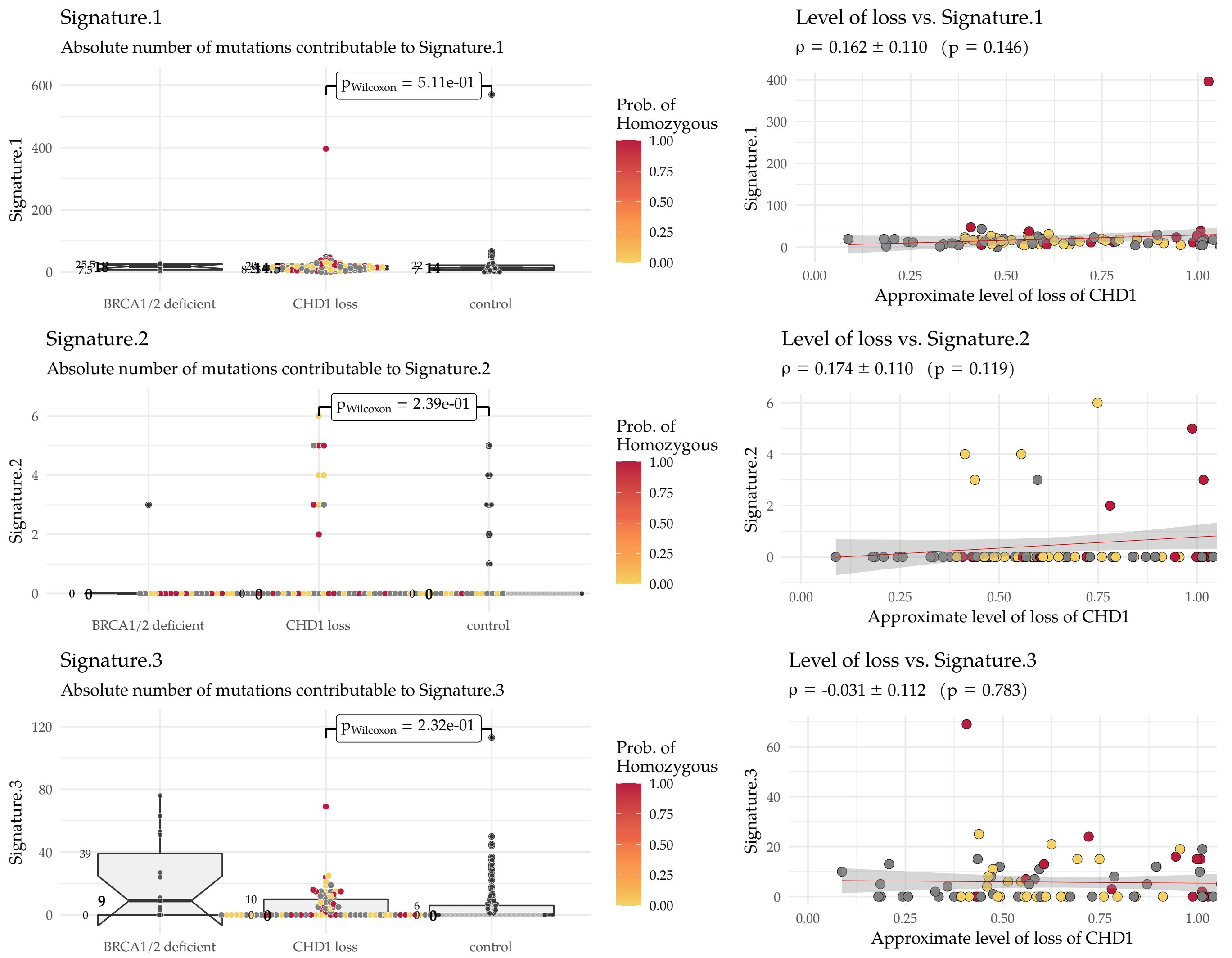

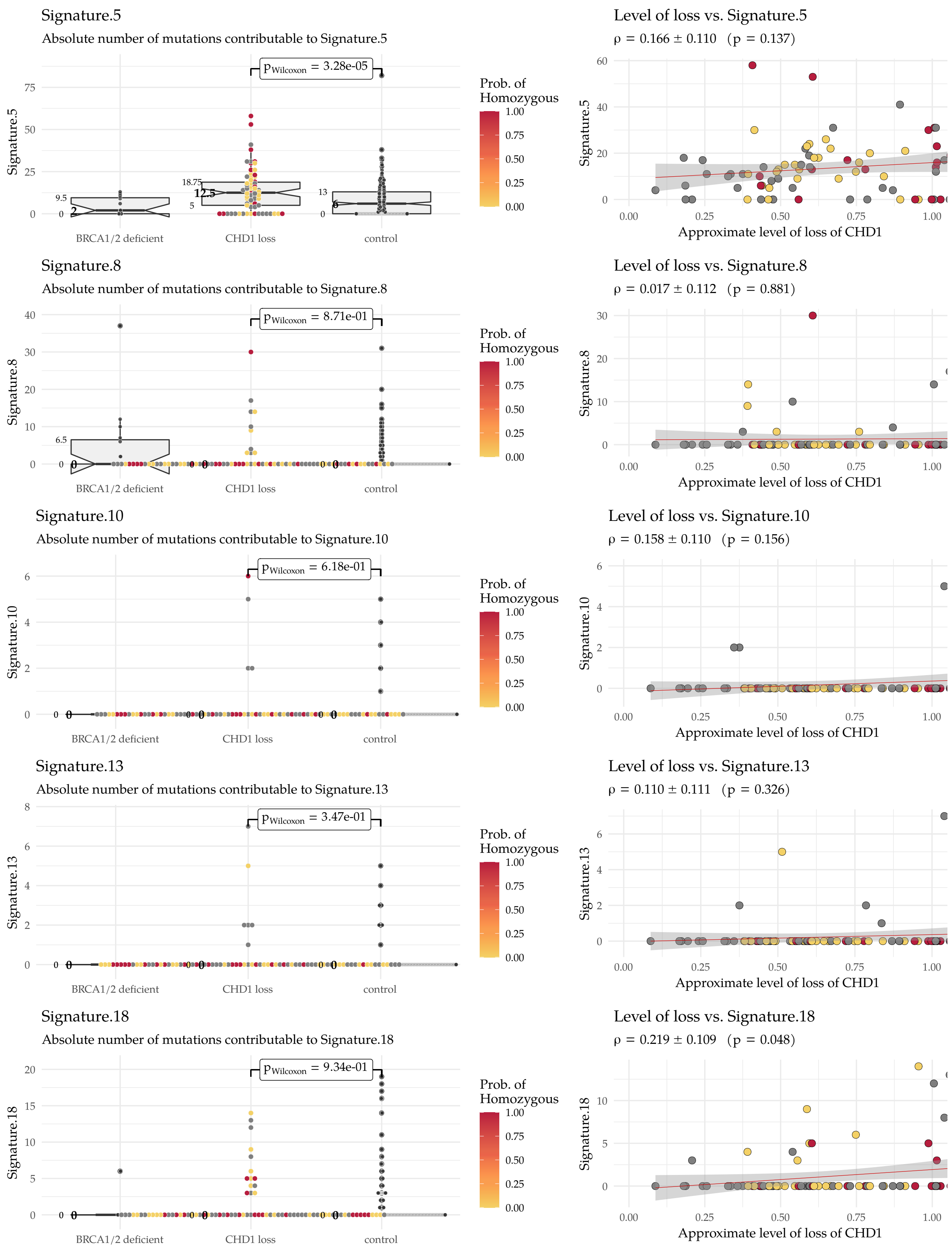

Signature.5
Absolute number of mutations contributable to Signature.5
$p_{Wilcoxon}$ = 3.28e-05
Signature.5
BRCA1/2 deficient
CHD1 loss
control
Prob. of Homozygous
Level of loss vs. Signature.5
ρ = 0.166 ± 0.110 (p = 0.137)
Approximate level of loss of CHD1
Signature.8
Absolute number of mutations contributable to Signature.8
$p_{Wilcoxon}$ = 8.71e-01
Level of loss vs. Signature.8
ρ = 0.017 ± 0.112 (p = 0.881)
Signature.10
Absolute number of mutations contributable to Signature.10
$p_{Wilcoxon}$ = 6.18e-01
Level of loss vs. Signature.10
ρ = 0.158 ± 0.110 (p = 0.156)
Signature.13
Absolute number of mutations contributable to Signature.13
$p_{Wilcoxon}$ = 3.47e-01
Level of loss vs. Signature.13
ρ = 0.110 ± 0.111 (p = 0.326)
Signature.18
Absolute number of mutations contributable to Signature.18
$p_{Wilcoxon}$ = 9.34e-01
Level of loss vs. Signature.18
ρ = 0.219 ± 0.109 (p = 0.048)

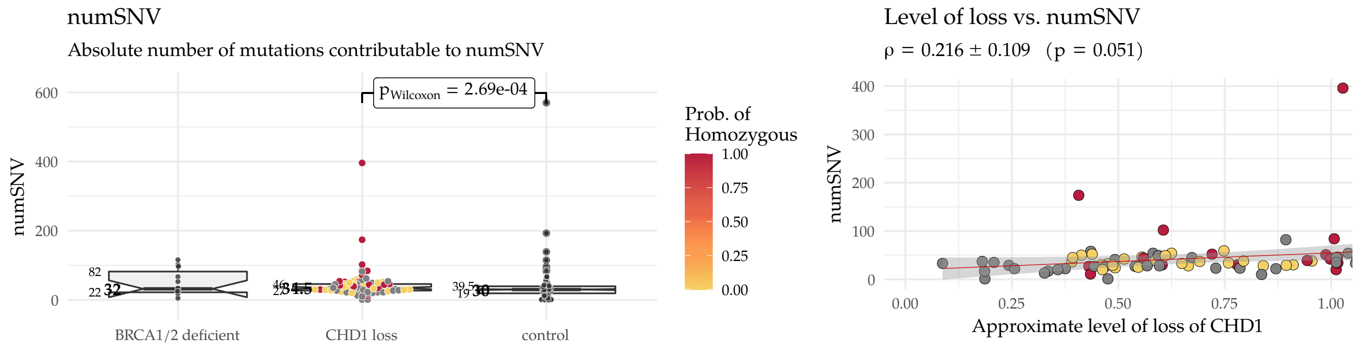

#### 10.3 Deletions

**Suppl.Fig. 42:** The "traditional" classification of deletions. ndel: number of deletions, del_ins_ratio: deletion insertion ratio, microhomology: number of microhomology mediated deletions, microhomology_del_ratio: ratio of microhomology mediated deletions relative to the number of deletions – WES

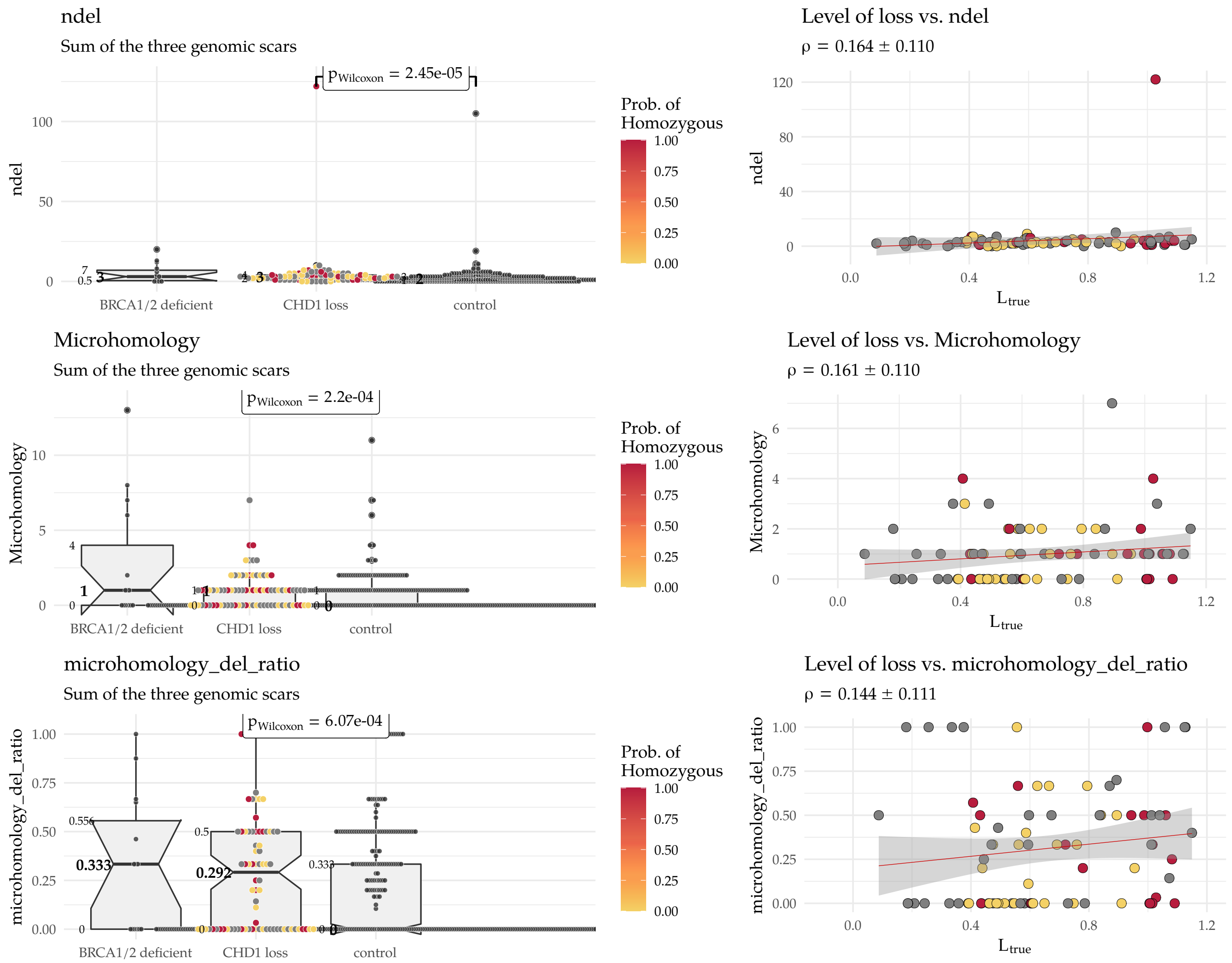

#### 10.4 HRDetect

For the whole exomes, we have used a different model, trained on 560 artificially derived (from whole genomes) breast cancer whole exomes [17]:

$$
\begin{aligned}
\text{intercept} &= -2.6192939 \\
\text{Signature.17} &= 0.067098 \\
\text{Signature.20} &= 0.09409 \\
\text{Signature.26} &= 0.16166 \\
\text{Signature.6} &= 0.310146 \\
\text{Signature.18} &= 0.31205 \\
\text{mhm.del.ratio} &= 0.314225 \\
\text{Signature.8} &= 0.61474 \\
\text{Signature.13} &= 0.83017 \\
\text{Signature.3} &= 2.00757 \\
\text{HRD-LOH} &= 2.3865
\end{aligned}
$$

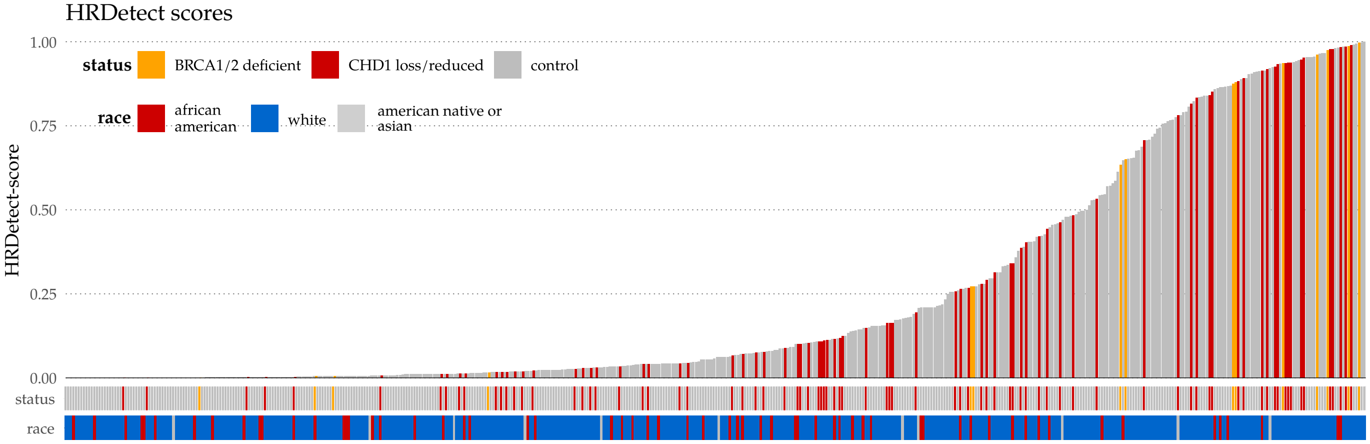

**Suppl.Fig. 43:** HRDetect score summary – WGS

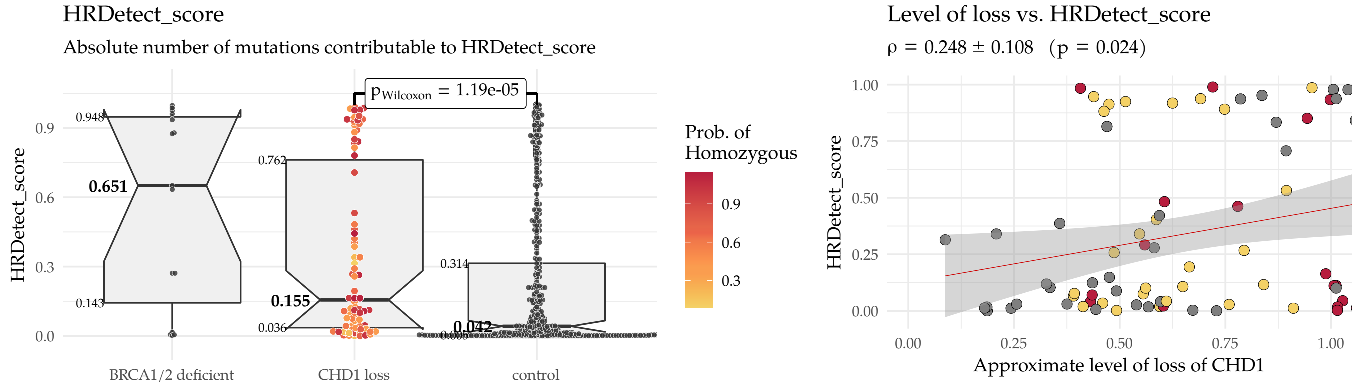

**Suppl.Fig. 44:** HRDetect scores in samples with *CHD1* loss vs. *CHD1* intact samples

### 11 Cell Line Cultures

#### 11.1 Illustartion of the experiment

CHD1 knock out (ko) was induced in the parental PC-3 and 22Rv1 cell lines. The *CHD1* ko cell populations were single cell cloned. Isogenic cell lines displaying homozygous *CHD1* ko were identified. DNA was extracted directly from the regenerated population (22Rv1_1 and PC3_1, low passage stage). Cells were further propagated through 45 generations, then the high passage cell populations were single cell cloned. DNA was extracted from two isogenic *CHD1* ko clones for each cell line (22Rv1_2, 22Rv1_3, and PC3_2, PC3_3 high passage stage) after propagation. The process is illustrated in Suppl. Figure 45. Normal references were downloaded from the Sequence Read Archive (SRA, 22Rv1: SRX5437595, PC-3: SRX5466646)

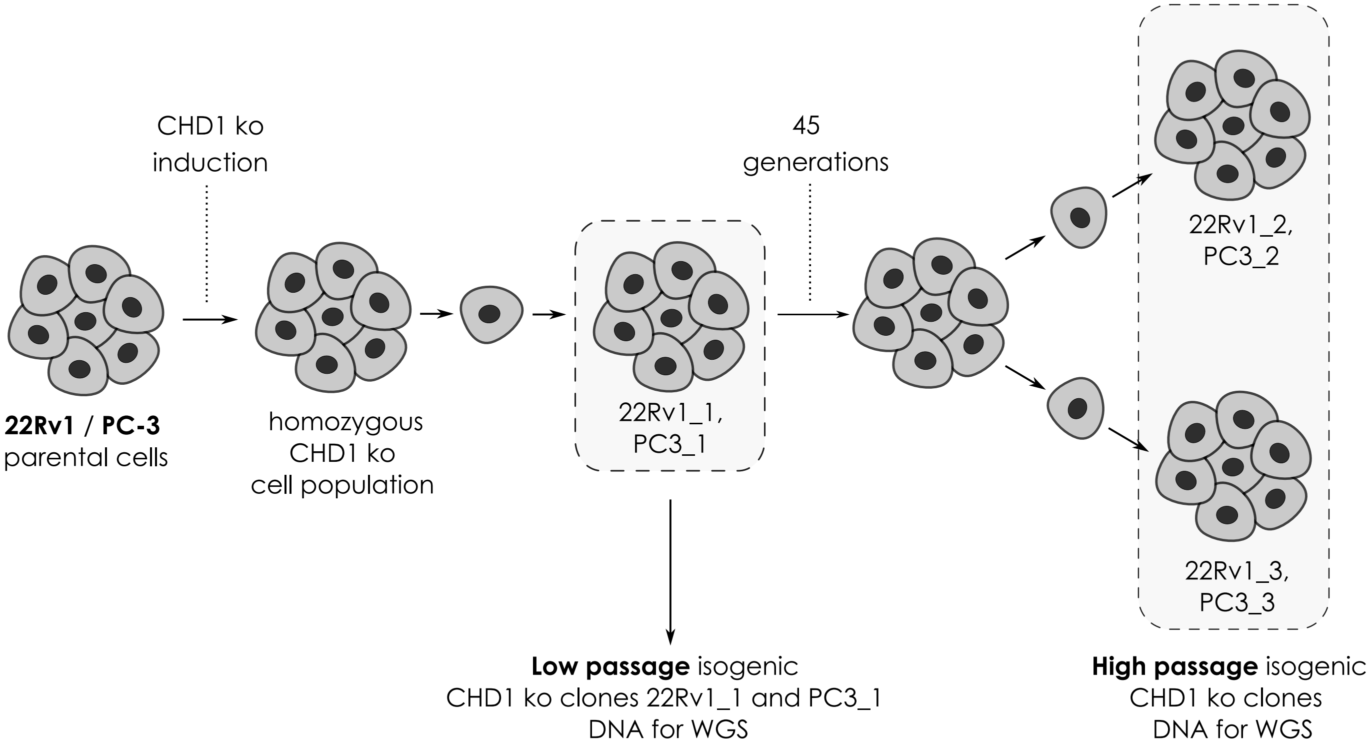

**Suppl.Fig. 45:** Illustration of the PC-3 and 22Rv1 *CHD1* ko isogenic clone generation for WGS

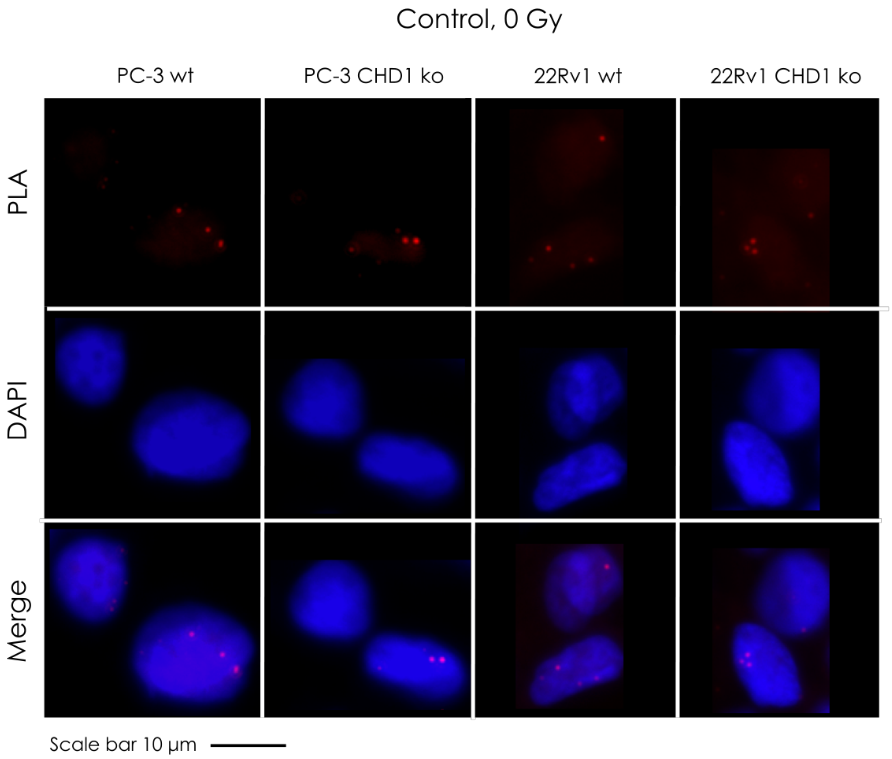

**Suppl.Fig. 46:** RAD51 foci formation. Examples of the most common staining patterns in WT and *CHD1* ko 22Rv1 and PC-3 cell lines. Control cells were fixed by 4% PFA without irradiation (IR=0Gy). PLA was carried out using antibodies against $\gamma$H2Ax and RAD51 proteins.

#### 11.2 22Rv1 cells response to HR-deficiency directed therapy

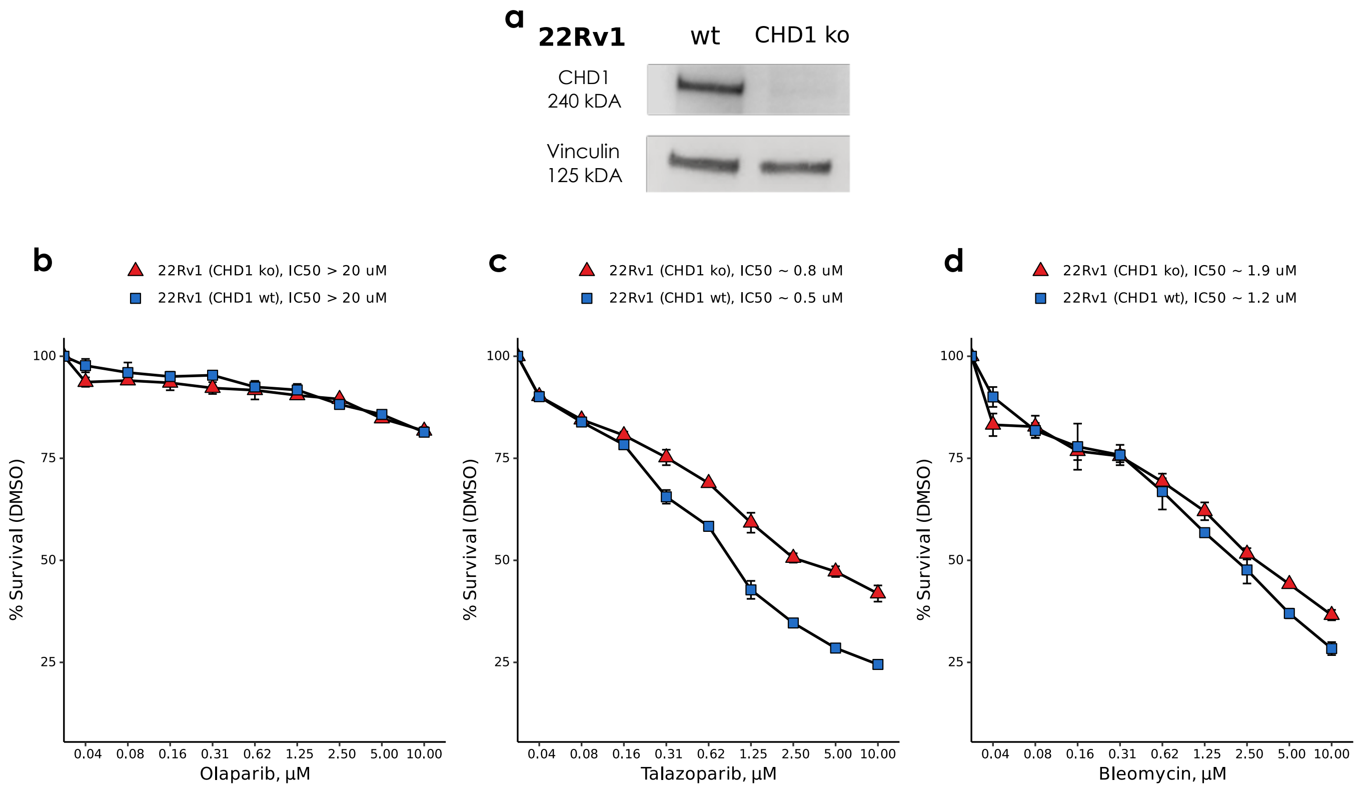

**Suppl.Fig. 47:** Immunoblot shows that *CHD1* was successfully knocked out in 22Rv1 cells (**a**). Sensitivity assays of parental wt and *CHD1* ko clones to PARP inhibitor Olaparib (**b**), Talazoparib (**b**), and the radiomimetics bleomycin sulfate (**d**). Cells viability was measured using PresoBlue$^{TM}$ reagent. SD of triplicates are shown, p-values were calculated using student's t-test. p-values $< 0.05$ were considered statistically significant.

[3] Axel Merseburger, Markus Kuczyk, Jurgen Serth, Carsten Bokemeyer, Denise Young, Leon Sun, Roger Connelly, David McLeod, Fathollah Mostofi, Shiv Srivastava, A. Stenzl, Judd Moul, and Isabell Sesterhenn. Limitations of tissue microarrays in the evaluation of focal alterations of bcl-2 and p53 in whole mount derived prostate tissues. *Oncology Reports*, January 2003.

[4] B Furusato, S-H Tan, D Young, A Dobi, C Sun, A A Mohamed, R Thangapazham, Y Chen, G McMaster, T Sreenath, G Petrovics, D G McLeod, S Srivastava, and I A Sesterhenn. ERG oncoprotein expression in prostate cancer: clonal progression of ERG-positive tumor cells and potential for ERG-based stratification. *Prostate Cancer and Prostatic Diseases*, 13(3):228–237, June 2010.

[5] B. Decker, D. M. Karyadi, B. W. Davis, E. Karlins, L. S. Tillmans, J. L. Stanford, S. N. Thibodeau, and E. A. Ostrander. Biallelic BRCA2 Mutations Shape the Somatic Mutational Landscape of Aggressive Prostate Tumors. *Am. J. Hum. Genet.*, 98(5):818–829, 05 2016.

[6] Tesfaye B Mersha and Tilahun Abebe. Self-reported race/ethnicity in the age of genomic research: its potential impact on understanding health disparities. *Human Genomics*, 9(1), January 2015.

[7] Konrad J. Karczewski, Ben Weisburd, Brett Thomas, Matthew Solomonson, Douglas M. Ruderfer, David Kavanagh, Tymor Hamamsy, Monkol Lek, Kaitlin E. Samocha, Beryl B. Cummings, Daniel Birnbaum, Mark J. Daly, and Daniel G. MacArthur. The ExAC browser: displaying reference data information from over 60 000 exomes. *Nucleic Acids Research*, 45(D1):D840–D845, November 2016.

[13] Ludmil B. Alexandrov, , Jaegil Kim, Nicholas J. Haradhvala, Mi Ni Huang, Alvin Wei Tian Ng, Yang Wu, Arnoud Boot, Kyle R. Covington, Dmitry A. Gordenin, Erik N. Bergstrom, S. M. Ashiqul Islam, Nuria Lopez-Bigas, Leszek J. Klimczak, John R. McPherson, Sandro Morganella, Radhakrishnan Sabarinathan, David A.

Wheeler, Ville Mustonen, Gad Getz, Steven G. Rozen, and Michael R. Stratton and. The repertoire of mutational signatures in human cancer. *Nature*, 578(7793):94–101, February 2020.

[14] Helen Davies, Dominik Glodzik, Sandro Morganella, Lucy R Yates, Johan Staaf, Xueqing Zou, Manasa Ramakrishna, Sancha Martin, Sandrine Boyault, Anieta M Sieuwerts, Peter T Simpson, Tari A King, Keiran Raine, Jorunn E Eyfjord, Gu Kong, Åke Borg, Ewan Birney, Hendrik G Stunnenberg, Marc J van de Vijver, Anne-Lise Børresen-Dale, John W M Martens, Paul N Span, Sunil R Lakhani, Anne Vincent-Salomon, Christos Sotiriou, Andrew Tutt, Alastair M Thompson, Steven Van Laere, Andrea L Richardson, Alain Viari, Peter J Campbell, Michael R Stratton, and Serena Nik-Zainal. HRDetect is a predictor of BRCA1 and BRCA2 deficiency based on mutational signatures. *Nature Medicine*, 23(4):517–525, March 2017.

[15] Ludmil B Alexandrov and Michael R Stratton. Mutational signatures: the patterns of somatic mutations hidden in cancer genomes. *Current Opinion in Genetics & Development*, 24:52–60, feb 2014.

[16] Arnoud Boot, Mi Ni Huang, Alvin W.T. Ng, Szu-Chi Ho, Jing Quan Lim, Yoshiiku Kawakami, Kazuaki Chayama, Bin Tean Teh, Hidewaki Nakagawa, and Steven G. Rozen. In-depth characterization of the cisplatin mutational signature in human cell lines and in esophageal and liver tumors. *Genome Research*, 28(5):654–665, April 2018.

[17] M. Diossy, L. Reiniger, Z. Sztupinszki, M. Krzystanek, K.M. Timms, C. Neff, C. Solimeno, D. Pruss, A.C. Eklund, E. Tóth, O. Kiss, O. Rusz, G. Cserni, T. Zombori, B. Székely, J. Kulka, J. Tímár, I. Csabai, and Z. Szallasi. Breast cancer brain metastases show increased levels of genomic aberration-based homologous recombination deficiency scores relative to their corresponding primary tumors. *Annals of Oncology*, 29(9):1948–1954, September 2018.

[18] Jia Zhou, Jiaqi Li, Rodolfo B Serafim, Steven Ketchum, Catarina G Ferreira, Jessica C Liu, Kathryn A Coe, Brendan D Price, and Timur Yusufzai. Human CHD1 is required for early DNA-damage signaling and is uniquely regulated by its n terminus. *Nucleic Acids Research*, 46(8):3891–3905, February 2018.
